# Incident heart failure and myocardial infarction in sodium-glucose cotransporter 2 versus dipeptidyl peptidase-4 inhibitor users

**DOI:** 10.1101/2021.11.21.21266648

**Authors:** Jiandong Zhou, Sharen Lee, Keith Sai Kit Leung, Abraham Ka Chung Wai, Tong Liu, Ying Liu, Dong Chang, Wing Tak Wong, Ian Chi Kei Wong, Bernard Man Yung Cheung, Qingpeng Zhang, Gary Tse

## Abstract

**Objectives:** To compare the rates of major cardiovascular adverse events in sodium-glucose cotransporter-2 inhibitors (SGLT2I) and dipeptidyl-peptidase-4 inhibitors (DPP4I) users in a Chinese population.

**Background:** SGLT2I and DPP4I are increasingly prescribed for type 2 diabetes mellitus patients. However, few population-based studies are comparing their effects on incident heart failure or myocardial infarction.

**Methods:** This was a population-based retrospective cohort study using the electronic health record database in Hong Kong, including type 2 diabetes mellitus patients receiving either SGLT2I or DPP4I between January 1st, 2015, to December 31st, 2020. Propensity-score matching was performed in a 1:1 ratio based on demographics, past comorbidities, non-SGLT2I/DPP4I medications with nearest-neighbor matching (caliper=0.1). Univariable and multivariable Cox models were used to identify significant predictors for new onset heart failure, new onset myocardial infarction, cardiovascular mortality, and all-cause mortality. Sensitivity analyses with competing risk models and multiple propensity score matching approaches were conducted. Subgroup age and gender analyses were presented.

**Results:** A total of 41994 patients (58.89% males, median admission age at 58 years old, interquartile rage [IQR]: 51.2-65.3) were included in the study cohorts with a median follow-up duration of 5.6 years (IQR: 5.32-5.82). After adjusting for significant demographics, past comorbidities, medication prescriptions and biochemical results, SGLT2I users have a significantly lower risk for myocardial infarction (hazard ratio [HR]: 0.34, 95% confidence interval [CI]: [0.28, 0.41], P < 0.0001), cardiovascular mortality (HR: 0.53, 95% CI: [0.38, 0.74], P = 0.0002) and all-cause mortality (HR: 0.21, 95% CI: [0.18, 0.25], P= 0.0001) under multivariable Cox regression. However, the risk for heart failure is comparable (HR: 0.87, 95% CI: [0.73, 1.04], P= 0.1343).

**Conclusions:** SGLT2 inhibitors are protective against adverse cardiovascular events compared to DPP4I. The prescription of SGLT2I is preferred especially for males and patients aged 65 or older to prevent cardiovascular risks.

## Introduction

Diabetes mellitus is an increasingly prevalent metabolic disease, currently affecting more than 400 million people, and the patient population is projected to increase up to 642 million by 2040. (1) Given the ever-increasing disease burden, new classes of antidiabetic agents have been introduced into the market over the past decade. The use of two novel classes of antidiabetic agents-sodium glucose cotransporter-2 inhibitors (SGLT2I) and dipeptidyl-peptidase-4 inhibitors (DPP4I) have increased significantly. (2, 3) Besides their favorable side effect profile, studies have reported beneficial effects on metabolic risk from these two classes of drugs (4). Based on findings from large-scale clinical trials, the cardiovascular mortality-lowering effects of SGLT2I are mostly attributed to its protection against heart failure (HF). (5-8) On the other hand, the cardiovascular effect of DPP4I appears to be more controversial. Whilst there were reports of DPP4I users having lower cardiovascular risks than non-users, there are also studies reporting an increased risk of HF in saxagliptin users. (9, 10)

Whilst small-scale trials are comparing the metabolic effects or specific disease outcomes of SGLT2I and DPP4I, there is a lack of large-scale population studies to evaluate the difference in the presentation of major cardiovascular adverse events between the use of the two drug classes. (11-13) Recently, Zheng *et al*. has demonstrated lower mortality in SGLT2I users in comparison to DPP4I users in a network meta-analysis. (14) However, ultimately the study is limited by the indirect comparison of the SGLT2I and DPP4I users. Other studies have reported on outcomes such as weight loss, improvement in the liver or renal function (15) and reduction in atrial fibrillation incidence (16). Another study recently investigated cardiovascular outcomes such as HF and MI, but only in Japanese, Korean and European cohorts (17). Therefore, the aim of the present study is to compare the occurrence of major cardiovascular adverse events in SGLT2I and DPP4I users to evaluate their cardiovascular protective effects in a Chinese population.

## Methods

### Study design and population

This study was approved by the Institutional Review Board of the University of Hong Kong/Hospital Authority Hong Kong West Cluster and from The Joint Chinese University of Hong Kong–New Territories East Cluster Clinical Research Ethics Committee. It included type 2 diabetes mellitus patients with SGLT2I or DPP4I prescriptions from January 1^st^, 2015 to December 31^st^, 2020. Patients who received both DPP4I and SGLT2I, in addition to patients who discontinued the medication during the study were excluded. The exclusion criteria for the HF study cohort were as follows: patients with prior HF diagnosis or with the use of medications for HF (e.g., diuretics for HF, beta-blockers for HF). For the MI study cohort, patients with prior old MI or MI diagnosis were excluded. The patients were identified from the Clinical Data Analysis and Reporting System (CDARS), a territory-wide database that centralizes patient information from individual local hospitals to establish comprehensive medical data, including clinical characteristics, disease diagnosis, laboratory results, and drug treatment details. The system has been previously used by both our team and other teams in Hong Kong to conduct population-based cohort studies (18, 19), including those on diabetes mellitus (20, 21).

This study made use of the Clinical and biochemical data were extracted for the present study. Patients’ demographics include sex and age of initial drug use (baseline). Prior comorbidities before initial drug use were extracted, including diabetes with chronic complication, diabetes without chronic complication, gout, hypertension, immune mediated enterocolitis, ischemic heart disease, liver diseases, peripheral vascular disease, renal diseases, stroke/transient ischemic attack, general symptoms, atrial fibrillation, ventricular tachycardia (VT)/ ventricular fibrillation (VF)/ sudden cardiac death (SCD), anemia, overweight, cancer. Charlson’s standard comorbidity index was also calculated. Mortality was recorded using the International Classification of Diseases Tenth Edition (ICD-10) coding, whilst the study outcomes and comorbidities were documented in CDARS under ICD-9 codes. **Supplementary Table 1** displays the ICD codes used to search for patient outcomes and comorbidities.

Non-SGLT2I/DPP4I medications were also extracted, including metformin, sulphonylurea, insulin, acarbose, thiazolidinedione, glucagon-like peptide-1 receptor agonists, and statins and fibrates. Baseline laboratory data were extracted. A limited number of enrolled patients have been prescribed calcium channel blockers; thus, they were not considered. Subclinical biomarkers were calculated accordingly, including neutrophil-to-lymphocyte ratio, platelet-to-lymphocyte ratio, neutrophil-to-high-density lipoprotein ratio, lymphocyte-to-high-density lipoprotein ratio, lymphocyte-to-low-density lipoprotein ratio, low density lipoprotein ratio-to-high density lipoprotein ratio, total cholesterol-to-high density lipoprotein ratio, triglyceride-glucose index, bilirubin-to-albumin ratio, protein-to-creatinine ratio, and prognostic nutritional index.

Standard deviation (SD) was calculated for glycemic and lipid profile parmaters once there are at least three examinations for each patient since initial drug exposure of SGLT2I or DPP4I. We also calculated more specific variability measures for HbA1c and fasting glucose profiles including SD, SD/initial, coefficient of variation (CV), and variability independent of mean as listed in **Supplementary Table 2**.

### Outcomes and statistical analysis

The study outcome are new onset HF, and new onset MI, cardiovascular mortality, and all-cause mortality as defined by the first incidence of ICD-9 codes of these adverse events(**Supplementary Table 1**). Mortality data were obtained from the Hong Kong Death Registry, a population-based official government registry with the registered death records of all Hong Kong citizens linked to CDARS. ICD-10 codes I00-I09, I11, I13, I20-I51 were used to identify cardiovascular mortality. Descriptive statistics are used to summarize baseline clinical and biochemical characteristics of patients with SGLT2I and DPP4I use. For baseline clinical characteristics, the continuous variables were presented as median (95% confidence interval [CI]/ interquartile range [IQR]) or mean (standard deviation [SD]) and the categorical variables were presented as total number (percentage). Continuous variables were compared using the two-tailed Mann-Whitney U test, whilst the two-tailed chi-square test with Yates’ correction was used to test 2×2 contingency data. Univariable Cox regression was used to identify significant predictors for the primary and secondary outcomes. Propensity-score matching was performed to generate control of SGLT2I users to compare against DPP4I users in a 1:1 ratio based on baseline age, sex, prior comorbidities, and non-SGLT2I/DPP4I medications using nearest-neighbor matching strategy. To minimize the outcome bias caused by variations in baseline characteristics, inverse probability of treatment weighting (IPTW) and stable inverse probability of treatment weighting (SIPTW) were then implemented to balance the covariables between groups using the calculated propensity scores.

Multivariable Cox analysis models after being adjusted for significant risk factors of demographics, past comorbidities, non-SGLT2I/DPP4I medications, subclinical biomarkers, HbA1c and fasting glucose to identify the treatment effects of SGLT2I v.s. DPP4I on the mentioned adverse outcomes. Cause-specific and subdistribution hazard models were conducted to consider possible competing risks. Lastly, subgroup analyses were done on age (≤65 and >65 years) and sex on drug exposure effects. A standardized mean difference (SMD) of no less than 0.2 between the treatment groups post-weighting was considered negligible. The hazard ratio (HR), 95% CI and P-value were reported. Statistical significance is defined as P-value < 0.05. The statistical analysis was performed with RStudio software (Version: 1.1.456) and Python (Version: 3.6).

## Results

### Baseline characteristics of HF and MI cohorts before and after propensity score matching

Type 2 diabetes mellitus patients with SGLT2I or DPP4I use from January 1^st^, 2015, to December 31^st^, 2020, were included (**Table 1**). Patients with the use of both classes, or with prior HF diagnoses or admissions due to HF or with anti-HF drugs (e.g., beta-blockers for HF, diuretics for HF), were excluded. After exclusion, 41994 persons (58.89% males, median admission age at 58 years old, interquartile rage [IQR]: 51.2-65.3) fulfilled the eligibility criteria in the study cohort for subsequent analysis. The study cohort has a median follow-up duration of 5.6 years (IQR: 5.32-5.82). **Supplementary Table 3, 4, 5** summarized the baseline and clinical characteristics of patients with/without HF, MI and mortality risk, respectively, before and after propensity score matching (1:1). **Supplementary Table 6** describes the baseline and clinical characteristics of the present cohort stratified by age and gender before and after propensity score matching of 1:1 ratio. **Figure 2** presented the cumulative incidence curves for HF, MI, cardiovascular mortality, and all-cause mortality stratified by drug exposure of SGLT2I v.s. DPP4I in the matched cohort, demonstrating that SGLT2I users have a significantly lower incidence of all study outcomes.

**Table 1.**
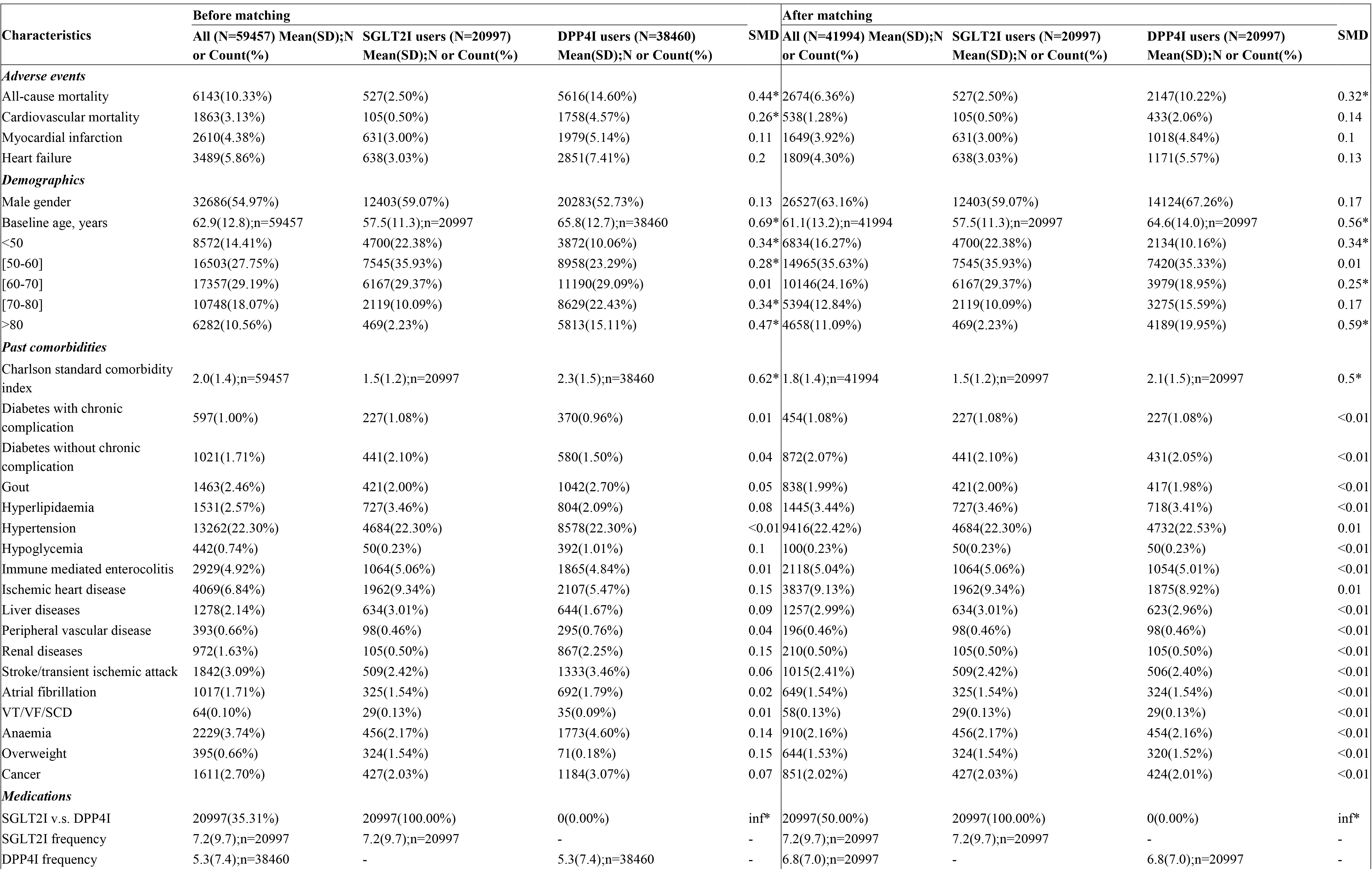
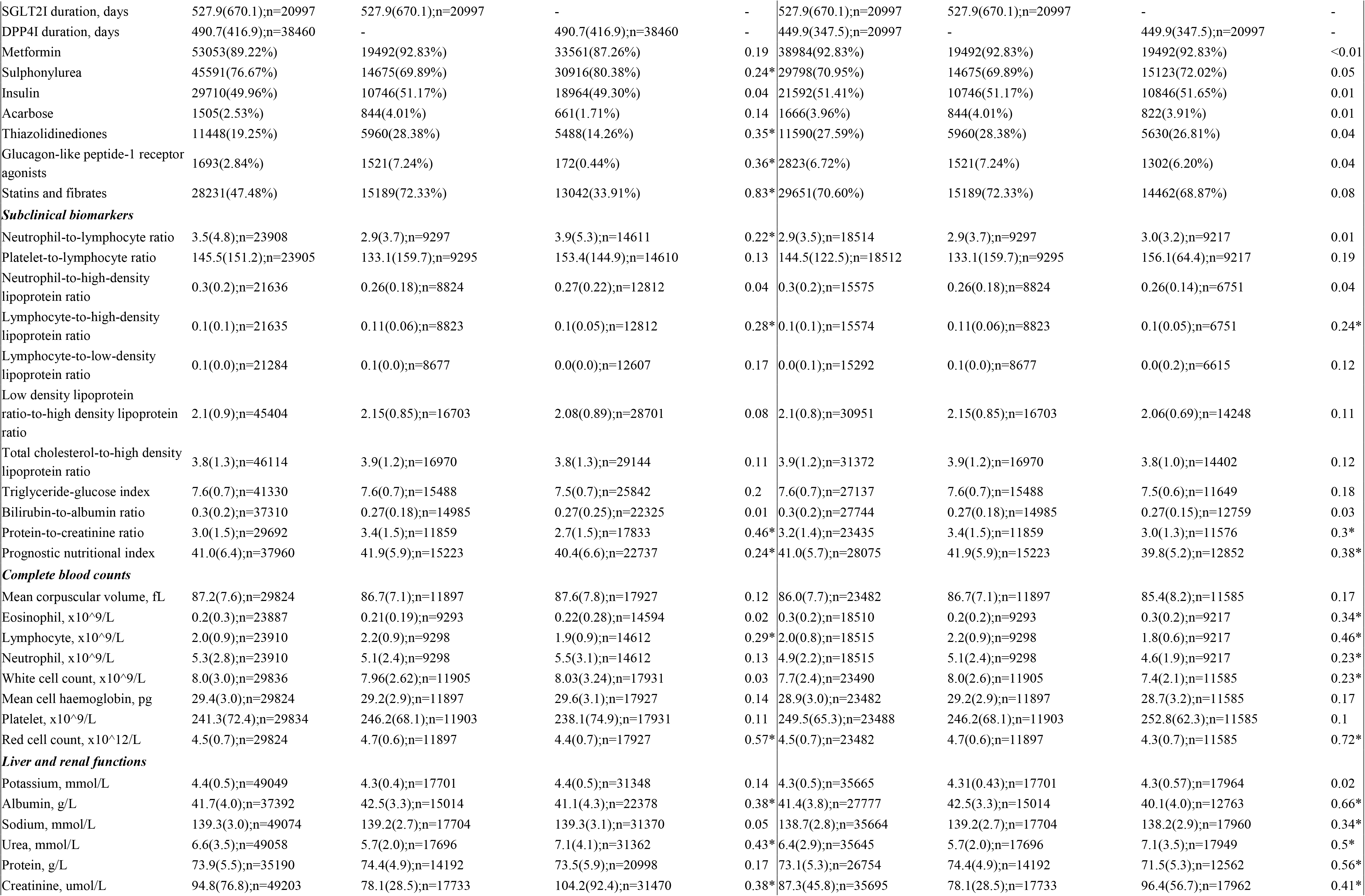
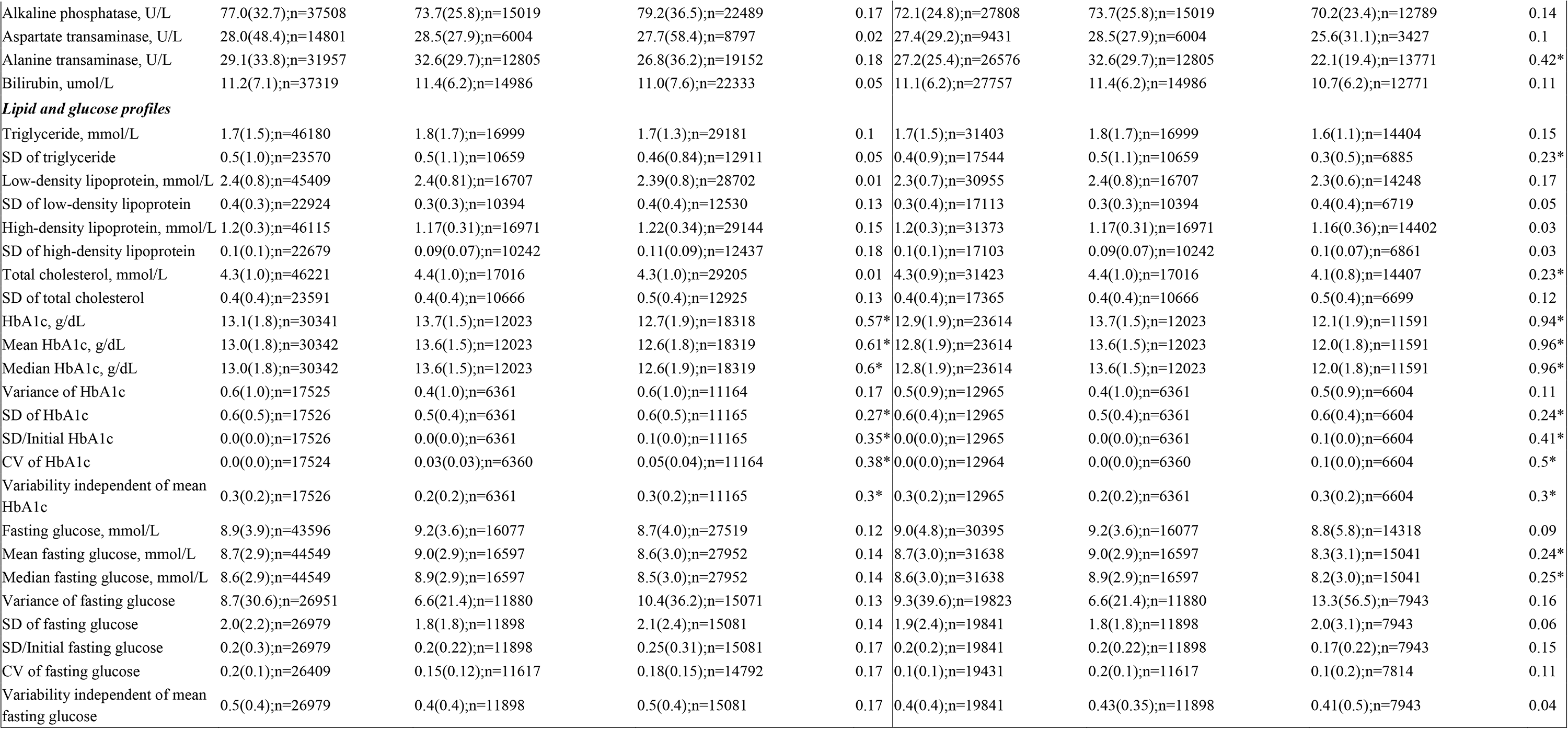
Baseline and clinical characteristics of patients with SGLT2I v.s. DPP4I use before and after propensity score matching (1:1). * for SMD≥0.2; SD: standard deviation; SCD: sudden cardiac death; VF: ventricular fibrillation; VT: ventricular tachycardia; SGLT2I: sodium glucose cotransporter-2 inhibitor; DPP4I: dipeptidyl peptidase-4 inhibitor; CV: coefficient of variation.

**Figure 1.**
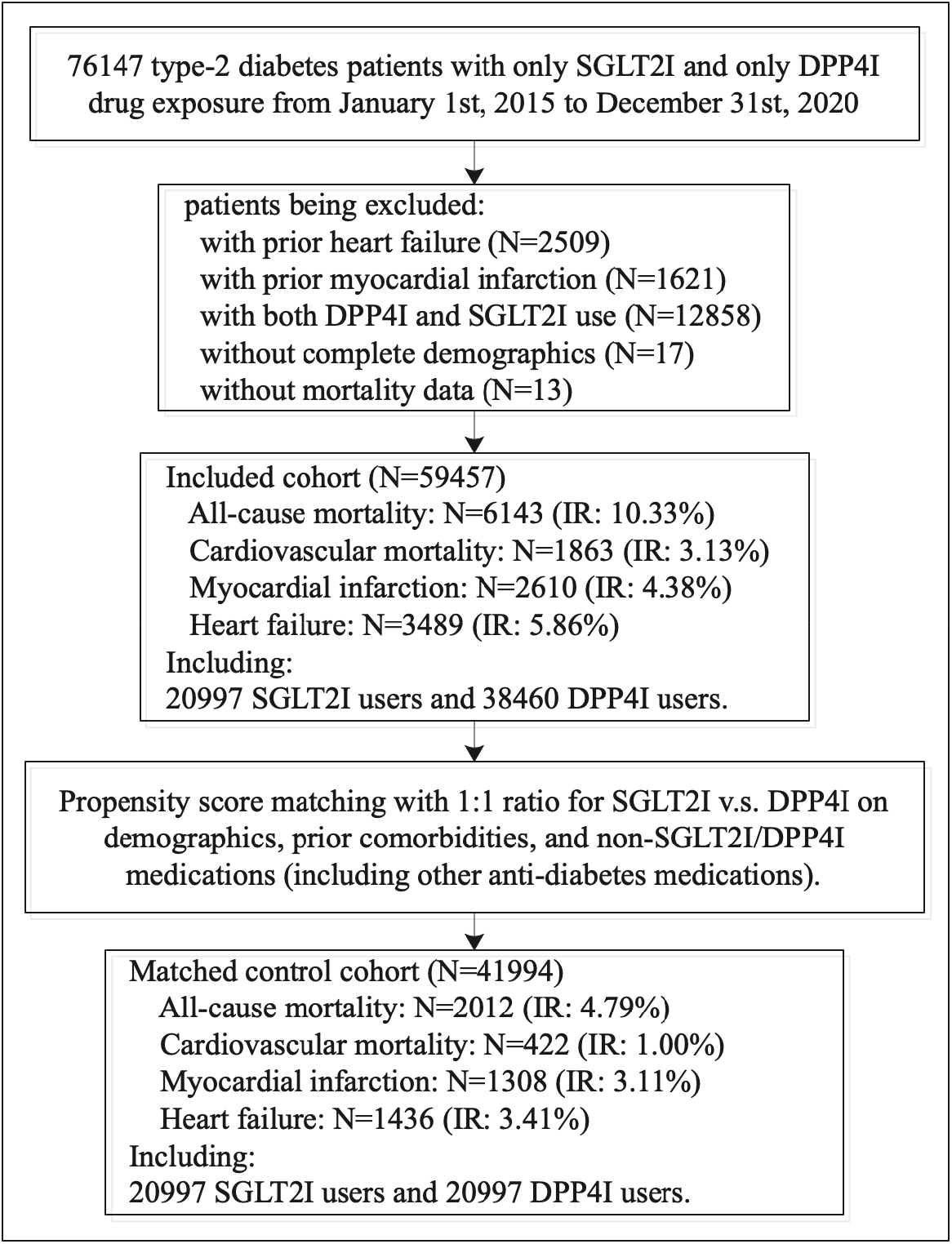
Flowchart of data processing. IR: incidence rate, SGLT2I: sodium glucose cotransporter 2 inhibitors, DPP4I: dipeptidyl peptidase-4 inhibitors.

**Figure 2.**
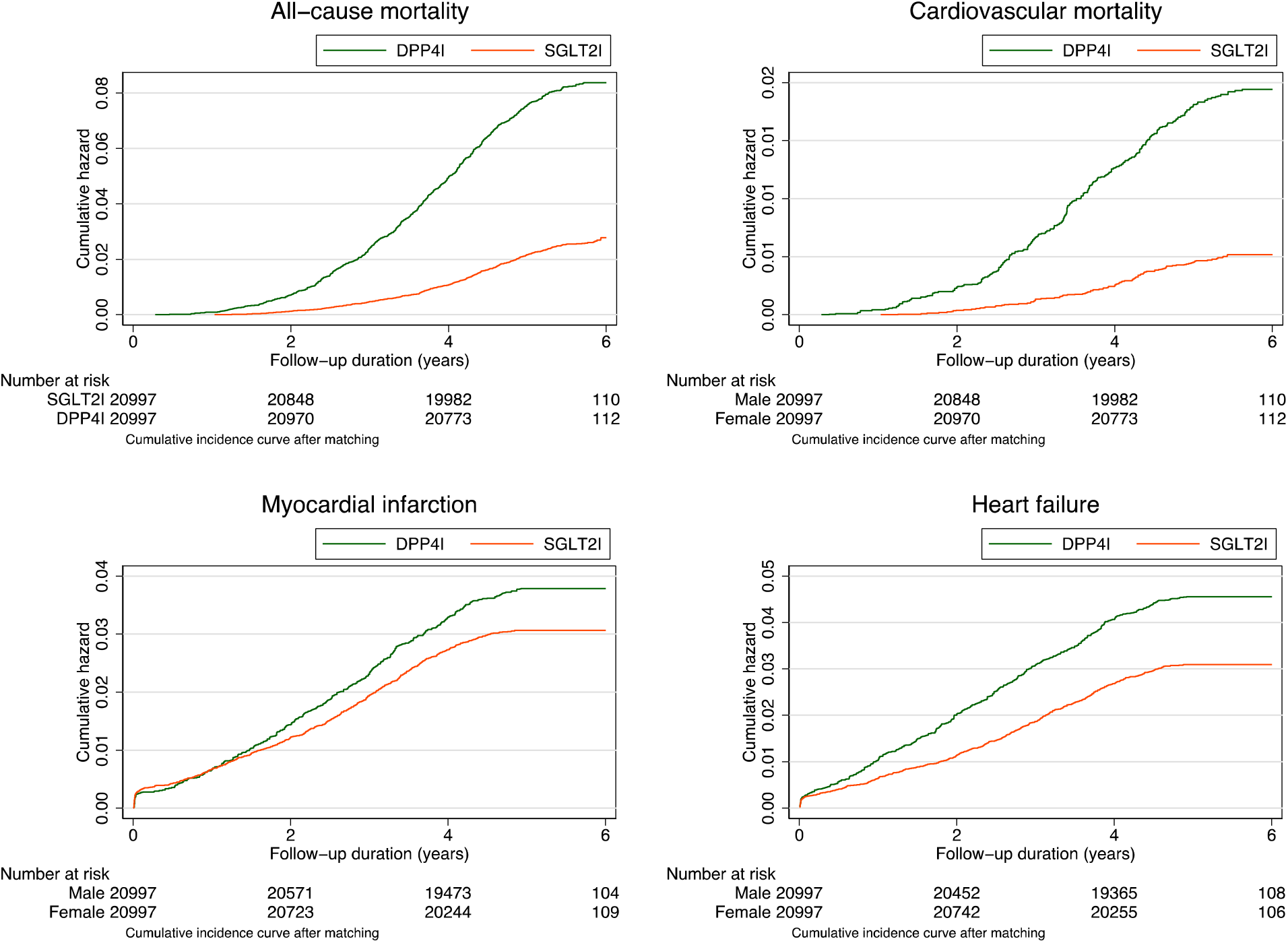
Cumulative incidence curves for heart failure, myocardial infarction, cardiovascular mortality, and all-cause mortality stratified by drug exposure of SGLT2I v.s. DPP4I in the matched cohort.

**Supplementary Figure 1** presented the propensity score matching comparisons for SGLT2I v.s. DPP4I use before and after 1:1 matching with nearest neighbor search strategy with a caliper of 0.1. In addition, we conducted bootstrapping procedures for propensity matching estimates and found that the estimations of bootstrapped standard error (replications=50) were less than 0.001. No significant confounding characteristics remained significant after propensity matching.

### Significant predictors of the study outcomes

**Table 2** presented the multivariable Cox analysis for HF, MI, cardiovascular mortality, and all-cause mortality in the matched cohort, adjusted for significant predictors identified for each outcome under univariable Cox regression **Supplementary tables 7** and **8** summarized the univariable Cox regression models to predict HF, MI, cardiovascular and all-cause mortality before and after 1:1 propensity score matching. After adjusting for significant demographics, past comorbidities, medication prescriptions and biochemical results, SGLT2I users have a significantly lower risk for MI (HR: 0.34, 95% CI: [0.28, 0.41], P < 0.0001), cardiovascular mortality (HR: 0.53, 95% CI: [0.38, 0.74], P = 0.0002) and all-cause mortality (HR: 0.21, 95% CI: [0.18, 0.25], P= 0.0001) under multivariable Cox regression. However, the risk for HF is comparable (HR: 0.87, 95% CI: [0.73, 1.04], P= 0.1343).

**Table 2.**
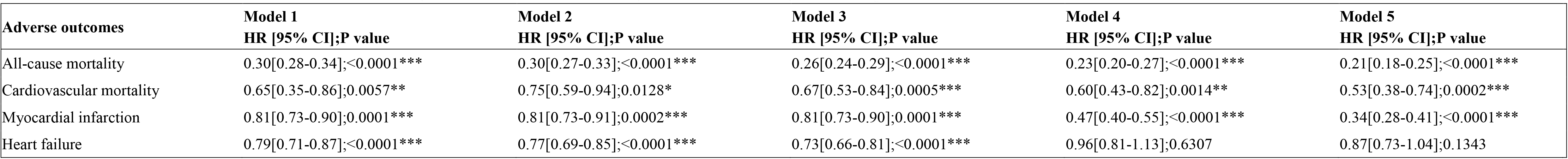
Multivariable Cox analysis for heart failure, myocardial infarction, cardiovascular mortality, and all-cause mortality in the matched cohort. * for p≤ 0.05, ** for p ≤ 0.01, *** for p ≤ 0.001; HR: hazard ratio; CI: confidence interval; SGLT2I: sodium glucose cotransporter-2 inhibitor; DPP4I: dipeptidyl peptidase-4 inhibitor. IR: incidence rate. Model 1 adjusted for significant demographics. Model 2 adjusted for significant demographics, and past comorbidities. Model 3 adjusted for significant demographics, past comorbidities, and non-SGLT2I/DPP4I medications. Model 4 adjusted for significant demographics, past comorbidities, non-SGLT2I/DPP4I medications, and subclinical biomarkers. Model 5 adjusted for significant demographics, past comorbidities, non-SGLT2I/DPP4I medications, subclinical biomarkers, HbA1c and glucose tests.

To evaluate the predictiveness of the models, a series of sensitivity analyses have been performed. **Table 3** shows that SGLT2I users are at a higher risk for all study outcomes, in comparison to DPP4I users, with a one-year lag time in the 1:1 ratio propensity score-matched cohort. The association of SGLT2I/ DPP4I use and the development of HF, MI, cardiovascular and all-cause mortality is preserved after the application of competing risk models (**Table 4**) and different propensity score matching approaches (**Table 5**).

**Table 3.** Sensitivity analysis 1: Hazard ratios for SGLT2I v.s. DPP4I exposure effects for heart failure, myocardial infarction, cardiovascular mortality, and all-cause mortality in the matched cohort, with one-year lag time. for p≤ 0.05, ** for p ≤ 0.01, *** for p ≤ 0.001; SGLT2I: Sodium-glucose cotransporter-2 inhibitors; DPP4I: Dipeptidyl peptidase-4 inhibitors; HR: hazard ratio; CI: confidence interval.

| Adverse outcomes | SGLT2I v.s. DPP4I<br>HR [95% CI];P value |
| --- | --- |
| All-cause mortality | 0.25[0.23-0.29];<0.0001 *** |
| Cardiovascular mortality | 0.26[0.20-0.34];<0.0001 *** |
| Myocardial infarction | 0.46[0.40-0.52];<0.0001 *** |
| Heart failure | 0.40[0.35-0.46];<0.0001 *** |

**Table 4.** Sensitivity analysis 2: Associations of SGLT2I v.s. DPP4I on heart failure, myocardial infarction, cardiovascular mortality, and all-cause mortality after propensity score matching (1:1) with competing risk models. for p≤ 0.05, ** for p ≤ 0.01, *** for p ≤ 0.001; SGLT2I: Sodium-glucose cotransporter-2 inhibitors; DPP4I: Dipeptidyl peptidase-4 inhibitors; HR: hazard ratio; CI: confidence interval.

| Models | Outcomes | SGLT2I v.s. DPP4I<br>HR [95% CI];P value |
| --- | --- | --- |
| *Cause-specific hazard models* |  |  |
|  | All-cause mortality | 0.24[0.21-0.27];<0.0001 *** |
|  | Cardiovascular mortality | 0.30[0.22-0.40];<0.0001 *** |
|  | Myocardial infarction | 0.49[0.42-0.57];<0.0001 *** |
|  | Heart failure | 0.45[0.39-0.53];<0.0001 *** |
| *Subdistribution hazard models* |  |  |
|  | All-cause mortality | 0.21[0.19-0.24];<0.0001 *** |
|  | Cardiovascular mortality | 0.25[0.19-0.32];<0.0001 *** |
|  | Myocardial infarction | 0.61[0.55-0.69];<0.0001 *** |
|  | Heart failure | 0.56[0.50-0.62];<0.0001 *** |

**Table 5.** Sensitivity analysis 3: Hazard ratios of SGLT2I v.s. DPP4I treatment for incident heart failure, myocardial infarction, cardiovascular mortality, and all-cause mortality in the matched cohort using different propensity matching approaches (1:1). for p≤ 0.05, ** for p ≤ 0.01, *** for p ≤ 0.001; SGLT2I: Sodium-glucose cotransporter-2 inhibitors; DPP4I: Dipeptidyl peptidase-4 inhibitors; HR: hazard ratio; CI: confidence interval; PS: propensity score; IPTW: inverse probability of treatment weighting, SIPTW: stable inverse probability of treatment weighting.

| Outcomes | PS stratification<br>HR [95% CI];P value | PS with IPTW<br>HR [95% CI];P value | PS with SIPTW<br>HR [95% CI];P value |
| --- | --- | --- | --- |
| All-cause mortality | 0.26[0.11-0.36];<0.0001 *** | 0.24[0.18-0.34];<0.0001 *** | 0.27[0.18-0.36];<0.0001 *** |
| Cardiovascular mortality | 0.31[0.28-0.51];<0.0001 *** | 0.35[0.24-0.45];<0.0001 *** | 0.32[0.24-0.45];<0.0001 *** |
| Myocardial infarction | 0.51[0.45-0.69];<0.0001 *** | 0.54[0.42-0.65];<0.0001 *** | 0.60[0.32-0.65];<0.0001 *** |
| Heart failure | 0.59[0.51-0.67];<0.0001 *** | 0.63[0.31-0.69];<0.0001 *** | 0.62[0.30-0.71];<0.0001 *** |

The descriptive statistics and HR of individual SGLT2Isfor the study outcomes in the matched cohort is summarized in **Table 6**. Dapagliflozin (n= 12416) is the most commonly prescribed SGLT2I, and it is protective against all study outcomes (P < 0.0001). Similarly, ertugliflozin (n= 2394) is also protective against all outcomes (P < 0.05). Canagliflozin (n= 4836) users have a comparable risk for HF in comparison to DPP4I users (HR: 0.93, 95% CI: [0.79, 1.08], P= 0.3249) but are protected against the remaining outcomes (P < 0.0001). Empagliflozin (n= 4464) is only protective against all-cause (HR: 0.28, 95% CI: [0.23, 0.35], P < 0.0001) and cardiovascular mortality (HR: 0.22, 95% CI: [0.13, 0.37], P < 0.0001).

**Table 6.** Descriptive statistics and hazard ratios of individual SGLT2I drugs for new onset heart failure, new onset myocardial infarction, cardiovascular mortality, and all-cause mortality in the matched cohort. for p≤ 0.05, ** for p ≤ 0.01, *** for p ≤ 0.001; SD: Standard deviation; HR: hazard ratio, CI: confidence interval, SGLT2I: sodium glucose cotransporter 2 inhibitors, DPP4I: dipeptidyl peptidase-4 inhibitors.

| SGLT2I drugs | All SGLT2I users<br>(N= 20997)<br>N or Count(%) | All-cause mortality<br>(N=2674) |  | Cardiovascular mortality<br>(N=538) |  | New onset heart failure<br>(N=1809) |  | New onset myocardial infarction<br>(N=1649) |  |
| --- | --- | --- | --- | --- | --- | --- | --- | --- | --- |
|  |  | N or<br>Count(%) | HR [95% CI];P value | N or<br>Count(%) | HR [95% CI];P value | N or<br>Count(%) | HR [95% CI];P value | N or<br>Count(%) | HR [95% CI];P value |
| Dapagliflozin | 12416(59.13%) | 328(12.26%) | 0.32[0.29-0.36];<0.0001*** | 68(12.63%) | 0.33[0.26-0.42];<0.0001*** | 333(18.40%) | 0.52[0.46-0.59];<0.0001*** | 311(18.85%) | 0.54[0.48-0.61];<0.0001*** |
| Empagliflozin | 4464(21.26%) | 90(3.36%) | 0.28[0.23-0.35];<0.0001*** | 14(2.60%) | 0.22[0.13-0.37];<0.0001*** | 178(9.83%) | 0.90[0.77-1.05];0.1872 | 170(10.30%) | 0.95[0.81-1.11];0.5363 |
| Canagliflozin | 4836(23.03%) | 110(4.11%) | 0.32[0.26-0.39];<0.0001*** | 19(3.53%) | 0.27[0.17-0.43];<0.0001*** | 158(8.73%) | 0.72[0.61-0.85];0.0001*** | 180(10.91%) | 0.93[0.79-1.08];0.3249 |
| Ertugliflozin | 2394(11.40%) | 62(2.31%) | 0.38[0.30-0.49];<0.0001*** | 13(2.41%) | 0.40[0.23-0.69];0.0010** | 65(3.59%) | 0.60[0.47-0.77];0.0001*** | 75(4.54%) | 0.78[0.62-0.98];0.0314* |

### Stratification age and gender analysis

**Supplementary Table 4** illustrated the baseline and clinical characteristics of patients stratified by age and gender before and after propensity score matching (1:1). **Figure 3** presented the cumulative incidence curves for HF, MI, cardiovascular mortality, and all-cause mortality stratified by combinations of baseline age and drug exposure of SGLT2I/ DPP4I in the matched cohort. The risk for the study outcomes is higher amongst patients older than age 65 regardless of SGLT2I/ DPP4I use, and within the two age groups DPP4I users have a higher risk of adverse cardiovascular outcomes than SGLT2I users.

**Figure 3.**
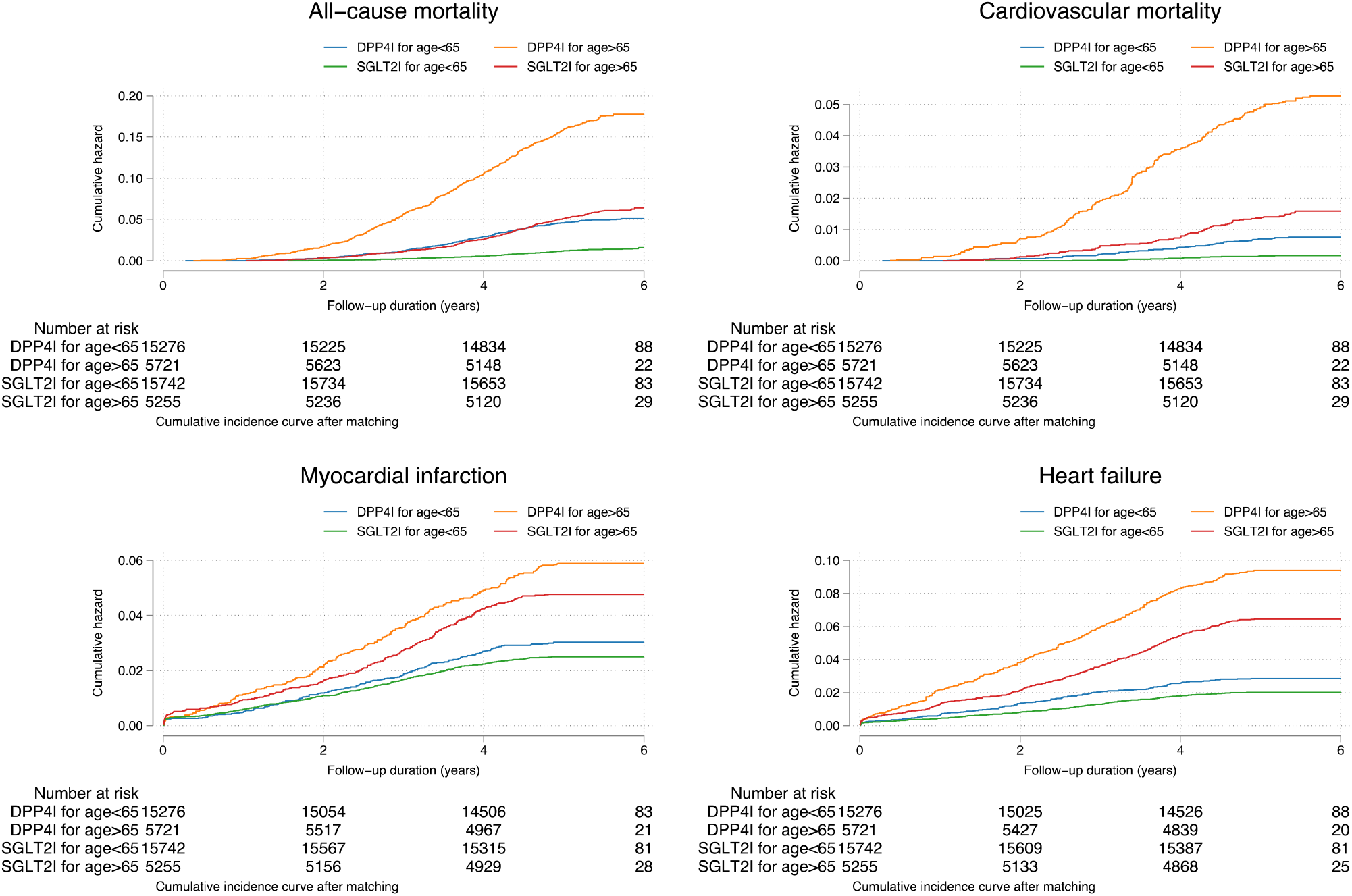
Cumulative incidence curves for heart failure, myocardial infarction, cardiovascular mortality, and all-cause mortality stratified by combinations of drug exposure of SGLT2I v.s. DPP4I and admission age in the matched cohort.

Figure 4 showed the cumulative incidence curves for HF, MI, cardiovascular mortality, and all-cause mortality stratified by combinations of gender and SGLT2I/ DPP4I use in the matched cohort. DPP4I users, regardless of gender, have a higher risk for mortality and HF than SGLT2I users, with male patients having a consistently higher risk than their female counterparts. However, in terms of the risk of MI, male SGLT2I users have a higher than female DPP4I users, suggesting that the cardioprotective effect of SGLT2I remains insufficient to compensate for the elevated MI risk of males.

**Figure 4.**
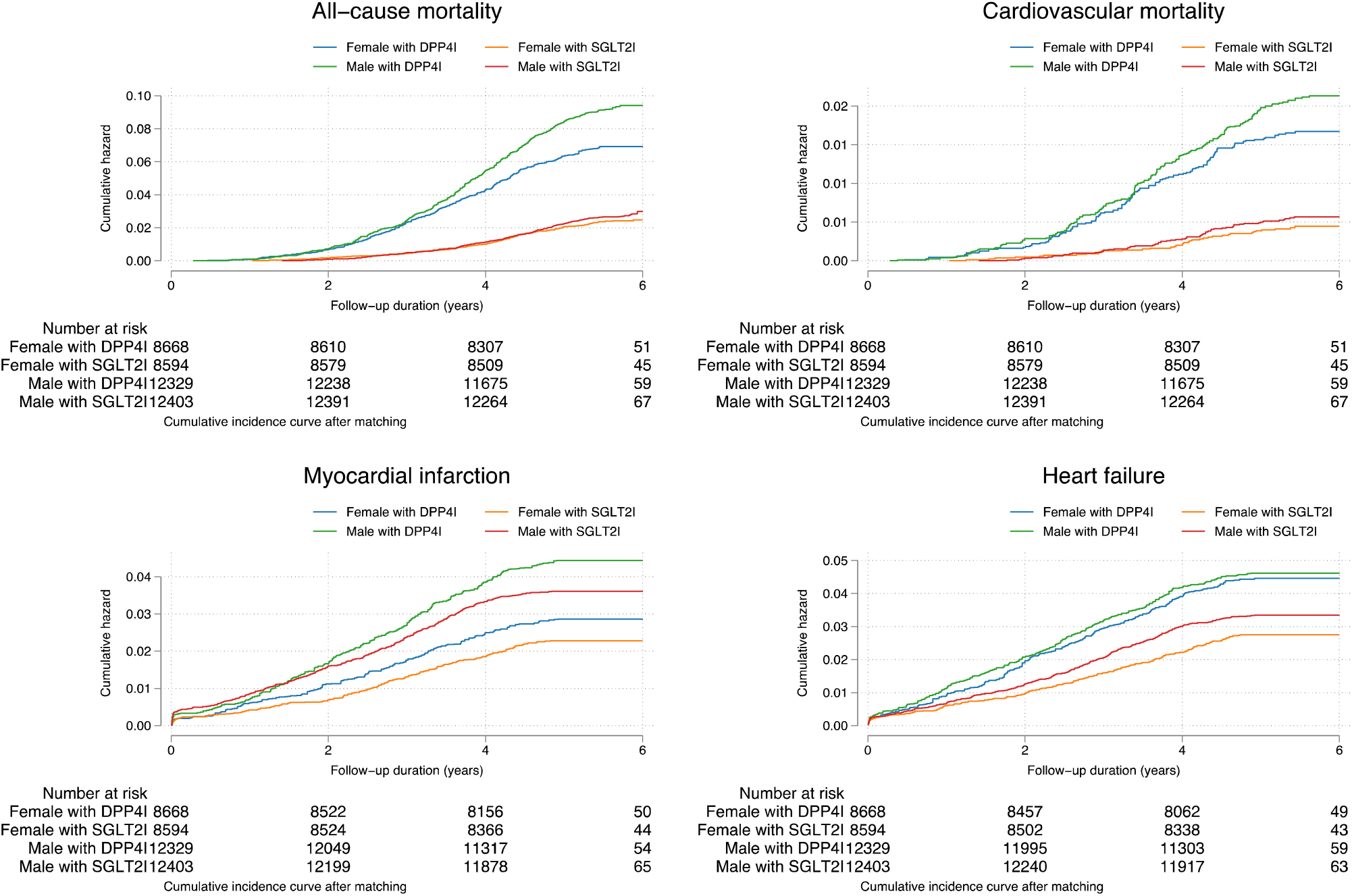
Cumulative incidence curves for heart failure, myocardial infarction, cardiovascular mortality, and all-cause mortality stratified by combinations of drug exposure of SGLT2I v.s. DPP4I and gender in the matched cohort.

## Discussion

The main findings of this study are that: 1) using DPP4I as a reference, SGLT2I use was associated with a lower risk of all-cause mortality, cardiovascular mortality, new onset of HF and MI; 2) the cardioprotective effect of SGLT2I is insufficient to compensate for the elevated MI risk of males; 3) the protective effects of empagliflozin against MI, which was only reported in pre-clinical studies in the past, is for the first time in humans at a population level amongst patients of different comorbidities. (22)

Findings from the present study are largely consistent with existing studies. A network meta-analysis of 236 trials has reported the superior cardiovascular protective effects of SGLT2I against DPP4I when users of either medication are compared against the control group. However, the control groups were not matched and no direct comparison was made. (14) A recent study evaluating the cardiovascular effects of SGLT2I and DPP4I amongst cardiorenal disease-free diabetic patients shows that SGLTI users have a lower risk of HF. (17) However, this study found the effect of SGLT2I on the prevention of acute MI to be neutral, which may be explained by the inherent difference between patients with renal failure and the general population. With a structured follow-up and close monitoring, patients with renal failure would have their cardiovascular risk factors optimized as a part of their disease management. Moreover, recent meta-analyses have reported the benefits of SGLT2I in preventing cardiac remodelling in heart failure patients regardless of glycemic status (23) and reduce major clinical events in patients with established heart failure (24), with a neutral effect on arrhythmic outcomes (25). Furthermore, a meta-analysis including more than 34 000 patients found that the protective effect of SGLT2I on major cardiovascular adverse events of atherosclerotic origin is limited to patients with established atherosclerotic disease. (26) The difference in the proportion of patients with established atherosclerosis may explain the different effects of SGLT2I on MI observed. The present study demonstrates that the cardiovascular beneficent effects of SGLT2I persist in diabetic patients with pre-existing cardiovascular impairment.

There are several hypotheses for the mechanisms underlying the cardiovascular-protective effects of SGLT2I. First of all, the modulatory effect of SGLT2I on the proximal tubules results in glucosuria and natriuresis, thus lowering the preload and the resulting stress on the ventricles. (27) It is speculated that SGLT2I has a unique effect of selectively contracting interstitial fluid specifically, without affecting the intravascular volume, thus particularly useful in the prevention of HF. (28) The hypothesis is supported by studies comparing the vascular effects of dapagliflozin and bumetanide, where dapagliflozin has been shown to have little effect on the intravascular volume. (29, 30)

Moreover, inhibition of the sodium-hydrogen ion exchanger in the myocardium, which is activated under HF to increase intracytoplasmic sodium and calcium level, was also hypothesized to be a part of the underlying mechanism. (31, 32) However, since SGLT2 receptors are absent in the heart, the exact inhibitory mechanism remains unclear. Other hypotheses on the anti-fibrosis and adipokine-reducing effects, which are effective against both HF and MI, suggest that the cardiovascular-protective effects of SGLT2I may involve multiple biochemical pathways and thus protect against different cardiovascular diseases. (28, 33)

The multiple processes involved in the cardiovascular-protective effect of SGLTI may also explain its superior outcome against DPP4I. Whilst previous studies reported the benefits of SGLT2I on cardiovascular health are mainly attributed to its protection against HF, a recent territory-wide study has shown that SGLT2I users have a lower incidence of new-onset atrial fibrillation than DPP4I users, which supports the lower cardiovascular and all-cause mortality reported in the present study. (34) This may be attributed to the anti-fibrotic effects of SGLT2I, since atrial remodeling and fibrosis are common pathogenic pathways of atrial fibrillation. (35) The favorable pleiotropic effects of SGLT2I may also improve the patients’ cardiometabolic risk, thus further lowering their MI and cardiovascular mortality risk. (36) It should be noted that randomized controlled trials have reported that saxagliptin increases the hospitalization rate for HF, despite having a neutral effect on the occurrence of major cardiovascular adverse effects. (37, 38) Since the present study focuses on the incident occurrence of HF and MI, patients on saxagliptin were kept in the study. Among the 69521 patients with type-2 diabetes, there were in total 353 patients who used saxagliptin use with a quite low incidence rate as 0.51%.

### Limitations

There are several limitations to the present study. Firstly, inherent information bias with a risk of under-coding and coding errors should be noted, given its observational and retrospective nature. However, the difference in patient characteristics, past comorbidities and other medication usages between SGLT4I/DPP4I users and controls were addressed through matching using propensity scores, although residual bias may remain. There are also patients with missing data for the laboratory parameters since not all blood tests were routinely performed for all. Moreover, we were unable to access important lifestyle predictors for cardiovascular adverse events, such as body mass index, smoking, and alcoholism. Thirdly, coding for clinical diagnoses of HF was used but echocardiographic data are not coded in the administrative database and therefore different types of HF based on ejection fraction could not be examined. Finally, DPP4I use is associated with an increased risk of HF compared to placebo, and therefore this study could not distinguish between whether gliptins cause HF and whether SGLT2Is reduce HF.

## Conclusions

In comparison to DPP4I, SGLT2I use is associated with lower risks of all-cause mortality, cardiovascular mortality, new onset HF and MI after adjustments for confounders. The prescription of SGLT2I is preferred after taking into consideration the patient’s cardiovascular and metabolic risk profile in addition to drug-drug interactions.

## Data Availability

All data produced in the present study are available upon reasonable request to the authors

## Author contributions

Jiandong Zhou, Sharen Lee: conception of study and literature search, preparation of figures, study design, data collection, data contribution, statistical analysis, data interpretation, manuscript drafting, and critical revision of the manuscript.

Keith Sai Kit Leung, Abraham Ka Chung Wai, Tong Liu, Ying Liu, Dong Chang, Wing Tak Wong, Ian Chi Kei Wong, Bernard Man Yung Cheung

Qingpeng Zhang, Gary Tse: conception of study and literature search, study design, data collection, data analysis, data contribution, manuscript drafting, and critical revision of manuscript, study supervision.

## Conflicts of Interest

None.

## Acknowledgements

None.

## Notes

### Competing Interest Statement

The authors have declared no competing interest.

### Funding Statement

This study did not receive any funding

### Author Declarations

This study was approved by the Institutional Review Board of the University of Hong Kong/Hospital Authority Hong Kong West Cluster and from The Joint Chinese University of Hong Kong-New Territories East Cluster Clinical Research Ethics Committee.

## References

1. Zheng Y, Ley SH, Hu FB. Global aetiology and epidemiology of type 2 diabetes mellitus and its complications. Nat Rev Endocrinol. 2018;14(2):88–98.

2. Curtis HJ, Dennis JM, Shields BM, Walker AJ, Bacon S, Hattersley AT, et al. Time trends and geographical variation in prescribing of drugs for diabetes in England from 1998 to 2017. Diabetes Obes Metab. 2018;20(9):2159–68.

3. Cho YK, Kang YM, Lee SE, Lee J, Park JY, Lee WJ, et al. Efficacy and safety of combination therapy with SGLT2 and DPP4 inhibitors in the treatment of type 2 diabetes: A systematic review and meta-analysis. Diabetes Metab. 2018;44(5):393–401.

4. Fei Y, Tsoi M-F, Cheung BMY. Cardiovascular outcomes in trials of new antidiabetic drug classes: a network meta-analysis. Cardiovascular Diabetology. 2019;18(1):112.

5. Zinman B, Wanner C, Lachin JM, Fitchett D, Bluhmki E, Hantel S, et al. Empagliflozin, Cardiovascular Outcomes, and Mortality in Type 2 Diabetes. N Engl J Med. 2015;373(22):2117–28.

6. Verma S, Mazer CD, Fitchett D, Inzucchi SE, Pfarr E, George JT, et al. Empagliflozin reduces cardiovascular events, mortality and renal events in participants with type 2 diabetes after coronary artery bypass graft surgery: subanalysis of the EMPA-REG OUTCOME(R) randomised trial. Diabetologia. 2018;61(8):1712–23.

7. Fitchett D, Butler J, van de Borne P, Zinman B, Lachin JM, Wanner C, et al. Effects of empagliflozin on risk for cardiovascular death and heart failure hospitalization across the spectrum of heart failure risk in the EMPA-REG OUTCOME(R) trial. Eur Heart J. 2018;39(5):363–70.

8. Radholm K, Figtree G, Perkovic V, Solomon SD, Mahaffey KW, de Zeeuw D, et al. Canagliflozin and Heart Failure in Type 2 Diabetes Mellitus: Results From the CANVAS Program. Circulation. 2018;138(5):458–68.

9. Ou HT, Chang KC, Li CY, Wu JS. Risks of cardiovascular diseases associated with dipeptidyl peptidase-4 inhibitors and other antidiabetic drugs in patients with type 2 diabetes: a nation-wide longitudinal study. Cardiovasc Diabetol. 2016;15:41.

10. Scirica BM, Braunwald E, Raz I, Cavender MA, Morrow DA, Jarolim P, et al. Heart failure, saxagliptin, and diabetes mellitus: observations from the SAVOR-TIMI 53 randomized trial. Circulation. 2014;130(18):1579–88.

11. Cha SA, Park YM, Yun JS, Lim TS, Song KH, Yoo KD, et al. A comparison of effects of DPP-4 inhibitor and SGLT2 inhibitor on lipid profile in patients with type 2 diabetes. Lipids Health Dis. 2017;16(1):58.

12. Gautam S, Agiro A, Barron J, Power T, Weisman H, White J. Heart failure hospitalization risk associated with use of two classes of oral antidiabetic medications: an observational, real-world analysis. Cardiovasc Diabetol. 2017;16(1):93.

13. Fuchigami A, Shigiyama F, Kitazawa T, Okada Y, Ichijo T, Higa M, et al. Efficacy of dapagliflozin versus sitagliptin on cardiometabolic risk factors in Japanese patients with type 2 diabetes: a prospective, randomized study (DIVERSITY-CVR). Cardiovasc Diabetol. 2020;19(1):1.

14. Zheng SL, Roddick AJ, Aghar-Jaffar R, Shun-Shin MJ, Francis D, Oliver N, et al. Association Between Use of Sodium-Glucose Cotransporter 2 Inhibitors, Glucagon-like Peptide 1 Agonists, and Dipeptidyl Peptidase 4 Inhibitors With All-Cause Mortality in Patients With Type 2 Diabetes: A Systematic Review and Meta-analysis. JAMA. 2018;319(15):1580–91.

15. Shao S-C, Chang K-C, Lin S-J, Chien R-N, Hung M-J, Chan Y-Y, et al. Favorable pleiotropic effects of sodium glucose cotransporter 2 inhibitors: head-to-head comparisons with dipeptidyl peptidase-4 inhibitors in type 2 diabetes patients. Cardiovascular Diabetology. 2020;19(1):17.

16. Ling AW-C, Chan C-C, Chen S-W, Kao Y-W, Huang C-Y, Chan Y-H, et al. The risk of new-onset atrial fibrillation in patients with type 2 diabetes mellitus treated with sodium glucose cotransporter 2 inhibitors versus dipeptidyl peptidase-4 inhibitors. Cardiovascular Diabetology. 2020;19(1):188.

17. Birkeland KI, Bodegard J, Banerjee A, Kim DJ, Norhammar A, Eriksson JW, et al. Lower cardiorenal risk with sodium-glucose cotransporter-2 inhibitors versus dipeptidyl peptidase-4 inhibitors in patients with type 2 diabetes without cardiovascular and renal diseases: A large multinational observational study. Diabetes, Obesity and Metabolism. 2021;23(1):75–85.

18. Zhou J, Wang X, Lee S, Wu WKK, Cheung BMY, Zhang Q, et al. Proton pump inhibitor or famotidine use and severe COVID-19 disease: a propensity score-matched territory-wide study. Gut. 2020.

19. Ju C, Lai RWC, Li KHC, Hung JKF, Lai JCL, Ho J, et al. Comparative cardiovascular risk in users versus non-users of xanthine oxidase inhibitors and febuxostat versus allopurinol users. Rheumatology (Oxford). 2020;59(9):2340–9.

20. Lee S, Liu T, Zhou J, Zhang Q, Wong WT, Tse G. Predictions of diabetes complications and mortality using hba1c variability: a 10-year observational cohort study. Acta Diabetol. 2020.

21. Mui JV, Zhou J, Lee S, Leung KSK, Lee TTL, Chou OHI, et al. Sodium-Glucose Cotransporter 2 (SGLT2) Inhibitors vs. Dipeptidyl Peptidase-4 (DPP4) Inhibitors for New-Onset Dementia: A Propensity Score-Matched Population-Based Study With Competing Risk Analysis. Front Cardiovasc Med. 2021;8:747620.

22. Oshima H, Miki T, Kuno A, Mizuno M, Sato T, Tanno M, et al. Empagliflozin, an SGLT2 Inhibitor, Reduced the Mortality Rate after Acute Myocardial Infarction with Modification of Cardiac Metabolomes and Antioxidants in Diabetic Rats. J Pharmacol Exp Ther. 2019;368(3):524–34.

23. Zhang N, Wang Y, Tse G, Korantzopoulos P, Letsas KP, Zhang Q, et al. Effect of sodium-glucose cotransporter-2 inhibitors on cardiac remodelling: a systematic review and meta-analysis. Eur J Prev Cardiol. 2021.

24. Bazoukis G, Papadatos SS, Thomopoulos C, Tse G, Cheilidis S, Tsioufis K, et al. Impact of SGLT2 inhibitors on major clinical events and safety outcomes in heart failure patients: a meta-analysis of randomized clinical trials. J Geriatr Cardiol. 2021;18(10):783–95.

25. Sfairopoulos D, Zhang N, Wang Y, Chen Z, Letsas KP, Tse G, et al. Association between sodium-glucose cotransporter-2 inhibitors and risk of sudden cardiac death or ventricular arrhythmias: a meta-analysis of randomized controlled trials. Europace. 2021.

26. Zelniker TA, Wiviott SD, Raz I, Im K, Goodrich EL, Bonaca MP, et al. SGLT2 inhibitors for primary and secondary prevention of cardiovascular and renal outcomes in type 2 diabetes: a systematic review and meta-analysis of cardiovascular outcome trials. Lancet. 2019;393(10166):31–9.

27. Sattar N, McLaren J, Kristensen SL, Preiss D, McMurray JJ. SGLT2 Inhibition and cardiovascular events: why did EMPA-REG Outcomes surprise and what were the likely mechanisms? Diabetologia. 2016;59(7):1333–9.

28. Verma S, McMurray JJV. SGLT2 inhibitors and mechanisms of cardiovascular benefit: a state-of-the-art review. Diabetologia. 2018;61(10):2108–17.

29. Wilcox CS, Shen W, Boulton DW, Leslie BR, Griffen SC. Interaction Between the Sodium-Glucose-Linked Transporter 2 Inhibitor Dapagliflozin and the Loop Diuretic Bumetanide in Normal Human Subjects. J Am Heart Assoc. 2018;7(4).

30. Hallow KM, Helmlinger G, Greasley PJ, McMurray JJV, Boulton DW. Why do SGLT2 inhibitors reduce heart failure hospitalization? A differential volume regulation hypothesis. Diabetes Obes Metab. 2018;20(3):479–87.

31. Packer M, Anker SD, Butler J, Filippatos G, Zannad F. Effects of Sodium-Glucose Cotransporter 2 Inhibitors for the Treatment of Patients With Heart Failure: Proposal of a Novel Mechanism of Action. JAMA Cardiol. 2017;2(9):1025–9.

32. Baartscheer A, Schumacher CA, Wust RC, Fiolet JW, Stienen GJ, Coronel R, et al. Empagliflozin decreases myocardial cytoplasmic Na(+) through inhibition of the cardiac Na(+)/H(+) exchanger in rats and rabbits. Diabetologia. 2017;60(3):568–73.

33. Shao Q, Meng L, Lee S, Tse G, Gong M, Zhang Z, et al. Empagliflozin, a sodium glucose co-transporter-2 inhibitor, alleviates atrial remodeling and improves mitochondrial function in high-fat diet/streptozotocin-induced diabetic rats. Cardiovasc Diabetol. 2019;18(1):165.

34. Ling AW, Chan CC, Chen SW, Kao YW, Huang CY, Chan YH, et al. The risk of new-onset atrial fibrillation in patients with type 2 diabetes mellitus treated with sodium glucose cotransporter 2 inhibitors versus dipeptidyl peptidase-4 inhibitors. Cardiovasc Diabetol. 2020;19(1):188.

35. Fedak PW, Verma S, Weisel RD, Li RK. Cardiac remodeling and failure From molecules to man (Part II). Cardiovasc Pathol. 2005;14(2):49–60.

36. Shao SC, Chang KC, Lin SJ, Chien RN, Hung MJ, Chan YY, et al. Favorable pleiotropic effects of sodium glucose cotransporter 2 inhibitors: head-to-head comparisons with dipeptidyl peptidase-4 inhibitors in type 2 diabetes patients. Cardiovasc Diabetol. 2020;19(1):17.

37. Scirica BM, Bhatt DL, Braunwald E, Steg PG, Davidson J, Hirshberg B, et al. Saxagliptin and cardiovascular outcomes in patients with type 2 diabetes mellitus. N Engl J Med. 2013;369(14):1317–26.

38. Udell JA, Bhatt DL, Braunwald E, Cavender MA, Mosenzon O, Steg PG, et al. Saxagliptin and cardiovascular outcomes in patients with type 2 diabetes and moderate or severe renal impairment: observations from the SAVOR-TIMI 53 Trial. Diabetes Care. 2015;38(4):696–705.

