## Supplementary Appendix for "Incident heart failure and myocardial infarction in sodium-glucose cotransporter 2 versus dipeptidyl peptidase-4 inhibitor users"

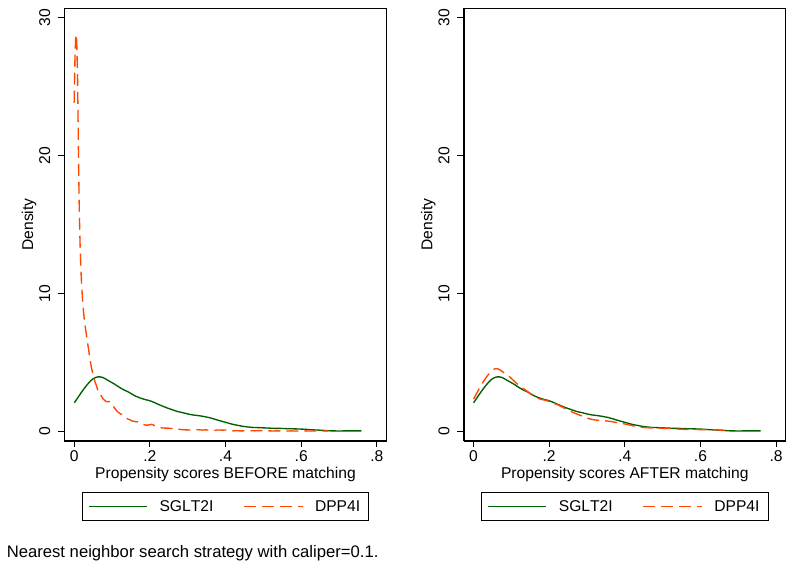

**Supplementary Figure 1. Propensity score matching comparisons for SGLT2I v.s. DPP4I before and after 1:1 matching with nearest neighbor search strategy using a caliper of 0.1**

**Supplementary Table 1. ICD-9 codes for comorbidities and ICD10 codes for outcomes**

Diabetes mellitus 250	250.01	250.02	250.03	250.1	250.11	250.12	250.13	250.2	250.21	250.22	250.23	250.3	250.31	250.32	250.33	250.4	250.41	250.42	250.43	250.5	250.51	250.52	250.53	250.6	250.61	250.62	250.63	250.7	250.71	250.72	250.73	250.8	250.81	250.82	250.83	250.9	250.91	250.92	250.93

Renal diseases	582	582	582.1	582.2	582.4	582.8	582.81	582.89	582.9	583	583	583.1	583.2	583.4	583.6	583.7	585	585.1	585.2	585.3	585.4	585.5	585.6	585.9	586	588	588	588.1	588.8	588.81	588.89	588.9

Acute myocardial infarction 410	410.01	410.02	410.1	410.11	410.12	410.2	410.21	410.22	410.3	410.31	410.32	410.4	410.41	410.42	410.5	410.51	410.52	410.6	410.61	410.62	410.7	410.71	410.72	410.8	410.81	410.82	410.9	410.91	410.92

Hypertension	401	401.1	401.9	402	402.01	402.1	402.11	402.9	402.91	403	403.01	403.1	403.11	403.9	403.91	404	404.01	404.02	404.03	404.1	404.11	404.12	404.13	404.9	404.91	404.92	404.93	405	405.01	405.09	405.1	405.11	405.19	405.9	405.91	405.99	437.2

Heart failure	428	428	428.1	428.2	428.2	428.21	428.22	428.23	428.3	428.3	428.31	428.32	428.33	428.4	428.4	428.41	428.42	428.43	428.9	398.91	402.01	402.11	402.91	404.01	404.03	404.11	404.13	404.91	404.93

Atrial fibrillation	427.31	429.4

Liver diseases	456	456.1	456.2	572.2	572.3	572.4	572.8	571.4	571.5	571.6

Chronic obstructive pulmonary disease	490	491	492	493	494	495	496	491.1	491.2	491.21	491.22	491.8	491.9	492.8	493.01	493.02	493.1	493.11	493.12	493.2	493.21	493.22	493.8	493.81	493.82	493.9	493.91	493.92	494.1	495.1	495.2	495.3	495.4	495.5	495.6	495.7	495.8	495.9

PVD	250.7	443.9	443	443.1	443.2	443.21	443.22	443.23	443.24	443.29	443.8	443.81	443.82	443.89	441	443.9	785.4	V43.4

Stroke/transient ischemic attack	435	435.1	435.2	435.3	435.8	435.9	433.81	433.91	434	436	437	437.1	433.31	433.01	434.01	434.1	434.11	434.9	434.91	437.2	437.3	437.4	437.5	437.6	437.7	437.8	437.9	430	431	432	432.1	432.9

Ischemic heart disease	410.01	410.02	410.1	410.11	410.12	410.2	410.21	410.22	410.3	410.31	410.32	410.4	410.41	410.42	410.5	410.51	410.52	410.6	410.61	410.62	410.7	410.71	410.72	410.8	410.81	410.82	410.9	410.91	410.92	411	411.1	411.8	411.81	411.89	413	413.1	413.9	414	414.01	414.02	414.03	414.04	414.05	414.06	414.07	414.1	414.11	414.12	414.19	414.2	414.3	414.4	414.8	414.9	410	412

Cancer 140	140.1	140.3	140.4	140.5	140.6	140.8	140.9	141	141.1	141.2	141.3	141.4	141.5	141.6	141.8	141.9	142	142.1	142.2	142.8	142.9	143	143.1	143.8	143.9	144	144.1	144.8	144.9	145	145.1	145.2	145.3	145.4	145.5	145.6	145.8	145.9	146	146.1	146.2	146.3	146.4	146.5	146.6	146.7	146.8	146.9	147	147.1	147.2	147.3	147.8	147.9	148	148.1	148.2	148.3	148.8	148.9	149	149.1	149.8	149.9	150	150.1	150.2	150.3	150.4	150.5	150.8	150.9	151	151.1	151.2	151.3	151.4	151.5	151.6	151.8	151.9	152	152.1	152.2	152.3	152.8	152.9	153	153.1	153.2	153.3	153.4	153.5	153.6	153.7	153.8	153.9	154	154.1	154.2	154.3	154.8	155	155.1	155.2	156	156.1	156.2	156.8	156.9	157	157.1	157.2	157.3	157.4	157.8	157.9	158	158.8	158.9	159	159.1	159.8	159.9	160	160.1	160.2	160.3	160.4	160.5	160.8	160.9	161	161.1	161.2	161.3	161.8	161.9	162	162.2	162.3	162.4	162.5	162.8	162.9	163	163.1	163.8	163.9	164	164.1	164.2	164.3	164.8	164.9	165	165.8	165.9	170	170.1	170.2	170.3	170.4	170.5	170.6	170.7	170.8	170.9	171	171.2	171.3	171.4	171.5	171.6	171.7	171.8	171.9	172	172.1	172.2	172.3	172.4	172.5	172.6	172.7	172.8	172.9 173	173.01	173.02	173.09	173.1	173.11	173.12	173.19	173.2	173.21	173.22	173.29	173.3	173.31	173.32	173.39	173.4	173.41	173.42	173.49	173.5	173.51	173.52	173.59	173.6	173.61	173.62	173.69	173.7	173.71	173.72	173.79	173.8	173.81	173.82	173.89	173.9	173.91	173.92	173.99	174	174.1	174.2	174.3	174.4	174.5	174.6	174.8	174.9	175	175.9	176	176.1	176.2	176.3	176.4	176.5	176.8	176.9	179	180	180.1	180.8	180.9	181	182	182.1	182.8	183	183.2	183.3	183.4	183.5	183.8	183.9	184	184.1	184.2	184.3	184.4	184.8	184.9	185	186	186.9	187	187.1	187.2	187.3	187.4	187.5	187.6	187.7	187.8	187.9	188	188.1	188.2	188.3	188.4	188.5	188.6	188.7	188.8	188.9	189	189.1	189.2	189.3	189.4	189.8	189.9	190	190.1	190.2	190.3	190.4	190.5	190.6	190.7	190.8	190.9	191	191.1	191.2	191.3	191.4	191.5	191.6	191.7	191.8	191.9	192	192.1	192.2	192.3	192.8	192.9	193	194	194.1	194.3	194.4	194.5	194.6	194.8	194.9	195	195.1	195.2	195.3	195.4	195.5	195.8	200	200.01	200.02	200.03	200.04	200.05	200.06	200.07	200.08	200.1	200.11	200.12	200.13	200.14	200.15	200.16	200.17	200.18	200.2	200.21	200.22	200.23	200.24	200.25	200.26	200.27	200.28	200.3	200.31	200.32	200.33	200.34	200.35	200.36	200.37	200.38	200.4	200.41	200.42	200.43	200.44	200.45	200.46	200.47	200.48	200.5	200.51	200.52	200.53	200.54	200.55	200.56	200.57	200.58	200.6	200.61	200.62	200.63	200.64	200.65	200.66	200.67	200.68	200.7	200.71	200.72	200.73	200.74	200.75	200.76	200.77	200.78	200.8	200.81	200.82	200.83	200.84	200.85	200.86	200.87	200.88	201	201.01	201.02	201.03	201.04	201.05	201.06	201.07	201.08	201.1	201.11	201.12	201.13	201.14	201.15	201.16	201.17	201.18	201.2	201.21	201.22	201.23	201.24	201.25	201.26	201.27	201.28	201.4	201.41	201.42	201.43	201.44	201.45	201.46	201.47	201.48	201.5	201.51	201.52	201.53	201.54	201.55	201.56	201.57	201.58	201.6	201.61	201.62	201.63	201.64	201.65	201.66	201.67	201.68	201.7	201.71	201.72	201.73	201.74	201.75	201.76	201.77	201.78	201.9	201.91	201.92	201.93	201.94	201.95	201.96	201.97	201.98	202	202.01	202.02	202.03	202.04	202.05	202.06	202.07	202.08	202.1	202.11	202.12	202.13	202.14	202.15	202.16	202.17	202.18	202.2	202.21	202.22	202.23	202.24	202.25	202.26	202.27	202.28	202.3	202.31	202.32	202.33	202.34	202.35	202.36	202.37	202.38	202.4	202.41	202.42	202.43	202.44	202.45	202.46	202.47	202.48	202.5	202.51	202.52	202.53	202.54	202.55	202.56	202.57	202.58	202.6	202.61	202.62	202.63	202.64	202.65	202.66	202.67	202.68	202.7	202.71	202.72	202.73	202.74	202.75	202.76	202.77	202.78	202.8	202.81	202.82	202.83	202.84	202.85	202.86	202.87	202.88	202.9	202.91	202.92	202.93	202.94	202.95	202.96	202.97	202.98	203	203.01	203.02	203.1	203.11	203.12	203.8	203.81	203.82	204	204.01	204.02	204.1	204.11	204.12	204.2	204.21	204.22	204.8	204.81	204.82	204.9	204.91	204.92	205	205.01	205.02	205.1	205.11	205.12	205.2	205.21	205.22	205.3	205.31	205.32	205.8	205.81	205.82	205.9	205.91	205.92	206	206.01	206.02	206.1	206.11	206.12	206.2	206.21	206.22	206.8	206.81	206.82	206.9	206.91	206.92	207	207.01	207.02	207.1	207.11	207.12	207.2	207.21	207.22	207.8	207.81	207.82	208	208.01	208.02	208.1	208.11	208.12	208.2	208.21	208.22	208.8	208.81	208.82	208.9	208.91	208.92 196	196.1	196.2	196.3	196.5	196.6	196.8	196.9	197	197.1	197.2	197.3	197.4	197.5	197.6	197.7	197.8	198	198.1	198.2	198.3	198.4	198.5	198.6	198.7	198.8	198.81	198.82	198.89	199	199.1

Obesity	278.01	278	278

Hypertension	401	401.1	401.9	402	402.01	402.1	402.11	402.9	402.91	403	403.01	403.1	403.11	403.9	403.91	404	404.01	404.02	404.03	404.1	404.11	404.12	404.13	404.9	404.91	404.92	404.93	405	405.01	405.09	405.1	405.11	405.19	405.9	405.91	405.99	437.2

Immune mediated enterocolitis	558	558.1	558.2	558.3	558.4	558.41	558.42	558.9

Anemia	280	280.1	280.8	280.9	281	281.1	281.2	281.3	281.4	281.8	281.9	282.2	282.3	282.8	282.9	283	283.1	283.11	283.19	283.2	283.9	284	284.01	284.09	284.1	284.11	284.12	284.19	284.81	284.9	285	285.1	285.2	285.21	285.22	285.29	285.3	285.8	285.9

Overweight	278	278	278	278.01	278.02	278.03	278.1	278.2	278.3	278.4	278.8

Gout	274	274.01	274.02	274.03	274.1	274.11	274.19	274.8	274.81	274.82	274.89	274.9

Cardiovascular mortality I00-I09, I11, I13, I20-I51

**Supplementary Table 2. Calculations for measures of variability**

| **Variability measure** | **Definition** |
| --- | --- |
| Standard deviation | $\sqrt{\frac{1}{Number of measurements}\sum_{i=1}^{Number of measurements} {({test}_{i}-individual mean)}^{2}}$ |
| SD/initial | $\frac{SD}{individual initial value}$ |
| Coefficient of variation | $\frac{SD}{individual mean}$ |
| Variability independent of mean | $\frac{SD}{{individual mean}^{\frac{ln(population SD)}{ln(population mean)}}}$ |

.

**Supplementary Table 3. Baseline and clinical characteristics of patients with/without heart failure before and after propensity score matching (1:1).**

* for SMD$\geq$0.2; SD: standard deviation; SCD: sudden cardiac death; VF: ventricular fibrillation; VT: ventricular tachycardia; SGLT2I: sodium glucose cotransporter-2 inhibitor; DPP4I: dipeptidyl peptidase-4 inhibitor; CV: coefficient of variation.

| **Characteristics** | **Before matching** |  | **SMD** | **After matching** |  | **SMD** |
| --- | --- | --- | --- | --- | --- | --- |
|  | **Heart failure (N=3489)**  **Mean(SD);N or Count(%)** | **No heart failure (N=55968) Mean(SD);N or Count(%)** |  | **Heart failure (N=1809)**  **Mean(SD);N or Count(%)** | **No heart failure (N=40185)**  **Mean(SD);N or Count(%)** |  |
| ***Demographics*** |  |  |  |  |  |  |
| Male gender | 1782(51.07%) | 30904(55.21%) | 0.08 | 1166(64.45%) | 25361(63.11%) | 0.03 |
| Baseline age, years | 73.0(12.0);n=3489 | 62.2(12.6);n=55968 | 0.88* | 69.5(11.0);n=1809 | 60.7(13.2);n=40185 | 0.73* |
| <50 | 151(4.32%) | 8421(15.04%) | 0.37* | 108(5.97%) | 6726(16.73%) | 0.34* |
| [50-60] | 394(11.29%) | 16109(28.78%) | 0.45* | 216(11.94%) | 14749(36.70%) | 0.60* |
| [60-70] | 722(20.69%) | 16635(29.72%) | 0.21* | 521(28.80%) | 9625(23.95%) | 0.11 |
| [70-80] | 1040(29.80%) | 9708(17.34%) | 0.30* | 715(39.52%) | 4679(11.64%) | 0.67* |
| >80 | 1182(33.87%) | 5100(9.11%) | 0.63* | 249(13.76%) | 4409(10.97%) | 0.08 |
| ***Past comorbidities*** |  |  |  |  |  |  |
| Charlson standard comorbidity index | 3.1(1.5);n=3489 | 1.9(1.4);n=55968 | 0.79* | 2.6(1.2);n=1809 | 1.8(1.4);n=40185 | 0.66* |
| Diabetes with chronic complication | 67(1.92%) | 530(0.94%) | 0.08 | 43(2.37%) | 411(1.02%) | 0.1 |
| Diabetes without chronic complication | 86(2.46%) | 935(1.67%) | 0.06 | 60(3.31%) | 812(2.02%) | 0.08 |
| Gout | 196(5.61%) | 1267(2.26%) | 0.17 | 80(4.42%) | 758(1.88%) | 0.15 |
| Hyperlipidaemia | 77(2.20%) | 1454(2.59%) | 0.03 | 27(1.49%) | 1418(3.52%) | 0.13 |
| Hypertension | 1207(34.59%) | 12055(21.53%) | 0.29* | 548(30.29%) | 8868(22.06%) | 0.19 |
| Hypoglycemia | 59(1.69%) | 383(0.68%) | 0.09 | 4(0.22%) | 96(0.23%) | 0 |
| Immune mediated enterocolitis | 192(5.50%) | 2737(4.89%) | 0.03 | 69(3.81%) | 2049(5.09%) | 0.06 |
| Ischemic heart disease | 390(11.17%) | 3679(6.57%) | 0.16 | 331(18.29%) | 3506(8.72%) | 0.28* |
| Liver diseases | 63(1.80%) | 1215(2.17%) | 0.03 | 32(1.76%) | 1225(3.04%) | 0.08 |
| Peripheral vascular disease | 70(2.00%) | 323(0.57%) | 0.13 | 19(1.05%) | 177(0.44%) | 0.07 |
| Renal diseases | 130(3.72%) | 842(1.50%) | 0.14 | 12(0.66%) | 198(0.49%) | 0.02 |
| Stroke/transient ischemic attack | 215(6.16%) | 1627(2.90%) | 0.16 | 77(4.25%) | 938(2.33%) | 0.11 |
| Atrial fibrillation | 222(6.36%) | 795(1.42%) | 0.26* | 99(5.47%) | 550(1.36%) | 0.23* |
| VT/VF/SCD | 8(0.22%) | 56(0.10%) | 0.03 | 4(0.22%) | 54(0.13%) | 0.02 |
| Anaemia | 272(7.79%) | 1957(3.49%) | 0.19 | 70(3.86%) | 840(2.09%) | 0.1 |
| Overweight | 19(0.54%) | 376(0.67%) | 0.02 | 34(1.87%) | 610(1.51%) | 0.03 |
| Cancer | 140(4.01%) | 1471(2.62%) | 0.08 | 42(2.32%) | 809(2.01%) | 0.02 |
| ***Medications*** |  |  |  |  |  |  |
| SGLT2I v.s. DPP4I | 638(18.28%) | 20359(36.37%) | 0.41* | 638(35.26%) | 20359(50.66%) | 0.31* |
| SGLT2I frequency | 7.8(12.6);n=638 | 7.1(9.6);n=20359 | 0.06 | 7.8(12.6);n=638 | 7.1(9.6);n=20359 | 0.06 |
| DPP4I frequency | 5.4(9.7);n=2851 | 5.3(7.2);n=35609 | 0.02 | 5.9(8.8);n=1171 | 6.9(6.9);n=19826 | 0.12 |
| SGLT2I duration, days | 334.0(560.6);n=638 | 534.0(672.4);n=20359 | 0.32* | 334.0(560.6);n=638 | 534.0(672.4);n=20359 | 0.32* |
| DPP4I duration, days | 462.9(468.7);n=2851 | 493.0(412.3);n=35609 | 0.07 | 392.0(433.7);n=1171 | 453.3(341.4);n=19826 | 0.16 |
| Metformin | 2694(77.21%) | 50359(89.97%) | 0.35* | 1540(85.12%) | 37444(93.17%) | 0.26* |
| Sulphonylurea | 2694(77.21%) | 42897(76.64%) | 0.01 | 957(52.90%) | 28841(71.77%) | 0.40* |
| Insulin | 2776(79.56%) | 26934(48.12%) | 0.69* | 1558(86.12%) | 20034(49.85%) | 0.84* |
| Acarbose | 66(1.89%) | 1439(2.57%) | 0.05 | 38(2.10%) | 1628(4.05%) | 0.11 |
| Thiazolidinedione | 346(9.91%) | 11102(19.83%) | 0.28* | 241(13.32%) | 11349(28.24%) | 0.37* |
| Glucagon-like peptide-1 receptor agonists | 39(1.11%) | 1654(2.95%) | 0.13 | 148(8.18%) | 2675(6.65%) | 0.06 |
| Statins and fibrates | 904(25.91%) | 27327(48.82%) | 0.49* | 969(53.56%) | 28682(71.37%) | 0.37* |
| ***Subclinical biomarkers*** |  |  |  |  |  |  |
| Neutrophil-to-lymphocyte ratio | 4.6(5.5);n=1991 | 3.4(4.7);n=21917 | 0.22* | 4.6(5.3);n=726 | 2.9(3.4);n=17788 | 0.39* |
| Platelet-to-lymphocyte ratio | 164.2(172.3);n=1991 | 143.8(149.0);n=21914 | 0.13 | 144.0(97.7);n=726 | 144.5(123.4);n=17786 | 0.01 |
| Neutrophil-to-high-density lipoprotein ratio | 0.31(0.27);n=1721 | 0.26(0.2);n=19915 | 0.19 | 0.4(0.2);n=659 | 0.3(0.2);n=14916 | 0.66* |
| Lymphocyte-to-high-density lipoprotein ratio | 0.09(0.06);n=1721 | 0.1(0.06);n=19914 | 0.18 | 0.13(0.08);n=659 | 0.11(0.05);n=14915 | 0.31* |
| Lymphocyte-to-low-density lipoprotein ratio | 0.0(0.0);n=1698 | 0.1(0.0);n=19586 | 0.08 | 0.1(0.0);n=652 | 0.0(0.1);n=14640 | 0.16 |
| Low density lipoprotein ratio-to-high density lipoprotein ratio | 2.09(0.96);n=2591 | 2.11(0.87);n=42813 | 0.02 | 2.2(0.7);n=1356 | 2.1(0.8);n=29595 | 0.11 |
| Total cholesterol-to-high density lipoprotein ratio | 3.82(1.4);n=2626 | 3.83(1.26);n=43488 | 0.01 | 4.0(1.0);n=1365 | 3.8(1.2);n=30007 | 0.1 |
| Triglyceride-glucose index | 7.5(0.7);n=2382 | 7.6(0.7);n=38948 | 0.05 | 7.7(0.7);n=1296 | 7.6(0.7);n=25841 | 0.09 |
| Bilirubin-to-albumin ratio | 0.28(0.18);n=2475 | 0.27(0.23);n=34835 | 0.04 | 0.28(0.17);n=881 | 0.27(0.17);n=26863 | 0.09 |
| Protein-to-creatinine ratio | 2.3(1.3);n=2264 | 3.1(1.6);n=27428 | 0.53* | 2.6(1.3);n=792 | 3.2(1.4);n=22643 | 0.46* |
| Prognostic nutritional index | 38.9(5.9);n=2504 | 41.2(6.4);n=35456 | 0.37* | 39.7(5.0);n=888 | 41.0(5.7);n=27187 | 0.25* |
| ***Complete blood counts*** |  |  |  |  |  |  |
| Mean corpuscular volume, fL | 88.0(7.7);n=2265 | 87.1(7.6);n=27559 | 0.12 | 87.6(6.5);n=791 | 86.0(7.7);n=22691 | 0.24* |
| Eosinophil, x10^9/L | 0.23(0.48);n=1990 | 0.22(0.22);n=21897 | 0.05 | 0.2(0.3);n=726 | 0.3(0.2);n=17784 | 0.15 |
| Lymphocyte, x10^9/L | 1.8(0.9);n=1991 | 2.0(0.9);n=21919 | 0.28* | 2.1(1.1);n=726 | 2.0(0.8);n=17789 | 0.12 |
| Neutrophil, x10^9/L | 5.9(3.1);n=1991 | 5.3(2.8);n=21919 | 0.21* | 6.6(3.3);n=726 | 4.8(2.1);n=17789 | 0.65* |
| White cell count, x10^9/L | 8.4(3.0);n=2268 | 8.0(3.0);n=27568 | 0.13 | 9.4(3.3);n=793 | 7.6(2.3);n=22697 | 0.63* |
| Mean cell haemoglobin, pg | 29.7(3.1);n=2265 | 29.4(3.0);n=27559 | 0.11 | 29.6(2.4);n=791 | 28.9(3.0);n=22691 | 0.28* |
| Platelet, x10^9/L | 229.5(73.2);n=2269 | 242.3(72.2);n=27565 | 0.18 | 231.7(66.3);n=793 | 250.1(65.2);n=22695 | 0.28* |
| Red cell count, x10^12/L | 4.2(0.7);n=2265 | 4.5(0.7);n=27559 | 0.49* | 4.4(0.7);n=791 | 4.5(0.7);n=22691 | 0.17 |
| ***Liver and renal functions*** |  |  |  |  |  |  |
| Potassium, mmol/L | 4.4(0.6);n=2982 | 4.3(0.5);n=46067 | 0.03 | 4.2(0.5);n=1443 | 4.3(0.5);n=34222 | 0.22* |
| Albumin, g/L | 39.3(4.5);n=2481 | 41.8(3.9);n=34911 | 0.61* | 39.9(4.0);n=883 | 41.5(3.8);n=26894 | 0.4* |
| Sodium, mmol/L | 139.1(3.7);n=2985 | 139.3(2.9);n=46089 | 0.06 | 139.6(4.1);n=1443 | 138.6(2.8);n=34221 | 0.27* |
| Urea, mmol/L | 8.5(4.9);n=2984 | 6.4(3.4);n=46074 | 0.5* | 8.0(3.7);n=1442 | 6.3(2.9);n=34203 | 0.51* |
| Protein, g/L | 72.6(6.7);n=2335 | 73.9(5.4);n=32855 | 0.22* | 72.7(5.8);n=863 | 73.1(5.3);n=25891 | 0.07 |
| Creatinine, umol/L | 124.6(103.4);n=2988 | 92.9(74.4);n=46215 | 0.35* | 116.2(52.7);n=1443 | 86.1(45.1);n=34252 | 0.61* |
| Alkaline phosphatase, U/L | 82.0(35.3);n=2484 | 76.6(32.5);n=35024 | 0.16 | 83.1(33.4);n=882 | 71.7(24.4);n=26926 | 0.39* |
| Aspartate transaminase, U/L | 28.3(47.2);n=1060 | 28.0(48.5);n=13741 | 0.01 | 26.7(26.2);n=393 | 27.5(29.3);n=9038 | 0.03 |
| Alanine transaminase, U/L | 24.0(30.5);n=2055 | 29.5(34.0);n=29902 | 0.17 | 22.6(22.3);n=1103 | 27.4(25.5);n=25473 | 0.2 |
| Bilirubin, umol/L | 10.8(6.6);n=2475 | 11.2(7.1);n=34844 | 0.05 | 11.2(6.6);n=881 | 11.0(6.2);n=26876 | 0.02 |
| ***Lipid and glucose profiles*** |  |  |  |  |  |  |
| Triglyceride, mmol/L | 1.66(1.17);n=2634 | 1.71(1.49);n=43546 | 0.04 | 1.7(1.0);n=1367 | 1.71(1.52);n=30036 | 0 |
| SD of triglyceride | 0.4(0.7);n=1380 | 0.5(1.0);n=22190 | 0.05 | 0.5(0.7);n=579 | 0.4(0.9);n=16965 | 0.13 |
| Low-density lipoprotein, mmol/L | 2.3(0.8);n=2593 | 2.4(0.8);n=42816 | 0.1 | 2.4(0.7);n=1357 | 2.3(0.7);n=29598 | 0.06 |
| SD of low-density lipoprotein | 0.38(0.37);n=1349 | 0.37(0.34);n=21575 | 0.05 | 0.5(0.4);n=522 | 0.3(0.4);n=16591 | 0.32* |
| High-density lipoprotein, mmol/L | 1.19(0.34);n=2626 | 1.2(0.33);n=43489 | 0.05 | 1.1(0.3);n=1365 | 1.2(0.3);n=30008 | 0.14 |
| SD of high-density lipoprotein | 0.12(0.11);n=1341 | 0.1(0.08);n=21338 | 0.21* | 0.12(0.08);n=566 | 0.09(0.07);n=16537 | 0.35* |
| Total cholesterol, mmol/L | 4.2(1.0);n=2641 | 4.4(1.0);n=43580 | 0.11 | 4.28(0.84);n=1370 | 4.25(0.93);n=30053 | 0.03 |
| SD of total cholesterol | 0.5(0.4);n=1366 | 0.4(0.4);n=22225 | 0.08 | 0.6(0.5);n=558 | 0.4(0.4);n=16807 | 0.34* |
| HbA1c, g/dL | 12.2(2.0);n=2292 | 13.2(1.8);n=28049 | 0.51* | 12.8(1.9);n=796 | 12.9(1.9);n=22818 | 0.04 |
| Mean HbA1c, g/dL | 12.0(1.9);n=2292 | 13.1(1.8);n=28050 | 0.58* | 12.7(1.7);n=796 | 12.8(1.9);n=22818 | 0.1 |
| Median HbA1c, g/dL | 12.0(1.9);n=2292 | 13.1(1.8);n=28050 | 0.58* | 12.7(1.7);n=796 | 12.8(1.9);n=22818 | 0.09 |
| Variance of HbA1c | 0.7(0.9);n=1723 | 0.5(1.0);n=15802 | 0.14 | 0.6(0.9);n=632 | 0.5(0.9);n=12333 | 0.09 |
| SD of HbA1c | 0.7(0.4);n=1723 | 0.6(0.5);n=15803 | 0.26* | 0.61(0.45);n=632 | 0.56(0.42);n=12333 | 0.11 |
| SD/Initial HbA1c | 0.1(0.0);n=1723 | 0.0(0.0);n=15803 | 0.28* | 0.1(0.1);n=632 | 0.0(0.0);n=12333 | 0.11 |
| CV of HbA1c | 0.1(0.0);n=1723 | 0.0(0.0);n=15801 | 0.34* | 0.05(0.04);n=632 | 0.04(0.04);n=12332 | 0.12 |
| Variability independent of mean HbA1c | 0.32(0.21);n=1723 | 0.26(0.21);n=15803 | 0.28* | 0.28(0.21);n=632 | 0.25(0.19);n=12333 | 0.11 |
| Fasting glucose, mmol/L | 9.0(5.0);n=2592 | 8.9(3.8);n=41004 | 0.02 | 9.2(7.1);n=1464 | 9.0(4.7);n=28931 | 0.03 |
| Mean fasting glucose, mmol/L | 8.8(3.3);n=2666 | 8.7(2.9);n=41883 | 0.03 | 8.6(3.6);n=1497 | 8.7(3.0);n=30141 | 0 |
| Median fasting glucose, mmol/L | 8.6(3.3);n=2666 | 8.7(2.9);n=41883 | 0.01 | 8.4(3.2);n=1497 | 8.6(2.9);n=30141 | 0.05 |
| Variance of fasting glucose | 14.6(45.1);n=1793 | 8.3(29.3);n=25158 | 0.17 | 22.2(104.3);n=734 | 8.8(34.6);n=19089 | 0.17 |
| SD of fasting glucose | 2.7(2.7);n=1793 | 1.9(2.1);n=25186 | 0.3* | 2.8(3.8);n=734 | 1.8(2.3);n=19107 | 0.3* |
| SD/Initial fasting glucose | 0.3(0.4);n=1793 | 0.2(0.3);n=25186 | 0.32* | 0.3(0.2);n=734 | 0.2(0.2);n=19107 | 0.32* |
| CV of fasting glucose | 0.22(0.17);n=1739 | 0.16(0.14);n=24670 | 0.41* | 0.2(0.2);n=716 | 0.1(0.1);n=18715 | 0.4* |
| Variability independent of mean fasting glucose | 0.6(0.5);n=1793 | 0.5(0.4);n=25186 | 0.36* | 0.6(0.5);n=734 | 0.4(0.4);n=19107 | 0.36* |

**Supplementary Table 4. Baseline and clinical characteristics of patients with/without myocardial infarction before and after propensity score matching (1:1).**

* for SMD$\geq$0.2; SD: standard deviation; SCD: sudden cardiac death; VF: ventricular fibrillation; VT: ventricular tachycardia; SGLT2I: sodium glucose cotransporter-2 inhibitor; DPP4I: dipeptidyl peptidase-4 inhibitor; CV: coefficient of variation.

| **Characteristics** | **Before matching** |  | **SMD** | **After matching** |  | **SMD** |
| --- | --- | --- | --- | --- | --- | --- |
|  | **Myocardial infarction (N=2610) Mean(SD);N or Count(%)** | **No myocardial infarction (N=56847) Mean(SD);N or Count(%)** |  | **Myocardial infarction (N=1649) Mean(SD);N or Count(%)** | **No myocardial infarction (N=40345) Mean(SD);N or Count(%)** |  |
| ***Demographics*** |  |  |  |  |  |  |
| Male gender | 1536(58.85%) | 31150(54.79%) | 0.08 | 1265(76.71%) | 25262(62.61%) | 0.31* |
| Baseline age, years | 70.1(12.6);n=2610 | 62.5(12.7);n=56847 | 0.6* | 66.7(10.6);n=1649 | 60.8(13.2);n=40345 | 0.49* |
| <50 | 171(6.55%) | 8401(14.77%) | 0.27* | 86(5.21%) | 6748(16.72%) | 0.37* |
| [50-60] | 451(17.27%) | 16052(28.23%) | 0.26* | 487(29.53%) | 14478(35.88%) | 0.14 |
| [60-70] | 586(22.45%) | 16771(29.50%) | 0.16 | 255(15.46%) | 9891(24.51%) | 0.23* |
| [70-80] | 719(27.54%) | 10029(17.64%) | 0.24* | 638(38.69%) | 4756(11.78%) | 0.65* |
| >80 | 683(26.16%) | 5599(9.84%) | 0.43* | 183(11.09%) | 4475(11.09%) | 0 |
| ***Past comorbidities*** |  |  |  |  |  |  |
| Charlson standard comorbidity index | 2.8(1.5);n=2610 | 2.0(1.4);n=56847 | 0.55* | 2.4(1.3);n=1649 | 1.8(1.4);n=40345 | 0.43* |
| Diabetes with chronic complication | 53(2.03%) | 544(0.95%) | 0.09 | 44(2.66%) | 410(1.01%) | 0.12 |
| Diabetes without chronic complication | 54(2.06%) | 967(1.70%) | 0.03 | 47(2.85%) | 825(2.04%) | 0.05 |
| Gout | 130(4.98%) | 1333(2.34%) | 0.14 | 30(1.81%) | 808(2.00%) | 0.01 |
| Hyperlipidaemia | 72(2.75%) | 1459(2.56%) | 0.01 | 39(2.36%) | 1406(3.48%) | 0.07 |
| Hypertension | 822(31.49%) | 12440(21.88%) | 0.22* | 252(15.28%) | 9164(22.71%) | 0.19 |
| Hypoglycemia | 52(1.99%) | 390(0.68%) | 0.11 | 2(0.12%) | 98(0.24%) | 0.03 |
| Immune mediated enterocolitis | 171(6.55%) | 2758(4.85%) | 0.07 | 65(3.94%) | 2053(5.08%) | 0.06 |
| Ischemic heart disease | 315(12.06%) | 3754(6.60%) | 0.19 | 398(24.13%) | 3439(8.52%) | 0.43* |
| Liver diseases | 36(1.37%) | 1242(2.18%) | 0.06 | 19(1.15%) | 1238(3.06%) | 0.13 |
| Peripheral vascular disease | 55(2.10%) | 338(0.59%) | 0.13 | 16(0.97%) | 180(0.44%) | 0.06 |
| Renal diseases | 126(4.82%) | 846(1.48%) | 0.19 | 8(0.48%) | 202(0.50%) | 0 |
| Stroke/transient ischemic attack | 140(5.36%) | 1702(2.99%) | 0.12 | 51(3.09%) | 964(2.38%) | 0.04 |
| Atrial fibrillation | 80(3.06%) | 937(1.64%) | 0.09 | 37(2.24%) | 612(1.51%) | 0.05 |
| VT/VF/SCD | 4(0.15%) | 60(0.10%) | 0.01 | 3(0.18%) | 55(0.13%) | 0.01 |
| Anaemia | 176(6.74%) | 2053(3.61%) | 0.14 | 61(3.69%) | 849(2.10%) | 0.1 |
| Overweight | 8(0.30%) | 387(0.68%) | 0.05 | 6(0.36%) | 638(1.58%) | 0.12 |
| Cancer | 78(2.98%) | 1533(2.69%) | 0.02 | 14(0.84%) | 837(2.07%) | 0.1 |
| ***Medications*** |  |  |  |  |  |  |
| SGLT2I v.s. DPP4I | 631(24.17%) | 20366(35.82%) | 0.26* | 631(38.26%) | 20366(50.47%) | 0.25* |
| SGLT2I frequency | 7.6(12.5);n=631 | 7.1(9.6);n=20366 | 0.04 | 7.6(12.5);n=631 | 7.1(9.6);n=20366 | 0.04 |
| DPP4I frequency | 5.1(9.1);n=1979 | 5.3(7.3);n=36481 | 0.02 | 5.6(6.9);n=1018 | 6.9(7.0);n=19979 | 0.19 |
| SGLT2I duration, days | 343.4(561.1);n=631 | 533.6(672.4);n=20366 | 0.31* | 343.4(561.1);n=631 | 533.6(672.4);n=20366 | 0.31* |
| DPP4I duration, days | 447.8(464.7);n=1979 | 493.1(414.0);n=36481 | 0.1 | 236.6(252.9);n=1018 | 460.8(348.2);n=19979 | 0.74* |
| Metformin | 2038(78.08%) | 51015(89.74%) | 0.32* | 1509(91.51%) | 37475(92.88%) | 0.05 |
| Sulphonylurea | 2031(77.81%) | 43560(76.62%) | 0.03 | 952(57.73%) | 28846(71.49%) | 0.29* |
| Insulin | 2135(81.80%) | 27575(48.50%) | 0.75* | 1136(68.89%) | 20456(50.70%) | 0.38* |
| Acarbose | 59(2.26%) | 1446(2.54%) | 0.02 | 131(7.94%) | 1535(3.80%) | 0.18 |
| Thiazolidinedione | 228(8.73%) | 11220(19.73%) | 0.32* | 149(9.03%) | 11441(28.35%) | 0.51* |
| Glucagon-like peptide-1 receptor agonists | 33(1.26%) | 1660(2.92%) | 0.12 | 44(2.66%) | 2779(6.88%) | 0.2 |
| Statins and fibrates | 784(30.03%) | 27447(48.28%) | 0.38* | 965(58.52%) | 28686(71.10%) | 0.27* |
| ***Subclinical biomarkers*** |  |  |  |  |  |  |
| Neutrophil-to-lymphocyte ratio | 5.0(7.4);n=1417 | 3.4(4.5);n=22491 | 0.25* | 3.3(4.1);n=720 | 2.9(3.5);n=17794 | 0.1 |
| Platelet-to-lymphocyte ratio | 172.9(207.5);n=1417 | 143.7(146.7);n=22488 | 0.16 | 146.8(102.0);n=720 | 144.4(123.2);n=17792 | 0.02 |
| Neutrophil-to-high-density lipoprotein ratio | 0.32(0.23);n=1237 | 0.26(0.2);n=20399 | 0.25* | 0.3(0.16);n=672 | 0.26(0.16);n=14903 | 0.27* |
| Lymphocyte-to-high-density lipoprotein ratio | 0.1(0.06);n=1237 | 0.1(0.06);n=20398 | 0.1 | 0.12(0.06);n=672 | 0.11(0.05);n=14902 | 0.19 |
| Lymphocyte-to-low-density lipoprotein ratio | 0.0(0.0);n=1214 | 0.1(0.0);n=20070 | 0.15 | 0.1(0.0);n=664 | 0.0(0.1);n=14628 | 0.09 |
| Low density lipoprotein ratio-to-high density lipoprotein ratio | 2.3(1.0);n=1911 | 2.1(0.9);n=43493 | 0.19 | 2.3(0.7);n=1399 | 2.1(0.8);n=29552 | 0.29* |
| Total cholesterol-to-high density lipoprotein ratio | 4.1(1.5);n=1946 | 3.8(1.3);n=44168 | 0.19 | 4.1(1.1);n=1409 | 3.8(1.2);n=29963 | 0.28* |
| Triglyceride-glucose index | 7.61(0.74);n=1762 | 7.56(0.72);n=39568 | 0.07 | 7.7(0.5);n=1324 | 7.6(0.7);n=25813 | 0.12 |
| Bilirubin-to-albumin ratio | 0.26(0.19);n=1793 | 0.27(0.23);n=35517 | 0.04 | 0.2(0.1);n=955 | 0.3(0.2);n=26789 | 0.29* |
| Protein-to-creatinine ratio | 2.4(1.4);n=1632 | 3.0(1.5);n=28060 | 0.45* | 2.9(1.1);n=769 | 3.2(1.4);n=22666 | 0.25* |
| Prognostic nutritional index | 39.0(6.7);n=1823 | 41.1(6.4);n=36137 | 0.33* | 41.7(5.8);n=964 | 41.0(5.7);n=27111 | 0.14 |
| ***Complete blood counts*** |  |  |  |  |  |  |
| Mean corpuscular volume, fL | 88.1(7.5);n=1639 | 87.1(7.6);n=28185 | 0.13 | 90.5(8.2);n=780 | 85.9(7.6);n=22702 | 0.58* |
| Eosinophil, x10^9/L | 0.23(0.25);n=1416 | 0.22(0.25);n=22471 | 0.07 | 0.2(0.1);n=720 | 0.3(0.2);n=17790 | 0.34* |
| Lymphocyte, x10^9/L | 1.8(0.9);n=1417 | 2.0(0.9);n=22493 | 0.23* | 1.96(0.82);n=720 | 1.98(0.8);n=17795 | 0.02 |
| Neutrophil, x10^9/L | 6.1(3.3);n=1417 | 5.3(2.8);n=22493 | 0.27* | 5.2(2.6);n=720 | 4.8(2.2);n=17795 | 0.14 |
| White cell count, x10^9/L | 8.6(3.2);n=1639 | 8.0(3.0);n=28197 | 0.2* | 7.9(2.8);n=780 | 7.7(2.4);n=22710 | 0.09 |
| Mean cell haemoglobin, pg | 29.9(3.0);n=1639 | 29.4(3.0);n=28185 | 0.15 | 30.4(3.0);n=780 | 28.9(3.0);n=22702 | 0.5* |
| Platelet, x10^9/L | 236.8(73.8);n=1639 | 241.6(72.3);n=28195 | 0.07 | 242.1(57.9);n=780 | 249.7(65.6);n=22708 | 0.12 |
| Red cell count, x10^12/L | 4.2(0.8);n=1639 | 4.5(0.7);n=28185 | 0.42* | 4.4(0.7);n=780 | 4.5(0.7);n=22702 | 0.2 |
| ***Liver and renal functions*** |  |  |  |  |  |  |
| Potassium, mmol/L | 4.4(0.6);n=2171 | 4.3(0.5);n=46878 | 0.02 | 4.3(0.51);n=1452 | 4.3(0.51);n=34213 | 0.01 |
| Albumin, g/L | 39.6(4.8);n=1796 | 41.8(3.9);n=35596 | 0.51* | 42.1(4.1);n=955 | 41.4(3.8);n=26822 | 0.18 |
| Sodium, mmol/L | 138.9(3.6);n=2172 | 139.3(2.9);n=46902 | 0.11 | 140.1(2.9);n=1451 | 138.6(2.8);n=34213 | 0.52* |
| Urea, mmol/L | 8.7(5.6);n=2169 | 6.5(3.4);n=46889 | 0.48* | 7.8(3.1);n=1451 | 6.3(2.9);n=34194 | 0.49* |
| Protein, g/L | 72.4(6.6);n=1680 | 73.9(5.4);n=33510 | 0.24* | 72.1(6.6);n=936 | 73.1(5.2);n=25818 | 0.17 |
| Creatinine, umol/L | 137.9(141.7);n=2172 | 92.8(71.8);n=47031 | 0.4* | 112.4(51.1);n=1451 | 86.3(45.3);n=34244 | 0.54* |
| Alkaline phosphatase, U/L | 83.9(38.9);n=1799 | 76.6(32.4);n=35709 | 0.2* | 69.7(25.6);n=955 | 72.2(24.8);n=26853 | 0.1 |
| Aspartate transaminase, U/L | 33.6(142.4);n=750 | 27.7(37.3);n=14051 | 0.06 | 20.3(18.5);n=635 | 28.0(29.7);n=8796 | 0.31* |
| Alanine transaminase, U/L | 24.4(34.0);n=1494 | 29.3(33.8);n=30463 | 0.15 | 20.4(12.8);n=1127 | 27.5(25.8);n=25449 | 0.35* |
| Bilirubin, umol/L | 10.3(6.9);n=1793 | 11.2(7.1);n=35526 | 0.14 | 9.4(4.4);n=955 | 11.1(6.2);n=26802 | 0.32* |
| ***Lipid and glucose profiles*** |  |  |  |  |  |  |
| Triglyceride, mmol/L | 1.8(1.5);n=1955 | 1.7(1.5);n=44225 | 0.06 | 1.8(1.3);n=1410 | 1.7(1.5);n=29993 | 0.04 |
| SD of triglyceride | 0.53(1.07);n=1071 | 0.48(0.96);n=22499 | 0.05 | 0.8(1.2);n=397 | 0.4(0.9);n=17147 | 0.35* |
| Low-density lipoprotein, mmol/L | 2.5(0.9);n=1912 | 2.4(0.8);n=43497 | 0.09 | 2.4(0.7);n=1400 | 2.3(0.7);n=29555 | 0.05 |
| SD of low-density lipoprotein | 0.42(0.39);n=1034 | 0.36(0.34);n=21890 | 0.15 | 0.4(0.3);n=343 | 0.3(0.4);n=16770 | 0.21* |
| High-density lipoprotein, mmol/L | 1.16(0.35);n=1946 | 1.2(0.33);n=44169 | 0.14 | 1.1(0.3);n=1409 | 1.2(0.3);n=29964 | 0.35* |
| SD of high-density lipoprotein | 0.12(0.1);n=1048 | 0.1(0.08);n=21631 | 0.17 | 0.1(0.08);n=389 | 0.09(0.07);n=16714 | 0.13 |
| Total cholesterol, mmol/L | 4.4(1.1);n=1959 | 4.3(1.0);n=44262 | 0.07 | 4.2(0.9);n=1411 | 4.3(0.9);n=30012 | 0.03 |
| SD of total cholesterol | 0.5(0.5);n=1066 | 0.4(0.4);n=22525 | 0.18 | 0.5(0.4);n=401 | 0.4(0.4);n=16964 | 0.17 |
| HbA1c, g/dL | 12.4(2.1);n=1660 | 13.2(1.8);n=28681 | 0.39* | 13.1(1.8);n=784 | 12.9(1.9);n=22830 | 0.1 |
| Mean HbA1c, g/dL | 12.2(2.0);n=1660 | 13.1(1.8);n=28682 | 0.47* | 13.0(1.6);n=784 | 12.8(1.9);n=22830 | 0.09 |
| Median HbA1c, g/dL | 12.2(2.0);n=1660 | 13.1(1.8);n=28682 | 0.47* | 13.0(1.6);n=784 | 12.8(1.9);n=22830 | 0.07 |
| Variance of HbA1c | 0.7(1.2);n=1204 | 0.5(1.0);n=16321 | 0.19 | 1.0(1.8);n=375 | 0.5(0.9);n=12590 | 0.38* |
| SD of HbA1c | 0.7(0.5);n=1204 | 0.6(0.4);n=16322 | 0.3* | 0.7(0.7);n=375 | 0.6(0.4);n=12590 | 0.33* |
| SD/Initial HbA1c | 0.1(0.1);n=1204 | 0.0(0.0);n=16322 | 0.31* | 0.1(0.1);n=375 | 0.0(0.0);n=12590 | 0.3* |
| CV of HbA1c | 0.1(0.0);n=1204 | 0.0(0.0);n=16320 | 0.38* | 0.1(0.1);n=375 | 0.0(0.0);n=12589 | 0.32* |
| Variability independent of mean HbA1c | 0.33(0.22);n=1204 | 0.26(0.21);n=16322 | 0.32* | 0.34(0.32);n=375 | 0.25(0.19);n=12590 | 0.33* |
| Fasting glucose, mmol/L | 9.3(5.3);n=1907 | 8.9(3.8);n=41689 | 0.09 | 8.8(4.1);n=1360 | 9.0(4.8);n=29035 | 0.05 |
| Mean fasting glucose, mmol/L | 9.1(3.6);n=1956 | 8.7(2.9);n=42593 | 0.11 | 8.6(2.9);n=1396 | 8.7(3.0);n=30242 | 0.02 |
| Median fasting glucose, mmol/L | 8.9(3.5);n=1956 | 8.6(2.9);n=42593 | 0.07 | 8.5(2.8);n=1396 | 8.6(3.0);n=30242 | 0.03 |
| Variance of fasting glucose | 15.9(50.8);n=1315 | 8.3(29.2);n=25636 | 0.18 | 16.4(62.9);n=479 | 9.1(38.8);n=19344 | 0.14 |
| SD of fasting glucose | 2.8(2.9);n=1315 | 1.9(2.1);n=25664 | 0.33* | 2.7(3.1);n=479 | 1.9(2.4);n=19362 | 0.3* |
| SD/Initial fasting glucose | 0.3(0.3);n=1315 | 0.2(0.3);n=25664 | 0.3* | 0.3(0.3);n=479 | 0.2(0.2);n=19362 | 0.33* |
| CV of fasting glucose | 0.23(0.17);n=1259 | 0.16(0.14);n=25150 | 0.41* | 0.2(0.2);n=461 | 0.1(0.1);n=18970 | 0.41* |
| Variability independent of mean fasting glucose | 0.6(0.5);n=1315 | 0.5(0.4);n=25664 | 0.38* | 0.6(0.5);n=479 | 0.4(0.4);n=19362 | 0.37* |

**Supplementary Table 5. Baseline characteristics of patients with/without mortality risks before and after propensity score matching (1:1).**

* for SMD$\geq$0.2; SD: standard deviation; SCD: sudden cardiac death; VF: ventricular fibrillation; VT: ventricular tachycardia; SGLT2I: sodium glucose cotransporter-2 inhibitor; DPP4I: dipeptidyl peptidase-4 inhibitor; CV: coefficient of variation.

| **Characteristics** | **Before matching** |  | **SMD** |  |  | **SMD** | **After matching** |  | **SMD** |  |  | **SMD** |
| --- | --- | --- | --- | --- | --- | --- | --- | --- | --- | --- | --- | --- |
|  | **All-cause mortality (N=6143) Mean(SD);N or Count(%)** | **Alive (N=53314) Mean(SD);N or Count(%)** |  | **Cardiovascular mortality (N=1863) Mean(SD);N or Count(%)** | **No cardiovascular mortality (N=57594) Mean(SD);N or Count(%)** |  | **All-cause mortality (N=2674) Mean(SD);N or Count(%)** | **Alive (N=39320) Mean(SD);N or Count(%)** |  | **Cardiovascular mortality (N=538) Mean(SD);N or Count(%)** | **No cardiovascular mortality (N=41456) Mean(SD);N or Count(%)** |  |
| ***Demographics*** |  |  |  |  |  |  |  |  |  |  |  |  |
| Male gender | 3357(54.64%) | 29329(55.01%) | 0.01 | 1088(58.40%) | 31598(54.86%) | 0.07 | 1622(60.65%) | 24905(63.33%) | 0.06 | 334(62.08%) | 26193(63.18%) | 0.02 |
| Female gender | 2786(45.35%) | 23985(44.98%) | 0.01 | 775(41.59%) | 25996(45.13%) | 0.07 | 1052(39.34%) | 14415(36.66%) | 0.06 | 204(37.91%) | 15263(36.81%) | 0.02 |
| Baseline age, years | 74.8(11.7);n=6143 | 61.5(12.2);n=53314 | 1.11* | 78.4(10.4);n=1863 | 62.4(12.6);n=57594 | 1.39* | 69.2(13.4);n=2674 | 60.5(13.0);n=39320 | 0.66* | 81.2(9.6);n=538 | 60.8(13.0);n=41456 | 1.78* |
| <50 | 185(3.01%) | 8387(15.73%) | 0.45* | 24(1.28%) | 8548(14.84%) | 0.51* | 274(10.24%) | 6560(16.68%) | 0.19 | 2(0.37%) | 6832(16.48%) | 0.61* |
| [50-60] | 583(9.49%) | 15920(29.86%) | 0.53* | 92(4.93%) | 16411(28.49%) | 0.67* | 634(23.70%) | 14331(36.44%) | 0.28* | 12(2.23%) | 14953(36.06%) | 0.95* |
| [60-70] | 1149(18.70%) | 16208(30.40%) | 0.27* | 235(12.61%) | 17122(29.72%) | 0.43* | 248(9.27%) | 9898(25.17%) | 0.43* | 65(12.08%) | 10081(24.31%) | 0.32* |
| [70-80] | 1798(29.26%) | 8950(16.78%) | 0.30* | 521(27.96%) | 10227(17.75%) | 0.24* | 871(32.57%) | 4523(11.50%) | 0.53* | 141(26.20%) | 5253(12.67%) | 0.35* |
| >80 | 2428(39.52%) | 3854(7.22%) | 0.83* | 991(53.19%) | 5291(9.18%) | 1.08* | 647(24.19%) | 4011(10.20%) | 0.38* | 318(59.10%) | 4340(10.46%) | 1.19* |
| ***Past comorbidities*** |  |  |  |  |  |  |  |  |  |  |  |  |
| Charlson standard comorbidity index | 3.4(1.6);n=6143 | 1.9(1.3);n=53314 | 1.06* | 3.7(1.4);n=1863 | 2.0(1.4);n=57594 | 1.26* | 2.7(1.7);n=2674 | 1.8(1.3);n=39320 | 0.6* | 4.0(1.2);n=538 | 1.8(1.4);n=41456 | 1.72* |
| Diabetes with chronic complication | 122(1.98%) | 475(0.89%) | 0.09 | 28(1.50%) | 569(0.98%) | 0.05 | 48(1.79%) | 406(1.03%) | 0.06 | 16(2.97%) | 438(1.05%) | 0.14 |
| Diabetes without chronic complication | 157(2.55%) | 864(1.62%) | 0.07 | 55(2.95%) | 966(1.67%) | 0.08 | 99(3.70%) | 773(1.96%) | 0.1 | 53(9.85%) | 819(1.97%) | 0.34* |
| Gout | 319(5.19%) | 1144(2.14%) | 0.16 | 122(6.54%) | 1341(2.32%) | 0.21* | 87(3.25%) | 751(1.90%) | 0.08 | 23(4.27%) | 815(1.96%) | 0.13 |
| Hyperlipidaemia | 134(2.18%) | 1397(2.62%) | 0.03 | 43(2.30%) | 1488(2.58%) | 0.02 | 52(1.94%) | 1393(3.54%) | 0.1 | 15(2.78%) | 1430(3.44%) | 0.04 |
| Hypertension | 2131(34.68%) | 11131(20.87%) | 0.31* | 695(37.30%) | 12567(21.81%) | 0.34* | 802(29.99%) | 8614(21.90%) | 0.19 | 265(49.25%) | 9151(22.07%) | 0.59* |
| Hypoglycemia | 136(2.21%) | 306(0.57%) | 0.14 | 47(2.52%) | 395(0.68%) | 0.15 | 19(0.71%) | 81(0.20%) | 0.07 | 7(1.30%) | 93(0.22%) | 0.12 |
| Immune mediated enterocolitis | 358(5.82%) | 2571(4.82%) | 0.04 | 121(6.49%) | 2808(4.87%) | 0.07 | 141(5.27%) | 1977(5.02%) | 0.01 | 47(8.73%) | 2071(4.99%) | 0.15 |
| Ischemic heart disease | 502(8.17%) | 3567(6.69%) | 0.06 | 147(7.89%) | 3922(6.80%) | 0.04 | 267(9.98%) | 3570(9.07%) | 0.03 | 90(16.72%) | 3747(9.03%) | 0.23* |
| Liver diseases | 138(2.24%) | 1140(2.13%) | 0.01 | 34(1.82%) | 1244(2.15%) | 0.02 | 60(2.24%) | 1197(3.04%) | 0.05 | 12(2.23%) | 1245(3.00%) | 0.05 |
| Peripheral vascular disease | 130(2.11%) | 263(0.49%) | 0.14 | 40(2.14%) | 353(0.61%) | 0.13 | 49(1.83%) | 147(0.37%) | 0.14 | 7(1.30%) | 189(0.45%) | 0.09 |
| Renal diseases | 358(5.82%) | 614(1.15%) | 0.26* | 102(5.47%) | 870(1.51%) | 0.22* | 28(1.04%) | 182(0.46%) | 0.07 | 17(3.15%) | 193(0.46%) | 0.20* |
| Stroke/transient ischemic attack | 410(6.67%) | 1432(2.68%) | 0.19 | 156(8.37%) | 1686(2.92%) | 0.24* | 137(5.12%) | 878(2.23%) | 0.15 | 80(14.86%) | 935(2.25%) | 0.46* |
| Atrial fibrillation | 251(4.08%) | 766(1.43%) | 0.16 | 96(5.15%) | 921(1.59%) | 0.2 | 117(4.37%) | 532(1.35%) | 0.18 | 50(9.29%) | 599(1.44%) | 0.35* |
| VT/VF/SCD | 17(0.27%) | 47(0.08%) | 0.04 | 4(0.21%) | 60(0.10%) | 0.03 | 13(0.48%) | 45(0.11%) | 0.07 | 4(0.74%) | 54(0.13%) | 0.09 |
| Anaemia | 600(9.76%) | 1629(3.05%) | 0.28* | 201(10.78%) | 2028(3.52%) | 0.28* | 120(4.48%) | 790(2.00%) | 0.14 | 40(7.43%) | 870(2.09%) | 0.25* |
| Overweight | 14(0.22%) | 381(0.71%) | 0.07 | 1(0.05%) | 394(0.68%) | 0.1 | 21(0.78%) | 623(1.58%) | 0.07 | 0(0.00%) | 644(1.55%) | 0.18 |
| Cancer | 346(5.63%) | 1265(2.37%) | 0.17 | 101(5.42%) | 1510(2.62%) | 0.14 | 107(4.00%) | 744(1.89%) | 0.12 | 31(5.76%) | 820(1.97%) | 0.2 |
| ***Medications*** |  |  |  |  |  |  |  |  |  |  |  |  |
| SGLT2I v.s. DPP4I | 527(8.57%) | 20470(38.39%) | 0.75* | 105(5.63%) | 20892(36.27%) | 0.81* | 527(19.70%) | 20470(52.06%) | 0.72* | 105(19.51%) | 20892(50.39%) | 0.68* |
| SGLT2I frequency | 10.8(15.5);n=527 | 7.1(9.5);n=20470 | 0.29* | 10.8(11.7);n=105 | 7.1(9.7);n=20892 | 0.34* | 10.8(15.5);n=527 | 7.1(9.5);n=20470 | 0.29* | 10.8(11.7);n=105 | 7.1(9.7);n=20892 | 0.34* |
| DPP4I frequency | 5.0(12.4);n=5616 | 5.3(6.2);n=32844 | 0.03 | 5.1(16.6);n=1758 | 5.3(6.7);n=36702 | 0.01 | 4.6(7.8);n=2147 | 7.1(6.9);n=18850 | 0.34* | 7.0(9.5);n=433 | 6.8(7.0);n=20564 | 0.02 |
| SGLT2I duration, days | 459.1(581.2);n=527 | 529.7(672.2);n=20470 | 0.11 | 433.1(551.4);n=105 | 528.4(670.6);n=20892 | 0.16 | 459.1(581.2);n=527 | 529.7(672.2);n=20470 | 0.11 | 433.1(551.4);n=105 | 528.4(670.6);n=20892 | 0.16 |
| DPP4I duration, days | 434.2(477.0);n=5616 | 500.4(404.9);n=32844 | 0.15 | 430.8(478.9);n=1758 | 493.6(413.4);n=36702 | 0.14 | 271.5(402.4);n=2147 | 470.2(334.7);n=18850 | 0.54* | 737.2(476.3);n=433 | 443.9(341.7);n=20564 | 0.71* |
| Metformin | 4450(72.44%) | 48603(91.16%) | 0.50* | 1338(71.81%) | 51715(89.79%) | 0.47* | 2328(87.06%) | 36656(93.22%) | 0.21* | 417(77.50%) | 38567(93.03%) | 0.45* |
| Sulphonylurea | 4772(77.68%) | 40819(76.56%) | 0.03 | 1507(80.89%) | 44084(76.54%) | 0.11 | 1292(48.31%) | 28506(72.49%) | 0.51* | 355(65.98%) | 29443(71.02%) | 0.11 |
| Insulin | 5074(82.59%) | 24636(46.20%) | 0.82* | 1639(87.97%) | 28071(48.73%) | 0.93* | 2426(90.72%) | 19166(48.74%) | 1.03* | 467(86.80%) | 21125(50.95%) | 0.84* |
| Acarbose | 153(2.49%) | 1352(2.53%) | 0 | 45(2.41%) | 1460(2.53%) | 0.01 | 115(4.30%) | 1551(3.94%) | 0.02 | 57(10.59%) | 1609(3.88%) | 0.26* |
| Thiozolidinedone | 531(8.64%) | 10917(20.47%) | 0.34* | 119(6.38%) | 11329(19.67%) | 0.40* | 967(36.16%) | 10623(27.01%) | 0.2 | 32(5.94%) | 11558(27.88%) | 0.61* |
| Glucagon-like peptide-1 receptor agonists | 20(0.32%) | 1673(3.13%) | 0.22* | 3(0.16%) | 1690(2.93%) | 0.23* | 60(2.24%) | 2763(7.02%) | 0.23* | 43(7.99%) | 2780(6.70%) | 0.05 |
| Statins and fibrates | 1387(22.57%) | 26844(50.35%) | 0.60* | 361(19.37%) | 27870(48.39%) | 0.64* | 1420(53.10%) | 28231(71.79%) | 0.39* | 301(55.94%) | 29350(70.79%) | 0.31* |
| ***Subclinical biomarkers*** |  |  |  |  |  |  |  |  |  |  |  |  |
| Neutrophil-to-lymphocyte ratio | 5.0(6.8);n=3471 | 3.3(4.3);n=20437 | 0.3* | 5.5(8.3);n=1090 | 3.4(4.5);n=22818 | 0.32* | 3.6(4.4);n=1161 | 2.9(3.4);n=17353 | 0.19 | 5.0(7.7);n=269 | 2.9(3.4);n=18245 | 0.35* |
| Platelet-to-lymphocyte ratio | 172.2(153.7);n=3470 | 140.9(150.3);n=20435 | 0.21* | 178.8(145.5);n=1090 | 143.9(151.2);n=22815 | 0.24* | 154.6(90.5);n=1160 | 143.8(124.3);n=17352 | 0.1 | 160.8(121.9);n=269 | 144.3(122.5);n=18243 | 0.13 |
| Neutrophil-to-high-density lipoprotein ratio | 0.3(0.22);n=2809 | 0.26(0.2);n=18827 | 0.16 | 0.3(0.24);n=845 | 0.27(0.2);n=20791 | 0.15 | 0.25(0.17);n=1065 | 0.26(0.16);n=14510 | 0.03 | 0.25(0.21);n=238 | 0.26(0.16);n=15337 | 0.02 |
| Lymphocyte-to-high-density lipoprotein ratio | 0.09(0.05);n=2809 | 0.1(0.06);n=18826 | 0.35* | 0.08(0.05);n=845 | 0.1(0.06);n=20790 | 0.43* | 0.09(0.04);n=1065 | 0.11(0.06);n=14509 | 0.38* | 0.07(0.04);n=238 | 0.11(0.05);n=15336 | 0.79* |
| Lymphocyte-to-low-density lipoprotein ratio | 0.0(0.0);n=2767 | 0.1(0.0);n=18517 | 0.23* | 0.0(0.0);n=835 | 0.1(0.0);n=20449 | 0.24* | 0.1(0.0);n=1058 | 0.0(0.2);n=14234 | 0.1 | 0.04(0.02);n=235 | 0.05(0.15);n=15057 | 0.05 |
| Low density lipoprotein ratio-to-high density lipoprotein ratio | 2.1(1.04);n=4315 | 2.11(0.85);n=41089 | 0.01 | 2.0(0.9);n=1267 | 2.1(0.9);n=44137 | 0.12 | 1.9(0.7);n=1787 | 2.1(0.8);n=29164 | 0.28* | 1.8(0.8);n=330 | 2.1(0.8);n=30621 | 0.38* |
| Total cholesterol-to-high density lipoprotein ratio | 3.8(1.44);n=4374 | 3.84(1.25);n=41740 | 0.03 | 3.7(1.3);n=1280 | 3.8(1.3);n=44834 | 0.13 | 3.5(1.1);n=1798 | 3.9(1.2);n=29574 | 0.36* | 3.4(1.1);n=334 | 3.9(1.2);n=31038 | 0.41* |
| Triglyceride-glucose index | 7.5(0.8);n=3946 | 7.6(0.7);n=37384 | 0.09 | 7.4(0.8);n=1139 | 7.6(0.7);n=40191 | 0.16 | 7.4(0.6);n=1741 | 7.6(0.7);n=25396 | 0.38* | 7.4(0.6);n=316 | 7.6(0.7);n=26821 | 0.23* |
| Bilirubin-to-albumin ratio | 0.29(0.32);n=4300 | 0.27(0.21);n=33010 | 0.07 | 0.27(0.16);n=1314 | 0.27(0.23);n=35996 | 0.01 | 0.27(0.16);n=1348 | 0.27(0.17);n=26396 | 0.02 | 0.27(0.15);n=326 | 0.27(0.17);n=27418 | 0 |
| Protein-to-creatinine ratio | 2.2(1.4);n=3983 | 3.1(1.5);n=25709 | 0.6* | 2.3(1.4);n=1254 | 3.0(1.5);n=28438 | 0.53* | 3.4(2.2);n=1289 | 3.2(1.4);n=22146 | 0.09 | 2.1(1.0);n=332 | 3.2(1.4);n=23103 | 0.89* |
| Prognostic nutritional index | 38.0(6.6);n=4365 | 41.4(6.3);n=33595 | 0.53* | 37.4(7.1);n=1342 | 41.2(6.3);n=36618 | 0.56* | 38.9(8.9);n=1397 | 41.1(5.5);n=26678 | 0.3* | 33.9(13.5);n=370 | 41.1(5.5);n=27705 | 0.7* |
| ***Complete blood counts*** |  |  |  |  |  |  |  |  |  |  |  |  |
| Mean corpuscular volume, fL | 88.2(8.1);n=4000 | 87.0(7.5);n=25824 | 0.15 | 88.1(8.3);n=1258 | 87.2(7.5);n=28566 | 0.12 | 89.8(7.4);n=1290 | 85.8(7.7);n=22192 | 0.53* | 89.8(8.6);n=332 | 86.0(7.7);n=23150 | 0.47* |
| Eosinophil, x10^9/L | 0.22(0.25);n=3464 | 0.22(0.25);n=20423 | 0.02 | 0.21(0.26);n=1089 | 0.22(0.25);n=22798 | 0.02 | 0.2(0.1);n=1161 | 0.3(0.2);n=17349 | 0.25* | 0.2(0.1);n=269 | 0.3(0.2);n=18241 | 0.4* |
| Lymphocyte, x10^9/L | 1.7(1.0);n=3471 | 2.1(0.9);n=20439 | 0.4* | 1.6(0.8);n=1090 | 2.0(0.9);n=22820 | 0.46* | 1.96(0.8);n=1161 | 1.98(0.8);n=17354 | 0.03 | 1.6(0.6);n=269 | 2.0(0.8);n=18246 | 0.49* |
| Neutrophil, x10^9/L | 5.9(3.4);n=3471 | 5.2(2.7);n=20439 | 0.22* | 6.2(3.8);n=1090 | 5.3(2.8);n=22820 | 0.27* | 5.6(2.4);n=1161 | 4.8(2.2);n=17354 | 0.33* | 5.9(3.3);n=269 | 4.8(2.2);n=18246 | 0.36* |
| White cell count, x10^9/L | 8.3(3.3);n=4000 | 8.0(3.0);n=25836 | 0.1 | 8.4(3.6);n=1258 | 8.0(3.0);n=28578 | 0.14 | 8.0(2.3);n=1290 | 7.7(2.4);n=22200 | 0.16 | 7.4(2.4);n=332 | 7.7(2.4);n=23158 | 0.11 |
| Mean cell haemoglobin, pg | 29.9(3.2);n=4000 | 29.4(3.0);n=25824 | 0.16 | 29.7(3.3);n=1258 | 29.4(3.0);n=28566 | 0.1 | 30.1(2.5);n=1290 | 28.8(3.0);n=22192 | 0.46* | 30.3(3.1);n=332 | 28.9(3.0);n=23150 | 0.45* |
| Platelet, x10^9/L | 228.6(80.1);n=3999 | 243.3(70.9);n=25835 | 0.19 | 228.9(78.5);n=1258 | 241.9(72.0);n=28576 | 0.17 | 262.3(99.6);n=1289 | 248.7(62.7);n=22199 | 0.16 | 210.5(56.9);n=332 | 250.0(65.3);n=23156 | 0.65* |
| Red cell count, x10^12/L | 4.1(0.7);n=4000 | 4.6(0.7);n=25824 | 0.69* | 4.1(0.7);n=1258 | 4.5(0.7);n=28566 | 0.64* | 4.1(0.7);n=1290 | 4.5(0.7);n=22192 | 0.57* | 4.0(0.8);n=332 | 4.5(0.7);n=23150 | 0.64* |
| ***Liver and renal functions*** |  |  |  |  |  |  |  |  |  |  |  |  |
| Potassium, mmol/L | 4.4(0.6);n=5203 | 4.3(0.5);n=43846 | 0.06 | 4.4(0.6);n=1588 | 4.3(0.5);n=47461 | 0.08 | 4.2(0.4);n=1977 | 4.3(0.5);n=33688 | 0.19 | 4.4(0.4);n=421 | 4.3(0.5);n=35244 | 0.11 |
| Albumin, g/L | 38.6(4.9);n=4304 | 42.1(3.6);n=33088 | 0.81* | 38.1(4.8);n=1316 | 41.8(3.9);n=36076 | 0.84* | 40.3(4.9);n=1348 | 41.5(3.8);n=26429 | 0.26* | 38.4(5.4);n=326 | 41.5(3.8);n=27451 | 0.65* |
| Sodium, mmol/L | 138.8(3.9);n=5208 | 139.3(2.8);n=43866 | 0.14 | 138.8(4.1);n=1590 | 139.3(2.9);n=47484 | 0.14 | 139.4(3.5);n=1977 | 138.6(2.8);n=33687 | 0.25* | 138.6(3.5);n=421 | 138.7(2.8);n=35243 | 0.02 |
| Urea, mmol/L | 8.9(5.7);n=5204 | 6.3(3.0);n=43854 | 0.58* | 8.6(5.0);n=1590 | 6.5(3.4);n=47468 | 0.5* | 7.9(3.4);n=1976 | 6.3(2.9);n=33669 | 0.5* | 7.8(3.5);n=422 | 6.4(2.9);n=35223 | 0.43* |
| Protein, g/L | 72.3(6.9);n=4070 | 74.1(5.3);n=31120 | 0.28* | 72.1(6.8);n=1247 | 73.9(5.4);n=33943 | 0.3* | 73.9(6.2);n=1322 | 73.0(5.2);n=25432 | 0.15 | 73.2(6.3);n=317 | 73.1(5.3);n=26437 | 0.02 |
| Creatinine, umol/L | 141.7(144.6);n=5212 | 89.2(61.9);n=43991 | 0.47* | 133.5(119.9);n=1591 | 93.5(74.6);n=47612 | 0.4* | 110.4(61.0);n=1978 | 86.0(44.4);n=33717 | 0.46* | 111.4(47.6);n=422 | 87.1(45.7);n=35273 | 0.52* |
| Alkaline phosphatase, U/L | 88.5(50.9);n=4311 | 75.5(29.3);n=33197 | 0.31* | 87.9(56.1);n=1317 | 76.6(31.5);n=36191 | 0.25* | 80.2(30.4);n=1350 | 71.7(24.4);n=26458 | 0.31* | 72.2(24.2);n=326 | 72.1(24.8);n=27482 | 0 |
| Aspartate transaminase, U/L | 28.8(50.8);n=1806 | 27.9(48.1);n=12995 | 0.02 | 28.0(61.27);n=511 | 28.03(47.92);n=14290 | 0 | 32.1(67.1);n=496 | 27.2(25.4);n=8935 | 0.1 | 21.1(10.3);n=143 | 27.5(29.4);n=9288 | 0.29* |
| Alanine transaminase, U/L | 24.0(36.9);n=3574 | 29.8(33.4);n=28383 | 0.16 | 21.7(34.6);n=1106 | 29.4(33.8);n=30851 | 0.23* | 23.5(42.8);n=1641 | 27.4(23.8);n=24935 | 0.11 | 16.7(14.8);n=278 | 27.3(25.5);n=26298 | 0.51* |
| Bilirubin, umol/L | 10.8(9.0);n=4300 | 11.2(6.8);n=33019 | 0.05 | 10.2(5.7);n=1314 | 11.2(7.1);n=36005 | 0.15 | 10.6(5.4);n=1348 | 11.1(6.2);n=26409 | 0.08 | 9.9(4.9);n=326 | 11.1(6.2);n=27431 | 0.21* |
| ***Lipid and glucose profiles*** |  |  |  |  |  |  |  |  |  |  |  |  |
| Triglyceride, mmol/L | 1.6(1.2);n=4380 | 1.7(1.5);n=41800 | 0.08 | 1.5(1.2);n=1284 | 1.7(1.5);n=44896 | 0.13 | 1.4(1.0);n=1799 | 1.7(1.5);n=29604 | 0.27* | 1.4(0.9);n=335 | 1.7(1.5);n=31068 | 0.22* |
| SD of triglyceride | 0.4(0.6);n=2107 | 0.5(1.0);n=21463 | 0.06 | 0.4(0.7);n=577 | 0.5(1.0);n=22993 | 0.07 | 0.38(0.6);n=495 | 0.43(0.93);n=17049 | 0.06 | 0.3(0.4);n=116 | 0.4(0.9);n=17428 | 0.16 |
| Low-density lipoprotein, mmol/L | 2.37(0.89);n=4316 | 2.4(0.79);n=41093 | 0.04 | 2.3(0.8);n=1267 | 2.4(0.8);n=44142 | 0.11 | 2.28(0.71);n=1788 | 2.35(0.74);n=29167 | 0.09 | 2.2(0.7);n=330 | 2.3(0.7);n=30625 | 0.14 |
| SD of low-density lipoprotein | 0.39(0.38);n=2056 | 0.36(0.34);n=20868 | 0.08 | 0.38(0.36);n=566 | 0.37(0.35);n=22358 | 0.04 | 0.35(0.38);n=483 | 0.35(0.35);n=16630 | 0 | 0.28(0.31);n=115 | 0.35(0.35);n=16998 | 0.2 |
| High-density lipoprotein, mmol/L | 1.22(0.38);n=4374 | 1.2(0.33);n=41741 | 0.04 | 1.25(0.4);n=1280 | 1.2(0.33);n=44835 | 0.12 | 1.3(0.3);n=1798 | 1.2(0.3);n=29575 | 0.34* | 1.3(0.4);n=334 | 1.2(0.3);n=31039 | 0.5* |
| SD of high-density lipoprotein | 0.13(0.11);n=2047 | 0.1(0.08);n=20632 | 0.3* | 0.14(0.13);n=564 | 0.1(0.08);n=22115 | 0.33* | 0.13(0.11);n=480 | 0.09(0.07);n=16623 | 0.38* | 0.13(0.11);n=112 | 0.09(0.07);n=16991 | 0.41* |
| Total cholesterol, mmol/L | 4.3(1.1);n=4389 | 4.4(1.0);n=41832 | 0.05 | 4.2(1.0);n=1286 | 4.4(1.0);n=44935 | 0.11 | 4.2(0.8);n=1801 | 4.3(0.9);n=29622 | 0.11 | 4.2(0.8);n=337 | 4.3(0.9);n=31086 | 0.03 |
| SD of total cholesterol | 0.5(0.5);n=2105 | 0.4(0.4);n=21486 | 0.11 | 0.5(0.4);n=569 | 0.4(0.4);n=23022 | 0.08 | 0.5(0.4);n=493 | 0.4(0.4);n=16872 | 0.07 | 0.5(0.4);n=114 | 0.4(0.4);n=17251 | 0.06 |
| HbA1c, g/dL | 12.0(2.0);n=4045 | 13.3(1.7);n=26296 | 0.68* | 11.9(1.8);n=1273 | 13.2(1.8);n=29068 | 0.68* | 12.3(1.9);n=1292 | 13.0(1.9);n=22322 | 0.35* | 11.9(2.0);n=332 | 12.9(1.9);n=23282 | 0.53* |
| Mean HbA1c, g/dL | 11.8(1.9);n=4045 | 13.2(1.7);n=26297 | 0.77* | 11.8(1.8);n=1273 | 13.1(1.8);n=29069 | 0.75* | 12.2(1.7);n=1292 | 12.9(1.9);n=22322 | 0.35* | 11.7(1.7);n=332 | 12.9(1.9);n=23282 | 0.68* |
| Median HbA1c, g/dL | 11.8(1.9);n=4045 | 13.2(1.7);n=26297 | 0.77* | 11.7(1.8);n=1273 | 13.1(1.8);n=29069 | 0.75* | 12.3(1.8);n=1292 | 12.9(1.9);n=22322 | 0.34* | 11.7(1.8);n=332 | 12.9(1.9);n=23282 | 0.61* |
| Variance of HbA1c | 0.8(1.1);n=2989 | 0.5(1.0);n=14536 | 0.25* | 0.7(1.0);n=920 | 0.5(1.0);n=16605 | 0.19 | 0.6(1.2);n=991 | 0.5(0.9);n=11974 | 0.08 | 1.1(1.8);n=246 | 0.5(0.9);n=12719 | 0.43* |
| SD of HbA1c | 0.7(0.5);n=2989 | 0.6(0.4);n=14537 | 0.38* | 0.7(0.5);n=920 | 0.6(0.5);n=16606 | 0.3* | 0.58(0.49);n=991 | 0.57(0.41);n=11974 | 0.04 | 0.8(0.7);n=246 | 0.6(0.4);n=12719 | 0.47* |
| SD/Initial HbA1c | 0.1(0.1);n=2989 | 0.0(0.0);n=14537 | 0.43* | 0.1(0.1);n=920 | 0.0(0.0);n=16606 | 0.34* | 0.05(0.05);n=991 | 0.05(0.04);n=11974 | 0.03 | 0.1(0.0);n=246 | 0.0(0.0);n=12719 | 0.41* |
| CV of HbA1c | 0.1(0.0);n=2989 | 0.0(0.0);n=14535 | 0.5* | 0.1(0.0);n=920 | 0.0(0.0);n=16604 | 0.4* | 0.04(0.04);n=991 | 0.04(0.04);n=11973 | 0.05 | 0.1(0.0);n=246 | 0.0(0.0);n=12718 | 0.41* |
| Variability independent of mean HbA1c | 0.3(0.2);n=2989 | 0.2(0.2);n=14537 | 0.41* | 0.33(0.22);n=920 | 0.26(0.21);n=16606 | 0.33* | 0.26(0.22);n=991 | 0.25(0.19);n=11974 | 0.04 | 0.4(0.3);n=246 | 0.3(0.2);n=12719 | 0.47* |
| Fasting glucose, mmol/L | 9.2(5.2);n=4503 | 8.9(3.7);n=39093 | 0.07 | 9.3(5.6);n=1343 | 8.9(3.8);n=42253 | 0.08 | 8.7(3.9);n=1838 | 9.0(4.9);n=28557 | 0.08 | 9.2(4.6);n=344 | 9.0(4.8);n=30051 | 0.04 |
| Mean fasting glucose, mmol/L | 9.0(3.7);n=4607 | 8.7(2.8);n=39942 | 0.09 | 9.1(4.0);n=1372 | 8.7(2.9);n=43177 | 0.11 | 8.6(2.9);n=1867 | 8.7(3.0);n=29771 | 0.03 | 9.0(3.8);n=351 | 8.7(3.0);n=31287 | 0.1 |
| Median fasting glucose, mmol/L | 8.8(3.6);n=4607 | 8.6(2.8);n=39942 | 0.06 | 8.9(4.0);n=1372 | 8.6(2.9);n=43177 | 0.09 | 8.5(2.9);n=1867 | 8.6(3.0);n=29771 | 0.04 | 8.8(3.9);n=351 | 8.6(2.9);n=31287 | 0.06 |
| Variance of fasting glucose | 15.5(42.6);n=3015 | 7.8(28.6);n=23936 | 0.21* | 17.4(50.4);n=888 | 8.4(29.7);n=26063 | 0.22* | 10.2(32.4);n=883 | 9.2(39.9);n=18940 | 0.03 | 11.3(27.3);n=223 | 9.3(39.7);n=19600 | 0.06 |
| SD of fasting glucose | 2.8(2.8);n=3015 | 1.9(2.1);n=23964 | 0.36* | 2.9(3.0);n=888 | 2.0(2.1);n=26091 | 0.35* | 2.2(2.3);n=883 | 1.9(2.4);n=18958 | 0.14 | 2.3(2.5);n=223 | 1.9(2.4);n=19618 | 0.18 |
| SD/Initial fasting glucose | 0.3(0.4);n=3015 | 0.2(0.3);n=23964 | 0.35* | 0.3(0.4);n=888 | 0.2(0.3);n=26091 | 0.33* | 0.24(0.25);n=883 | 0.18(0.22);n=18958 | 0.26* | 0.22(0.23);n=223 | 0.19(0.22);n=19618 | 0.17 |
| CV of fasting glucose | 0.23(0.17);n=2912 | 0.16(0.13);n=23497 | 0.45* | 0.23(0.17);n=852 | 0.16(0.14);n=25557 | 0.42* | 0.2(0.1);n=854 | 0.1(0.1);n=18577 | 0.28* | 0.2(0.2);n=208 | 0.1(0.1);n=19223 | 0.14 |
| Variability independent of mean fasting glucose | 0.6(0.5);n=3015 | 0.4(0.4);n=23964 | 0.42* | 0.6(0.5);n=888 | 0.5(0.4);n=26091 | 0.4* | 0.5(0.4);n=883 | 0.4(0.4);n=18958 | 0.21* | 0.5(0.4);n=223 | 0.4(0.4);n=19618 | 0.18 |

**Supplementary Table 6. Baseline and clinical characteristics of patients stratified by age and gender before and after propensity score matching (1:1).**

* for SMD$\geq$0.2; SD: standard deviation; SCD: sudden cardiac death; VF: ventricular fibrillation; VT: ventricular tachycardia; SGLT2I: sodium glucose cotransporter-2 inhibitor; DPP4I: dipeptidyl peptidase-4 inhibitor; CV: coefficient of variation.

| **Characteristics** | **Before matching** |  | **SMD** | **After matching** |  |  | **Before matching** |  |  | **After matching** |  |  |
| --- | --- | --- | --- | --- | --- | --- | --- | --- | --- | --- | --- | --- |
|  | **Age>=65 (N=25253) Mean(SD);N or Count(%)** | **Age<65 (N=34204) Mean(SD);N or Count(%)** |  | **Age>=65 (N=14375) Mean(SD);N or Count(%)** | **Age<65 (N=27619) Mean(SD);N or Count(%)** | **SMD** | **Male gender (N=32686) Mean(SD);N or Count(%)** | **Female gender (N=26771) Mean(SD);N or Count(%)** | **SMD** | **Male gender (N=26527) Mean(SD);N or Count(%)** | **Female gender (N=15467) Mean(SD);N or Count(%)** | **SMD** |
| ***Adverse events*** |  |  |  |  |  |  |  |  |  |  |  |  |
| All-cause mortality | 4862(19.25%) | 1281(3.74%) | 0.50* | 1653(11.49%) | 1021(3.69%) | 0.30* | 3357(10.27%) | 2786(10.40%) | 0 | 1622(6.11%) | 1052(6.80%) | 0.03 |
| Cardiovascular mortality | 1658(6.56%) | 205(0.59%) | 0.33* | 509(3.54%) | 29(0.10%) | 0.26* | 1088(3.32%) | 775(2.89%) | 0.02 | 334(1.25%) | 204(1.31%) | 0.01 |
| Myocardial infarction | 1694(6.70%) | 916(2.67%) | 0.19 | 931(6.47%) | 718(2.59%) | 0.19 | 1536(4.69%) | 1074(4.01%) | 0.03 | 1265(4.76%) | 384(2.48%) | 0.12 |
| Heart failure | 2612(10.34%) | 877(2.56%) | 0.32* | 1232(8.57%) | 577(2.08%) | 0.29* | 1782(5.45%) | 1707(6.37%) | 0.04 | 1166(4.39%) | 643(4.15%) | 0.01 |
| ***Demographics*** |  |  |  |  |  |  |  |  |  |  |  |  |
| Male gender | 12311(48.75%) | 20375(59.56%) | 0.22* | 8462(58.86%) | 18065(65.40%) | 0.14 | 32686(100.00%) | 0(0.00%) | inf* | 26527(100.00%) | 0(0.00%) | inf* |
| Female gender | 12942(51.24%) | 13829(40.43%) | 0.22* | 5913(41.13%) | 9554(34.59%) | 0.14 | 0(0.00%) | 26771(100.00%) | inf* | 0(0.00%) | 15467(100.00%) | inf* |
| Baseline age, years | 74.8(7.1);n=25253 | 54.1(8.2);n=34204 | 2.7* | 75.8(7.4);n=14375 | 53.4(8.1);n=27619 | 2.89* | 61.5(12.4);n=32686 | 64.6(13.2);n=26771 | 0.24* | 60.4(13.2);n=26527 | 62.1(13.2);n=15467 | 0.13 |
| <50 | 0(0.00%) | 8572(25.06%) | 0.82* | 0(0.00%) | 6834(24.74%) | 0.81* | 5267(16.11%) | 3305(12.34%) | 0.11 | 4512(17.00%) | 2322(15.01%) | 0.05 |
| [50-60] | 0(0.00%) | 16503(48.24%) | 1.37* | 0(0.00%) | 14965(54.18%) | 1.54* | 9994(30.57%) | 6509(24.31%) | 0.14 | 10338(38.97%) | 4627(29.91%) | 0.19 |
| [60-70] | 8225(32.57%) | 9132(26.69%) | 0.13 | 4323(30.07%) | 5823(21.08%) | 0.21* | 9501(29.06%) | 7856(29.34%) | 0.01 | 5332(20.10%) | 4814(31.12%) | 0.25* |
| [70-80] | 10748(42.56%) | 0(0.00%) | 1.22* | 5394(37.52%) | 0(0.00%) | 1.10* | 5384(16.47%) | 5364(20.03%) | 0.09 | 3443(12.97%) | 1951(12.61%) | 0.01 |
| >80 | 6282(24.87%) | 0(0.00%) | 0.81* | 4658(32.40%) | 0(0.00%) | 0.98* | 2543(7.78%) | 3739(13.96%) | 0.2 | 2904(10.94%) | 1754(11.34%) | 0.01 |
| ***Past comorbidities*** |  |  |  |  |  |  |  |  |  |  |  |  |
| Charlson standard comorbidity index | 3.2(1.2);n=25253 | 1.1(0.9);n=34204 | 1.96* | 3.2(1.0);n=14375 | 1.1(0.9);n=27619 | 2.28* | 1.9(1.4);n=32686 | 2.2(1.5);n=26771 | 0.22* | 1.7(1.4);n=26527 | 1.9(1.4);n=15467 | 0.15 |
| Diabetes with chronic complication | 280(1.10%) | 317(0.92%) | 0.02 | 172(1.19%) | 282(1.02%) | 0.02 | 308(0.94%) | 289(1.07%) | 0.01 | 231(0.87%) | 223(1.44%) | 0.05 |
| Diabetes without chronic complication | 414(1.63%) | 607(1.77%) | 0.01 | 318(2.21%) | 554(2.00%) | 0.01 | 460(1.40%) | 561(2.09%) | 0.05 | 408(1.53%) | 464(2.99%) | 0.1 |
| Gout | 912(3.61%) | 551(1.61%) | 0.13 | 365(2.53%) | 473(1.71%) | 0.06 | 966(2.95%) | 497(1.85%) | 0.07 | 569(2.14%) | 269(1.73%) | 0.03 |
| Hyperlipidaemia | 698(2.76%) | 833(2.43%) | 0.02 | 558(3.88%) | 887(3.21%) | 0.04 | 784(2.39%) | 747(2.79%) | 0.02 | 832(3.13%) | 613(3.96%) | 0.04 |
| Hypertension | 7846(31.06%) | 5416(15.83%) | 0.37* | 4316(30.02%) | 5100(18.46%) | 0.27* | 6583(20.14%) | 6679(24.94%) | 0.12 | 4943(18.63%) | 4473(28.91%) | 0.24* |
| Hypoglycemia | 346(1.37%) | 96(0.28%) | 0.12 | 61(0.42%) | 39(0.14%) | 0.05 | 167(0.51%) | 275(1.02%) | 0.06 | 46(0.17%) | 54(0.34%) | 0.03 |
| Immune mediated enterocolitis | 1351(5.34%) | 1578(4.61%) | 0.03 | 778(5.41%) | 1340(4.85%) | 0.03 | 1228(3.75%) | 1701(6.35%) | 0.12 | 887(3.34%) | 1231(7.95%) | 0.20* |
| Ischemic heart disease | 2505(9.91%) | 1564(4.57%) | 0.21* | 2191(15.24%) | 1646(5.95%) | 0.31* | 2634(8.05%) | 1435(5.36%) | 0.11 | 2478(9.34%) | 1359(8.78%) | 0.02 |
| Liver diseases | 426(1.68%) | 852(2.49%) | 0.06 | 356(2.47%) | 901(3.26%) | 0.05 | 736(2.25%) | 542(2.02%) | 0.02 | 745(2.80%) | 512(3.31%) | 0.03 |
| Peripheral vascular disease | 273(1.08%) | 120(0.35%) | 0.09 | 123(0.85%) | 73(0.26%) | 0.08 | 249(0.76%) | 144(0.53%) | 0.03 | 123(0.46%) | 73(0.47%) | 0 |
| Renal diseases | 590(2.33%) | 382(1.11%) | 0.09 | 101(0.70%) | 109(0.39%) | 0.04 | 592(1.81%) | 380(1.41%) | 0.03 | 118(0.44%) | 92(0.59%) | 0.02 |
| Stroke/transient ischemic attack | 1284(5.08%) | 558(1.63%) | 0.19 | 589(4.09%) | 426(1.54%) | 0.15 | 955(2.92%) | 887(3.31%) | 0.02 | 537(2.02%) | 478(3.09%) | 0.07 |
| Atrial fibrillation | 775(3.06%) | 242(0.70%) | 0.17 | 467(3.24%) | 182(0.65%) | 0.19 | 537(1.64%) | 480(1.79%) | 0.01 | 316(1.19%) | 333(2.15%) | 0.08 |
| VT/VF/SCD | 43(0.17%) | 21(0.06%) | 0.03 | 39(0.27%) | 19(0.06%) | 0.05 | 43(0.13%) | 21(0.07%) | 0.02 | 43(0.16%) | 15(0.09%) | 0.02 |
| Anaemia | 1425(5.64%) | 804(2.35%) | 0.17 | 411(2.85%) | 499(1.80%) | 0.07 | 810(2.47%) | 1419(5.30%) | 0.15 | 310(1.16%) | 600(3.87%) | 0.17 |
| Overweight | 37(0.14%) | 358(1.04%) | 0.12 | 79(0.54%) | 565(2.04%) | 0.13 | 212(0.64%) | 183(0.68%) | 0 | 268(1.01%) | 376(2.43%) | 0.11 |
| Cancer | 1098(4.34%) | 513(1.49%) | 0.17 | 514(3.57%) | 337(1.22%) | 0.15 | 741(2.26%) | 870(3.24%) | 0.06 | 333(1.25%) | 518(3.34%) | 0.14 |
| ***Medications*** |  |  |  |  |  |  |  |  |  |  |  |  |
| SGLT2I v.s. DPP4I | 5255(20.80%) | 15742(46.02%) | 0.55* | 5255(36.55%) | 15742(56.99%) | 0.42* | 12403(37.94%) | 8594(32.10%) | 0.12 | 12403(46.75%) | 8594(55.56%) | 0.18 |
| SGLT2I frequency | 8.6(11.1);n=5255 | 6.7(9.1);n=15742 | 0.19 | 8.6(11.1);n=5255 | 6.7(9.1);n=15742 | 0.19 | 6.7(9.0);n=12403 | 7.8(10.6);n=8594 | 0.11 | 6.7(9.0);n=12403 | 7.8(10.6);n=8594 | 0.11 |
| DPP4I frequency | 5.28(8.56);n=19998 | 5.26(5.94);n=18462 | 0 | 7.6(7.2);n=9120 | 6.3(6.8);n=11877 | 0.19 | 5.1(7.7);n=20283 | 5.4(7.1);n=18177 | 0.04 | 6.0(6.1);n=14124 | 8.5(8.4);n=6873 | 0.34* |
| SGLT2I duration, days | 581.7(704.0);n=5255 | 510.0(657.5);n=15742 | 0.11 | 581.7(704.0);n=5255 | 510.0(657.5);n=15742 | 0.11 | 497.1(660.4);n=12403 | 572.4(681.5);n=8594 | 0.11 | 497.1(660.4);n=12403 | 572.4(681.5);n=8594 | 0.11 |
| DPP4I duration, days | 488.9(435.8);n=19998 | 492.7(395.3);n=18462 | 0.01 | 453.8(336.5);n=9120 | 446.9(355.7);n=11877 | 0.02 | 472.0(416.6);n=20283 | 511.6(416.2);n=18177 | 0.09 | 400.6(339.6);n=14124 | 551.3(341.6);n=6873 | 0.44* |
| Metformin | 21436(84.88%) | 31617(92.43%) | 0.24* | 12695(88.31%) | 26289(95.18%) | 0.25* | 28980(88.66%) | 24073(89.92%) | 0.04 | 25038(94.38%) | 13946(90.16%) | 0.16 |
| Sulphonylurea | 19963(79.05%) | 25628(74.92%) | 0.1 | 10312(71.73%) | 19486(70.55%) | 0.03 | 25017(76.53%) | 20574(76.85%) | 0.01 | 18660(70.34%) | 11138(72.01%) | 0.04 |
| Insulin | 13985(55.37%) | 15725(45.97%) | 0.19 | 6976(48.52%) | 14616(52.92%) | 0.09 | 16096(49.24%) | 13614(50.85%) | 0.03 | 13011(49.04%) | 8581(55.47%) | 0.13 |
| Acarbose | 768(3.04%) | 737(2.15%) | 0.06 | 781(5.43%) | 885(3.20%) | 0.11 | 791(2.41%) | 714(2.66%) | 0.02 | 930(3.50%) | 736(4.75%) | 0.06 |
| Thiozolidinedone | 3345(13.24%) | 8103(23.69%) | 0.27* | 2654(18.46%) | 8936(32.35%) | 0.32* | 6223(19.03%) | 5225(19.51%) | 0.01 | 5939(22.38%) | 5651(36.53%) | 0.31* |
| Glucagon-like peptide-1 receptor agonists | 194(0.76%) | 1499(4.38%) | 0.23* | 362(2.51%) | 2461(8.91%) | 0.28* | 985(3.01%) | 708(2.64%) | 0.02 | 1718(6.47%) | 1105(7.14%) | 0.03 |
| Statins and fibrates | 9767(38.67%) | 18464(53.98%) | 0.31* | 10395(72.31%) | 19256(69.72%) | 0.06 | 15708(48.05%) | 12523(46.77%) | 0.03 | 18348(69.16%) | 11303(73.07%) | 0.09 |
| ***Subclinical biomarkers*** |  |  |  |  |  |  |  |  |  |  |  |  |
| Neutrophil-to-lymphocyte ratio | 4.0(5.4);n=11363 | 3.1(4.0);n=12545 | 0.19 | 3.1(3.4);n=8205 | 2.8(3.5);n=10309 | 0.09 | 3.6(4.7);n=12941 | 3.5(4.8);n=10967 | 0.03 | 3.0(3.5);n=11863 | 2.9(3.5);n=6651 | 0.02 |
| Platelet-to-lymphocyte ratio | 153.3(140.7);n=11361 | 138.4(159.7);n=12544 | 0.1 | 155.0(87.1);n=8204 | 136.2(144.0);n=10308 | 0.16 | 140.5(143.6);n=12940 | 151.3(159.4);n=10965 | 0.07 | 145.8(110.6);n=11862 | 142.2(141.1);n=6650 | 0.03 |
| Neutrophil-to-high-density lipoprotein ratio | 0.26(0.21);n=9965 | 0.27(0.2);n=11671 | 0.03 | 0.2(0.1);n=5675 | 0.3(0.2);n=9900 | 0.15 | 0.3(0.2);n=11872 | 0.2(0.2);n=9764 | 0.18 | 0.3(0.2);n=9337 | 0.2(0.2);n=6238 | 0.19 |
| Lymphocyte-to-high-density lipoprotein ratio | 0.09(0.05);n=9965 | 0.11(0.06);n=11670 | 0.32* | 0.09(0.05);n=5675 | 0.11(0.06);n=9899 | 0.43* | 0.11(0.06);n=11872 | 0.1(0.05);n=9763 | 0.17 | 0.11(0.06);n=9337 | 0.1(0.05);n=6237 | 0.12 |
| Lymphocyte-to-low-density lipoprotein ratio | 0.05(0.05);n=9876 | 0.05(0.03);n=11408 | 0.03 | 0.0(0.2);n=5631 | 0.1(0.0);n=9661 | 0.11 | 0.05(0.03);n=11669 | 0.05(0.05);n=9615 | 0.01 | 0.1(0.0);n=9220 | 0.0(0.2);n=6072 | 0.07 |
| Low density lipoprotein ratio-to-high density lipoprotein ratio | 2.0(0.8);n=19822 | 2.2(0.9);n=25582 | 0.32* | 1.9(0.7);n=9791 | 2.2(0.8);n=21160 | 0.4* | 2.2(0.9);n=24459 | 2.0(0.8);n=20945 | 0.25* | 2.2(0.8);n=18789 | 1.9(0.8);n=12162 | 0.38* |
| Total cholesterol-to-high density lipoprotein ratio | 3.6(1.1);n=19999 | 4.0(1.3);n=26115 | 0.33* | 3.6(1.0);n=9864 | 4.0(1.2);n=21508 | 0.39* | 4.0(1.3);n=24874 | 3.7(1.2);n=21240 | 0.24* | 4.0(1.1);n=18991 | 3.6(1.1);n=12381 | 0.34* |
| Triglyceride-glucose index | 7.4(0.7);n=17696 | 7.6(0.7);n=23634 | 0.28* | 7.4(0.6);n=9033 | 7.7(0.7);n=18104 | 0.38* | 7.5(0.7);n=22306 | 7.6(0.7);n=19024 | 0.04 | 7.58(0.69);n=15782 | 7.62(0.68);n=11355 | 0.05 |
| Bilirubin-to-albumin ratio | 0.27(0.23);n=16364 | 0.27(0.22);n=20946 | 0 | 0.26(0.2);n=10605 | 0.27(0.14);n=17139 | 0.1 | 0.3(0.3);n=20158 | 0.2(0.2);n=17152 | 0.18 | 0.27(0.15);n=16493 | 0.27(0.18);n=11251 | 0.01 |
| Protein-to-creatinine ratio | 2.6(1.4);n=13851 | 3.4(1.6);n=15841 | 0.53* | 2.7(1.2);n=9509 | 3.6(1.4);n=13926 | 0.64* | 2.5(1.1);n=15977 | 3.6(1.7);n=13715 | 0.79* | 2.7(1.1);n=13947 | 3.9(1.6);n=9488 | 0.88* |
| Prognostic nutritional index | 40.2(6.4);n=16659 | 41.7(6.3);n=21301 | 0.24* | 39.1(5.8);n=10730 | 42.1(5.4);n=17345 | 0.55* | 41.3(6.4);n=20492 | 40.7(6.4);n=17468 | 0.09 | 40.9(5.6);n=16649 | 41.1(5.8);n=11426 | 0.04 |
| ***Complete blood counts*** |  |  |  |  |  |  |  |  |  |  |  |  |
| Mean corpuscular volume, fL | 88.2(7.6);n=13900 | 86.4(7.4);n=15924 | 0.24* | 87.1(6.8);n=9519 | 85.2(8.2);n=13963 | 0.25* | 87.7(7.4);n=16035 | 86.6(7.7);n=13789 | 0.14 | 85.5(8.3);n=13973 | 86.8(6.7);n=9509 | 0.18 |
| Eosinophil, x10^9/L | 0.22(0.3);n=11346 | 0.22(0.2);n=12541 | 0.01 | 0.3(0.3);n=8202 | 0.2(0.2);n=10308 | 0.42* | 0.24(0.25);n=12931 | 0.19(0.24);n=10956 | 0.23* | 0.3(0.2);n=11859 | 0.2(0.2);n=6651 | 0.34* |
| Lymphocyte, x10^9/L | 1.9(0.9);n=11363 | 2.1(0.9);n=12547 | 0.32* | 1.7(0.6);n=8205 | 2.2(0.9);n=10310 | 0.6* | 1.97(0.93);n=12941 | 2.04(0.89);n=10969 | 0.08 | 1.9(0.8);n=11863 | 2.1(0.7);n=6652 | 0.33* |
| Neutrophil, x10^9/L | 5.4(3.0);n=11363 | 5.2(2.7);n=12547 | 0.06 | 4.6(2.1);n=8205 | 5.1(2.3);n=10310 | 0.25* | 5.34(2.85);n=12941 | 5.31(2.83);n=10969 | 0.01 | 4.7(2.1);n=11863 | 5.1(2.4);n=6652 | 0.17 |
| White cell count, x10^9/L | 7.9(3.2);n=13904 | 8.1(2.8);n=15932 | 0.04 | 7.4(2.1);n=9522 | 7.9(2.5);n=13968 | 0.23* | 8.04(3.15);n=16039 | 7.95(2.83);n=13797 | 0.03 | 7.68(2.34);n=13977 | 7.69(2.48);n=9513 | 0 |
| Mean cell haemoglobin, pg | 29.7(3.0);n=13900 | 29.2(3.0);n=15924 | 0.18 | 29.3(2.6);n=9519 | 28.7(3.3);n=13963 | 0.19 | 29.7(3.0);n=16035 | 29.1(3.1);n=13789 | 0.21* | 28.8(3.3);n=13973 | 29.1(2.6);n=9509 | 0.12 |
| Platelet, x10^9/L | 231.4(71.5);n=13903 | 250.0(72.0);n=15931 | 0.26* | 236.3(58.3);n=9521 | 258.4(68.3);n=13967 | 0.35* | 228.3(67.0);n=16039 | 256.4(75.4);n=13795 | 0.39* | 240.9(62.5);n=13976 | 262.0(67.4);n=9512 | 0.32* |
| Red cell count, x10^12/L | 4.3(0.6);n=13900 | 4.7(0.6);n=15924 | 0.67* | 4.1(0.6);n=9519 | 4.8(0.6);n=13963 | 1.16* | 4.7(0.7);n=16035 | 4.3(0.6);n=13789 | 0.57* | 4.5(0.8);n=13973 | 4.4(0.5);n=9509 | 0.16 |
| ***Liver and renal functions*** |  |  |  |  |  |  |  |  |  |  |  |  |
| Potassium, mmol/L | 4.4(0.5);n=21861 | 4.3(0.5);n=27188 | 0.2 | 4.28(0.59);n=13323 | 4.32(0.45);n=22342 | 0.08 | 4.4(0.5);n=26298 | 4.3(0.5);n=22751 | 0.01 | 4.3(0.5);n=21754 | 4.4(0.5);n=13911 | 0.24* |
| Albumin, g/L | 40.8(4.0);n=16403 | 42.3(3.8);n=20989 | 0.38* | 39.5(4.1);n=10616 | 42.6(3.1);n=17161 | 0.84* | 41.9(4.0);n=20194 | 41.4(3.9);n=17198 | 0.13 | 41.2(4.2);n=16507 | 41.7(3.2);n=11270 | 0.12 |
| Sodium, mmol/L | 139.3(3.2);n=21879 | 139.2(2.8);n=27195 | 0.05 | 138.4(3.0);n=13325 | 138.8(2.7);n=22339 | 0.15 | 139.2(2.9);n=26306 | 139.4(3.0);n=22768 | 0.07 | 138.6(2.7);n=21752 | 138.9(3.0);n=13912 | 0.1 |
| Urea, mmol/L | 7.3(3.7);n=21868 | 6.0(3.3);n=27190 | 0.37* | 7.6(3.3);n=13325 | 5.7(2.5);n=22320 | 0.65* | 6.8(3.6);n=26300 | 6.3(3.5);n=22758 | 0.13 | 6.7(3.2);n=21743 | 5.9(2.4);n=13902 | 0.27* |
| Protein, g/L | 73.5(5.7);n=15386 | 74.2(5.3);n=19804 | 0.13 | 71.4(5.6);n=10300 | 74.1(4.8);n=16454 | 0.52* | 73.5(5.5);n=19013 | 74.3(5.5);n=16177 | 0.14 | 72.5(5.5);n=15923 | 73.9(4.9);n=10831 | 0.26* |
| Creatinine, umol/L | 104.3(67.8);n=21937 | 87.1(82.6);n=27266 | 0.23* | 102.7(55.0);n=13337 | 78.2(36.5);n=22358 | 0.53* | 105.6(83.3);n=26383 | 82.3(66.5);n=22820 | 0.31* | 95.9(50.9);n=21768 | 73.9(32.4);n=13927 | 0.52* |
| Alkaline phosphatase, U/L | 77.5(34.1);n=16455 | 76.6(31.6);n=21053 | 0.03 | 69.1(26.1);n=10624 | 73.9(23.8);n=17184 | 0.19 | 75.7(33.7);n=20249 | 78.5(31.6);n=17259 | 0.09 | 69.6(23.9);n=16517 | 75.7(25.7);n=11291 | 0.25* |
| Aspartate transaminase, U/L | 26.6(66.9);n=6307 | 29.1(27.7);n=8494 | 0.05 | 26.4(40.8);n=2888 | 27.9(22.2);n=6543 | 0.05 | 28.7(33.3);n=8191 | 27.2(62.3);n=6610 | 0.03 | 27.0(21.3);n=5512 | 28.0(37.6);n=3919 | 0.03 |
| Alanine transaminase, U/L | 24.2(37.3);n=14321 | 33.1(30.1);n=17636 | 0.26* | 20.9(24.5);n=10270 | 31.1(25.2);n=16306 | 0.41* | 31.2(31.1);n=17142 | 26.7(36.6);n=14815 | 0.13 | 27.6(23.0);n=16817 | 26.4(29.1);n=9759 | 0.05 |
| Bilirubin, umol/L | 10.9(7.1);n=16368 | 11.3(7.1);n=20951 | 0.06 | 10.1(6.8);n=10606 | 11.6(5.7);n=17151 | 0.24* | 12.0(7.8);n=20160 | 10.2(6.0);n=17159 | 0.25* | 11.0(6.1);n=16493 | 11.1(6.3);n=11264 | 0.02 |
| ***Lipid and glucose profiles*** |  |  |  |  |  |  |  |  |  |  |  |  |
| Triglyceride, mmol/L | 1.5(1.0);n=20023 | 1.8(1.7);n=26157 | 0.21* | 1.5(1.0);n=9873 | 1.8(1.7);n=21530 | 0.19 | 1.71(1.61);n=24912 | 1.71(1.29);n=21268 | 0 | 1.7(1.46);n=19009 | 1.72(1.55);n=12394 | 0.02 |
| SD of triglyceride | 0.4(0.5);n=9159 | 0.6(1.1);n=14411 | 0.2* | 0.3(0.5);n=4965 | 0.5(1.0);n=12579 | 0.18 | 0.5(1.0);n=13016 | 0.4(0.9);n=10554 | 0.07 | 0.44(0.95);n=9680 | 0.4(0.88);n=7864 | 0.05 |
| Low-density lipoprotein, mmol/L | 2.3(0.8);n=19823 | 2.5(0.8);n=25586 | 0.3* | 2.2(0.7);n=9792 | 2.4(0.7);n=21163 | 0.33* | 2.37(0.8);n=24463 | 2.43(0.81);n=20946 | 0.08 | 2.3(0.7);n=18792 | 2.4(0.8);n=12163 | 0.21* |
| SD of low-density lipoprotein | 0.3(0.3);n=9057 | 0.4(0.4);n=13867 | 0.1 | 0.3(0.3);n=4981 | 0.4(0.4);n=12132 | 0.29* | 0.36(0.34);n=12609 | 0.38(0.36);n=10315 | 0.07 | 0.3(0.3);n=9546 | 0.4(0.4);n=7567 | 0.33* |
| High-density lipoprotein, mmol/L | 1.23(0.34);n=19999 | 1.18(0.32);n=26116 | 0.16 | 1.2(0.4);n=9864 | 1.1(0.3);n=21509 | 0.17 | 1.1(0.3);n=24875 | 1.3(0.3);n=21240 | 0.5* | 1.1(0.3);n=18992 | 1.3(0.4);n=12381 | 0.75* |
| SD of high-density lipoprotein | 0.11(0.09);n=8859 | 0.1(0.08);n=13820 | 0.1 | 0.1(0.07);n=4864 | 0.09(0.07);n=12239 | 0.06 | 0.1(0.08);n=12509 | 0.11(0.09);n=10170 | 0.16 | 0.08(0.07);n=9458 | 0.11(0.07);n=7645 | 0.37* |
| Total cholesterol, mmol/L | 4.2(0.9);n=20051 | 4.5(1.0);n=26170 | 0.3* | 4.1(0.9);n=9884 | 4.3(1.0);n=21539 | 0.29* | 4.2(1.0);n=24931 | 4.5(1.0);n=21290 | 0.24* | 4.1(0.9);n=19017 | 4.5(1.0);n=12406 | 0.42* |
| SD of total cholesterol | 0.4(0.4);n=9177 | 0.5(0.4);n=14414 | 0.12 | 0.3(0.3);n=4940 | 0.5(0.5);n=12425 | 0.37* | 0.4(0.4);n=13008 | 0.5(0.4);n=10583 | 0.07 | 0.4(0.4);n=9716 | 0.5(0.5);n=7649 | 0.24* |
| HbA1c, g/dL | 12.6(1.8);n=14168 | 13.6(1.8);n=16173 | 0.6* | 11.9(1.9);n=9559 | 13.6(1.6);n=14055 | 0.98* | 13.8(1.8);n=16306 | 12.4(1.6);n=14035 | 0.82* | 13.0(2.2);n=14051 | 12.8(1.4);n=9563 | 0.08 |
| Mean HbA1c, g/dL | 12.4(1.7);n=14169 | 13.5(1.7);n=16173 | 0.63* | 11.8(1.9);n=9559 | 13.6(1.5);n=14055 | 1.05* | 13.7(1.8);n=16307 | 12.3(1.5);n=14035 | 0.83* | 12.9(2.1);n=14051 | 12.8(1.4);n=9563 | 0.05 |
| Median HbA1c, g/dL | 12.4(1.7);n=14169 | 13.5(1.7);n=16173 | 0.62* | 11.8(1.9);n=9559 | 13.5(1.5);n=14055 | 1.03* | 13.6(1.8);n=16307 | 12.3(1.5);n=14035 | 0.82* | 12.9(2.1);n=14051 | 12.8(1.4);n=9563 | 0.05 |
| Variance of HbA1c | 0.6(0.9);n=8661 | 0.5(1.1);n=8864 | 0.01 | 0.54(0.8);n=6374 | 0.46(1.02);n=6591 | 0.09 | 0.6(1.1);n=9327 | 0.5(0.9);n=8198 | 0.05 | 0.5(1.0);n=8349 | 0.4(0.8);n=4616 | 0.1 |
| SD of HbA1c | 0.61(0.44);n=8662 | 0.58(0.46);n=8864 | 0.07 | 0.6(0.4);n=6374 | 0.5(0.4);n=6591 | 0.3* | 0.6(0.46);n=9328 | 0.58(0.44);n=8198 | 0.06 | 0.6(0.4);n=8349 | 0.5(0.4);n=4616 | 0.16 |
| SD/Initial HbA1c | 0.1(0.0);n=8662 | 0.0(0.0);n=8864 | 0.14 | 0.1(0.0);n=6374 | 0.0(0.0);n=6591 | 0.43* | 0.0(0.0);n=9328 | 0.1(0.0);n=8198 | 0.07 | 0.05(0.04);n=8349 | 0.04(0.04);n=4616 | 0.14 |
| CV of HbA1c | 0.05(0.04);n=8661 | 0.04(0.04);n=8863 | 0.16 | 0.1(0.0);n=6373 | 0.0(0.0);n=6591 | 0.52* | 0.04(0.04);n=9327 | 0.04(0.04);n=8197 | 0.05 | 0.04(0.04);n=8348 | 0.04(0.03);n=4616 | 0.2 |
| Variability independent of mean HbA1c | 0.27(0.2);n=8662 | 0.25(0.21);n=8864 | 0.1 | 0.3(0.2);n=6374 | 0.2(0.2);n=6591 | 0.36* | 0.27(0.21);n=9328 | 0.26(0.21);n=8198 | 0.02 | 0.3(0.2);n=8349 | 0.2(0.2);n=4616 | 0.15 |
| Fasting glucose, mmol/L | 8.5(3.7);n=18968 | 9.2(4.0);n=24628 | 0.16 | 7.8(4.5);n=11686 | 9.7(4.8);n=18709 | 0.4* | 8.91(4.0);n=23402 | 8.89(3.78);n=20194 | 0.01 | 8.6(4.7);n=18333 | 9.5(4.9);n=12062 | 0.18 |
| Mean fasting glucose, mmol/L | 8.5(2.8);n=19487 | 8.9(3.0);n=25062 | 0.15 | 7.7(2.7);n=11980 | 9.2(3.0);n=19658 | 0.51* | 8.7(2.9);n=23965 | 8.8(3.0);n=20584 | 0.03 | 8.4(2.9);n=19167 | 9.1(3.1);n=12471 | 0.25* |
| Median fasting glucose, mmol/L | 8.4(2.8);n=19487 | 8.9(3.0);n=25062 | 0.16 | 7.7(2.6);n=11980 | 9.1(3.0);n=19658 | 0.52* | 8.6(2.9);n=23965 | 8.7(3.0);n=20584 | 0.03 | 8.3(2.9);n=19167 | 9.0(3.1);n=12471 | 0.25* |
| Variance of fasting glucose | 9.3(28.8);n=11155 | 8.3(31.9);n=15796 | 0.03 | 10.1(47.1);n=5809 | 8.9(36.0);n=14014 | 0.03 | 8.8(32.6);n=14701 | 8.6(28.0);n=12250 | 0.01 | 8.0(39.6);n=10968 | 10.9(39.4);n=8855 | 0.07 |
| SD of fasting glucose | 2.1(2.2);n=11166 | 1.9(2.1);n=15813 | 0.06 | 1.86(2.58);n=5816 | 1.88(2.32);n=14025 | 0.01 | 1.98(2.21);n=14719 | 2.0(2.13);n=12260 | 0.01 | 1.6(2.3);n=10980 | 2.2(2.5);n=8861 | 0.21* |
| SD/Initial fasting glucose | 0.25(0.31);n=11166 | 0.21(0.24);n=15813 | 0.15 | 0.21(0.24);n=5816 | 0.18(0.21);n=14025 | 0.13 | 0.22(0.28);n=14719 | 0.23(0.27);n=12260 | 0.03 | 0.17(0.22);n=10980 | 0.21(0.22);n=8861 | 0.18 |
| CV of fasting glucose | 0.18(0.15);n=10877 | 0.16(0.13);n=15532 | 0.12 | 0.2(0.2);n=5605 | 0.1(0.1);n=13826 | 0.09 | 0.17(0.14);n=14378 | 0.17(0.14);n=12031 | 0.02 | 0.1(0.1);n=10750 | 0.2(0.1);n=8681 | 0.2 |
| Variability independent of mean fasting glucose | 0.5(0.4);n=11166 | 0.4(0.4);n=15813 | 0.1 | 0.43(0.45);n=5816 | 0.42(0.41);n=14025 | 0.03 | 0.46(0.41);n=14719 | 0.47(0.4);n=12260 | 0.01 | 0.4(0.4);n=10980 | 0.5(0.4);n=8861 | 0.23* |

**Supplementary Table 7. Univariable Cox regression models to predict heart failure and myocardial infarction before and after 1:1 matching.**

* for p≤ 0.05, ** for p ≤ 0.01, *** for p ≤ 0.001; HR: hazard ratio; CI: confidence interval; SD: standard deviation; SCD: sudden cardiac death; VF: ventricular fibrillation; VT: ventricular tachycardia; SGLT2I: sodium glucose cotransporter-2 inhibitor; DPP4I: dipeptidyl peptidase-4 inhibitor; CV: coefficient of variation.

| **Characteristics** | **Before matching** |  | **After matching** |  |
| --- | --- | --- | --- | --- |
|  | **Myocardial infarction**  **HR [95% CI];P value** | **Heart failure**  **HR [95% CI];P value** | **Myocardial infarction**  **HR [95% CI];P value** | **Heart failure**  **HR [95% CI];P value** |
| ***Demographics*** |  |  |  |  |
| Male gender | 1.18[1.09-1.27];<0.0001*** | 0.85[0.80-0.91];<0.0001*** | 1.96[1.75-2.20];<0.0001*** | 1.07[0.97-1.17];0.1996 |
| Female gender | 1.0[Reference] | 1.0[Reference] | 1.0[Reference] | 1.0[Reference] |
| Baseline age, years | 1.05[1.05-1.06];<0.0001*** | 1.08[1.07-1.08];<0.0001*** | 1.03[1.03-1.04];<0.0001*** | 1.050[1.046-1.053];<0.0001*** |
| <50 | 1.0[Reference] | 1.0[Reference] | 1.0[Reference] | 1.0[Reference] |
| [50-60] | 0.53[0.47-0.58];<0.0001*** | 0.32[0.29-0.35];<0.0001*** | 0.77[0.69-0.86];<0.0001*** | 0.24[0.21-0.28];<0.0001*** |
| [60-70] | 0.69[0.63-0.75];<0.0001*** | 0.62[0.57-0.67];<0.0001*** | 0.56[0.49-0.64];<0.0001*** | 1.26[1.14-1.40];<0.0001*** |
| [70-80] | 1.78[1.63-1.94];<0.0001*** | 2.00[1.86-2.15];<0.0001*** | 4.48[4.06-4.95];<0.0001*** | 4.68[4.26-5.15];<0.0001*** |
| >80 | 3.42[3.13-3.73];<0.0001*** | 5.12[4.77-5.49];<0.0001*** | 0.99[0.85-1.16];0.9328 | 1.28[1.12-1.46];0.0003*** |
| ***Past comorbidities*** |  |  |  |  |
| Charlson standard comorbidity index | 1.35[1.33-1.38];<0.0001*** | 1.45[1.43-1.47];<0.0001*** | 1.28[1.24-1.32];<0.0001*** | 1.40[1.36-1.43];<0.0001*** |
| Diabetes with chronic complication | 2.11[1.61-2.77];<0.0001*** | 2.00[1.57-2.54];<0.0001*** | 2.56[1.90-3.45];<0.0001*** | 2.30[1.70-3.12];<0.0001*** |
| Diabetes without chronic complication | 1.23[0.94-1.61];0.1279 | 1.48[1.19-1.83];0.0003*** | 1.39[1.04-1.86];0.0251* | 1.63[1.26-2.10];0.0002*** |
| Gout | 2.17[1.82-2.59];<0.0001*** | 2.52[2.18-2.91];<0.0001*** | 0.91[0.63-1.31];0.6072 | 2.33[1.86-2.92];<0.0001*** |
| Hyperlipidaemia | 1.07[0.85-1.35];0.5799 | 0.85[0.68-1.06];0.1561 | 0.67[0.49-0.92];0.0124* | 0.42[0.28-0.61];<0.0001*** |
| Hypertension | 1.65[1.52-1.79];<0.0001*** | 1.91[1.78-2.05];<0.0001*** | 0.62[0.54-0.71];<0.0001*** | 1.53[1.38-1.69];<0.0001*** |
| Hypoglycemia | 2.97[2.25-3.91];<0.0001*** | 2.53[1.96-3.27];<0.0001*** | 0.54[0.14-2.18];0.3895 | 0.99[0.37-2.63];0.9791 |
| Immune mediated enterocolitis | 1.37[1.17-1.60];0.0001*** | 1.13[0.98-1.31];0.0893 | 0.77[0.60-0.99];0.0385* | 0.74[0.58-0.94];0.0153* |
| Ischemic heart disease | 1.91[1.69-2.14];<0.0001*** | 1.75[1.58-1.95];<0.0001*** | 3.34[2.99-3.74];<0.0001*** | 2.29[2.03-2.58];<0.0001*** |
| Liver diseases | 0.63[0.46-0.88];0.0068** | 0.84[0.65-1.07];0.1594 | 0.37[0.24-0.58];<0.0001*** | 0.58[0.41-0.82];0.0020** |
| Peripheral vascular disease | 3.64[2.79-4.75];<0.0001*** | 3.47[2.74-4.40];<0.0001*** | 2.33[1.42-3.81];0.0008*** | 2.51[1.60-3.94];0.0001*** |
| Renal diseases | 3.38[2.82-4.04];<0.0001*** | 2.58[2.17-3.07];<0.0001*** | 0.98[0.49-1.97];0.9635 | 1.36[0.77-2.40];0.2899 |
| Stroke/transient ischemic attack | 1.85[1.56-2.19];<0.0001*** | 2.17[1.89-2.49];<0.0001*** | 1.30[0.98-1.72];0.0639 | 1.85[1.47-2.32];<0.0001*** |
| Atrial fibrillation | 1.92[1.54-2.40];<0.0001*** | 4.46[3.89-5.10];<0.0001*** | 1.49[1.08-2.07];0.0158* | 3.91[3.19-4.79];<0.0001*** |
| VT/VF/SCD | 1.49[0.56-3.98];0.4244 | 2.33[1.16-4.65];0.0171* | 1.38[0.44-4.27];0.5807 | 1.70[0.64-4.54];0.2878 |
| Anaemia | 1.97[1.69-2.29];<0.0001*** | 2.34[2.07-2.65];<0.0001*** | 1.77[1.37-2.28];<0.0001*** | 1.87[1.47-2.37];<0.0001*** |
| Overweight | 0.45[0.22-0.89];0.0229* | 0.80[0.51-1.26];0.3449 | 0.23[0.10-0.51];0.0003*** | 1.22[0.87-1.71];0.2488 |
| Cancer | 1.15[0.92-1.44];0.2209 | 1.58[1.33-1.87];<0.0001*** | 0.41[0.24-0.70];0.0010** | 1.16[0.85-1.57];0.3436 |
| ***Medications*** |  |  |  |  |
| SGLT2I v.s. DPP4I | 0.56[0.51-0.61];<0.0001*** | 0.39[0.36-0.42];<0.0001*** | 0.60[0.54-0.66];<0.0001*** | 0.52[0.48-0.58];<0.0001*** |
| SGLT2I frequency | 1.00[1.00-1.01];0.2400 | 1.01[1.00-1.01];0.0654 | 1.00[1.00-1.01];0.2400 | 1.01[1.00-1.01];0.0654 |
| DPP4I frequency | 1.00[0.99-1.00];0.1800 | 1.00[1.00-1.01];0.3181 | 0.96[0.95-0.97];<0.0001*** | 0.97[0.96-0.98];<0.0001*** |
| SGLT2I duration, days | 1.000[0.999-1.000];<0.0001*** | 0.999[0.999-1.000];<0.0001*** | 1.000[0.999-1.000];<0.0001*** | 0.999[0.999-1.000];<0.0001*** |
| DPP4I duration, days | 1.000[1.000-1.000];<0.0001*** | 1.000[1.000-1.000];<0.0001*** | 0.997[0.997-0.998];<0.0001*** | 0.999[0.999-1.000];<0.0001*** |
| Metformin | 0.40[0.36-0.44];<0.0001*** | 0.37[0.35-0.41];<0.0001*** | 0.83[0.70-0.99];0.0366* | 0.43[0.38-0.49];<0.0001*** |
| Sulphonylurea | 1.07[0.97-1.17];0.1581 | 1.03[0.95-1.12];0.4325 | 0.54[0.49-0.59];<0.0001*** | 0.44[0.40-0.48];<0.0001*** |
| Insulin | 4.78[4.32-5.27];<0.0001*** | 4.16[3.84-4.52];<0.0001*** | 2.18[1.96-2.41];<0.0001*** | 6.21[5.43-7.09];<0.0001*** |
| Acarbose | 0.89[0.68-1.15];0.3580 | 0.74[0.58-0.94];0.0142* | 2.07[1.73-2.47];<0.0001*** | 0.51[0.37-0.71];<0.0001*** |
| Thiazolidinedione | 0.39[0.34-0.44];<0.0001*** | 0.44[0.40-0.50];<0.0001*** | 0.26[0.22-0.31];<0.0001*** | 0.41[0.36-0.47];<0.0001*** |
| Glucagon-like peptide-1 receptor agonists | 0.42[0.30-0.59];<0.0001*** | 0.37[0.27-0.51];<0.0001*** | 0.37[0.27-0.50];<0.0001*** | 1.24[1.05-1.47];0.0113* |
| Statins and fibrates | 0.46[0.42-0.50];<0.0001*** | 0.37[0.34-0.40];<0.0001*** | 0.58[0.53-0.64];<0.0001*** | 0.47[0.43-0.52];<0.0001*** |
| ***Subclinical biomarkers*** |  |  |  |  |
| Neutrophil-to-lymphocyte ratio | 1.03[1.03-1.04];<0.0001*** | 1.03[1.02-1.03];<0.0001*** | 1.02[1.01-1.04];0.0033** | 1.05[1.04-1.05];<0.0001*** |
| Platelet-to-lymphocyte ratio | 1.000[1.000-1.001];<0.0001*** | 1.000[1.000-1.000];<0.0001*** | 1.000[1.000-1.001];0.5493 | 1.000[0.999-1.001];0.9423 |
| Neutrophil-to-high-density lipoprotein ratio | 1.53[1.39-1.68];<0.0001*** | 1.50[1.37-1.64];<0.0001*** | 1.66[1.43-1.93];<0.0001*** | 1.96[1.80-2.13];<0.0001*** |
| Lymphocyte-to-high-density lipoprotein ratio | 0.09[0.03-0.30];0.0001*** | 0.01[0.00-0.04];<0.0001*** | 5.80[3.08-10.91];<0.0001*** | 9.28[6.06-14.23];<0.0001*** |
| Lymphocyte-to-low-density lipoprotein ratio | 0.38[0.25-0.58];<0.0001*** | 0.46[0.28-0.74];0.0015** | 12.27[2.78-54.19];0.0009*** | 55.27[24.83-123.04];<0.0001*** |
| Low density lipoprotein ratio-to-high density lipoprotein ratio | 1.17[1.13-1.20];<0.0001*** | 0.97[0.93-1.02];0.2091 | 1.30[1.24-1.37];<0.0001*** | 1.13[1.06-1.20];0.0001*** |
| Total cholesterol-to-high density lipoprotein ratio | 1.12[1.09-1.14];<0.0001*** | 0.99[0.96-1.02];0.6082 | 1.18[1.14-1.21];<0.0001*** | 1.08[1.03-1.12];0.0004*** |
| Triglyceride-glucose index | 1.10[1.03-1.17];0.0036** | 0.93[0.88-0.99];0.0139* | 1.16[1.07-1.25];0.0002*** | 1.13[1.05-1.23];0.0015** |
| Bilirubin-to-albumin ratio | 0.77[0.56-1.06];0.1138 | 1.11[1.01-1.22];0.0345* | 0.04[0.02-0.07];<0.0001*** | 1.27[1.07-1.50];0.0074** |
| Protein-to-creatinine ratio | 0.69[0.66-0.72];<0.0001*** | 0.66[0.63-0.68];<0.0001*** | 0.84[0.79-0.89];<0.0001*** | 0.69[0.65-0.73];<0.0001*** |
| Prognostic nutritional index | 0.97[0.96-0.97];<0.0001*** | 0.97[0.96-0.97];<0.0001*** | 1.03[1.02-1.05];<0.0001*** | 0.97[0.96-0.98];<0.0001*** |
| ***Complete blood counts*** |  |  |  |  |
| Mean corpuscular volume, fL | 1.02[1.01-1.03];<0.0001*** | 1.02[1.01-1.02];<0.0001*** | 1.10[1.09-1.11];<0.0001*** | 1.03[1.02-1.04];<0.0001*** |
| Eosinophil, x10^9/L | 1.17[1.04-1.31];0.0068** | 1.20[1.08-1.33];0.0006*** | 0.12[0.07-0.21];<0.0001*** | 0.32[0.21-0.50];<0.0001*** |
| Lymphocyte, x10^9/L | 0.70[0.65-0.75];<0.0001*** | 0.65[0.61-0.69];<0.0001*** | 0.96[0.88-1.06];0.4724 | 1.11[1.05-1.16];0.0001*** |
| Neutrophil, x10^9/L | 1.06[1.05-1.07];<0.0001*** | 1.05[1.04-1.06];<0.0001*** | 1.06[1.03-1.08];<0.0001*** | 1.13[1.12-1.15];<0.0001*** |
| White cell count, x10^9/L | 1.02[1.02-1.03];<0.0001*** | 1.02[1.02-1.03];<0.0001*** | 1.04[1.01-1.06];0.0057** | 1.12[1.11-1.14];<0.0001*** |
| Mean cell haemoglobin, pg | 1.06[1.04-1.08];<0.0001*** | 1.04[1.02-1.05];<0.0001*** | 1.25[1.21-1.29];<0.0001*** | 1.10[1.07-1.13];<0.0001*** |
| Platelet, x10^9/L | 0.999[0.998-1.000];0.0047** | 0.997[0.997-0.998];<0.0001*** | 0.998[0.997-0.999];0.0024** | 1.00[0.99-1.00];<0.0001*** |
| Red cell count, x10^12/L | 0.51[0.47-0.54];<0.0001*** | 0.47[0.44-0.50];<0.0001*** | 0.75[0.67-0.83];<0.0001*** | 0.78[0.70-0.86];<0.0001*** |
| ***Liver and renal functions*** |  |  |  |  |
| Potassium, mmol/L | 1.06[0.97-1.15];0.2222 | 1.08[1.00-1.16];0.0547 | 0.98[0.89-1.08];0.6892 | 0.66[0.59-0.73];<0.0001*** |
| Albumin, g/L | 0.89[0.88-0.90];<0.0001*** | 0.88[0.88-0.89];<0.0001*** | 1.05[1.03-1.07];<0.0001*** | 0.91[0.89-0.92];<0.0001*** |
| Sodium, mmol/L | 0.96[0.95-0.97];<0.0001*** | 0.98[0.96-0.99];0.0001*** | 1.22[1.20-1.24];<0.0001*** | 1.13[1.11-1.15];<0.0001*** |
| Urea, mmol/L | 1.08[1.08-1.09];<0.0001*** | 1.08[1.08-1.09];<0.0001*** | 1.07[1.06-1.08];<0.0001*** | 1.08[1.07-1.09];<0.0001*** |
| Protein, g/L | 0.95[0.94-0.96];<0.0001*** | 0.96[0.95-0.96];<0.0001*** | 0.96[0.95-0.98];<0.0001*** | 0.99[0.97-1.00];0.0393* |
| Creatinine, umol/L | 1.002[1.002-1.003];<0.0001*** | 1.002[1.002-1.002];<0.0001*** | 1.004[1.004-1.004];<0.0001*** | 1.004[1.004-1.005];<0.0001*** |
| Alkaline phosphatase, U/L | 1.004[1.003-1.004];<0.0001*** | 1.003[1.002-1.004];<0.0001*** | 1.00[0.99-1.00];0.0040** | 1.007[1.006-1.008];<0.0001*** |
| Aspartate transaminase, U/L | 1.001[1.001-1.002];<0.0001*** | 1.000[0.999-1.001];0.7658 | 0.96[0.95-0.97];<0.0001*** | 0.999[0.995-1.003];0.6592 |
| Alanine transaminase, U/L | 0.988[0.985-0.991];<0.0001*** | 0.99[0.98-0.99];<0.0001*** | 0.97[0.96-0.97];<0.0001*** | 0.98[0.98-0.99];<0.0001*** |
| Bilirubin, umol/L | 0.97[0.96-0.98];<0.0001*** | 0.99[0.98-1.00];0.0104* | 0.93[0.91-0.94];<0.0001*** | 1.00[0.99-1.01];0.5222 |
| ***Lipid and glucose profiles*** |  |  |  |  |
| Triglyceride, mmol/L | 1.03[1.01-1.05];0.0141* | 0.97[0.94-1.00];0.0629 | 1.02[0.99-1.05];0.1309 | 1.00[0.96-1.03];0.9458 |
| SD of triglyceride | 1.04[0.99-1.09];0.0824 | 0.94[0.87-1.01];0.0773 | 1.11[1.08-1.15];<0.0001*** | 1.07[1.02-1.12];0.0050** |
| Low-density lipoprotein, mmol/L | 1.11[1.05-1.17];0.0001*** | 0.87[0.83-0.92];<0.0001*** | 1.07[1.00-1.15];0.0593 | 1.07[1.00-1.15];0.0520 |
| SD of low-density lipoprotein | 1.49[1.28-1.73];<0.0001*** | 1.14[0.99-1.33];0.0707 | 1.62[1.26-2.08];0.0001*** | 2.07[1.73-2.49];<0.0001*** |
| High-density lipoprotein, mmol/L | 0.63[0.54-0.73];<0.0001*** | 0.86[0.76-0.97];0.0144* | 0.31[0.26-0.38];<0.0001*** | 0.67[0.56-0.79];<0.0001*** |
| SD of high-density lipoprotein | 6.11[3.48-10.73];<0.0001*** | 8.84[5.52-14.17];<0.0001*** | 5.15[1.59-16.73];0.0063** | 31.72[14.88-67.61];<0.0001*** |
| Total cholesterol, mmol/L | 1.07[1.02-1.11];0.0024** | 0.89[0.85-0.92];<0.0001*** | 0.97[0.92-1.03];0.3232 | 1.03[0.97-1.08];0.3681 |
| SD of total cholesterol | 1.40[1.26-1.55];<0.0001*** | 1.18[1.06-1.32];0.0033** | 1.35[1.14-1.60];0.0006*** | 1.61[1.45-1.78];<0.0001*** |
| HbA1c, g/dL | 0.80[0.79-0.82];<0.0001*** | 0.77[0.76-0.79];<0.0001*** | 1.05[1.01-1.09];0.0086** | 0.98[0.94-1.02];0.2501 |
| Mean HbA1c, g/dL | 0.76[0.74-0.78];<0.0001*** | 0.73[0.72-0.75];<0.0001*** | 1.04[1.00-1.08];0.0347* | 0.95[0.92-0.99];0.0062** |
| Median HbA1c, g/dL | 0.77[0.75-0.79];<0.0001*** | 0.73[0.72-0.75];<0.0001*** | 1.03[0.99-1.07];0.1053 | 0.95[0.92-0.99];0.0103* |
| Variance of HbA1c | 1.12[1.09-1.16];<0.0001*** | 1.10[1.06-1.13];<0.0001*** | 1.18[1.15-1.22];<0.0001*** | 1.07[1.01-1.14];0.0256* |
| SD of HbA1c | 1.72[1.56-1.89];<0.0001*** | 1.59[1.46-1.73];<0.0001*** | 2.06[1.75-2.42];<0.0001*** | 1.28[1.08-1.51];0.0040** |
| SD/Initial HbA1c | 71.56[36.27-141.18];<0.0001*** | 51.22[27.86-94.15];<0.0001*** | 326.08[85.70-1240.78];<0.0001*** | 12.45[2.80-55.40];0.0009*** |
| CV of HbA1c | 1304.97[507.90-3352.91];<0.0001*** | 588.00[256.26-1349.18];<0.0001*** | 3950.62[617.90-25258.96];<0.0001*** | 21.81[3.31-143.80];0.0014** |
| Variability independent of mean HbA1c | 3.42[2.79-4.17];<0.0001*** | 2.91[2.44-3.46];<0.0001*** | 4.74[3.32-6.77];<0.0001*** | 1.67[1.17-2.40];0.0053** |
| Fasting glucose, mmol/L | 1.02[1.01-1.03];<0.0001*** | 1.01[1.00-1.02];0.2111 | 0.99[0.98-1.00];0.0781 | 1.01[1.00-1.02];0.1695 |
| Mean fasting glucose, mmol/L | 1.04[1.03-1.05];<0.0001*** | 1.01[1.00-1.02];0.0612 | 0.99[0.98-1.01];0.4659 | 1.00[0.98-1.02];0.9065 |
| Median fasting glucose, mmol/L | 1.03[1.01-1.04];0.0002*** | 1.00[0.98-1.01];0.7027 | 0.99[0.97-1.01];0.3742 | 0.98[0.96-1.00];0.0561 |
| Variance of fasting glucose | 1.003[1.002-1.003];<0.0001*** | 1.002[1.002-1.003];<0.0001*** | 1.002[1.001-1.003];0.0001*** | 1.003[1.002-1.004];<0.0001*** |
| SD of fasting glucose | 1.11[1.09-1.13];<0.0001*** | 1.10[1.09-1.11];<0.0001*** | 1.09[1.06-1.11];<0.0001*** | 1.10[1.08-1.11];<0.0001*** |
| SD/Initial fasting glucose | 1.71[1.58-1.85];<0.0001*** | 1.75[1.64-1.87];<0.0001*** | 1.76[1.54-2.00];<0.0001*** | 1.71[1.53-1.92];<0.0001*** |
| CV of fasting glucose | 12.02[8.91-16.22];<0.0001*** | 11.87[9.19-15.33];<0.0001*** | 8.43[5.46-13.01];<0.0001*** | 8.13[5.72-11.56];<0.0001*** |
| Variability independent of mean fasting glucose | 2.13[1.93-2.35];<0.0001*** | 2.06[1.90-2.24];<0.0001*** | 1.88[1.63-2.17];<0.0001*** | 1.88[1.67-2.11];<0.0001*** |

**Supplementary Table 8. Univariable Cox regression models to predict cardiovascular mortality and all-cause mortality before and after 1:1 matching.**

* for p≤ 0.05, ** for p ≤ 0.01, *** for p ≤ 0.001; HR: hazard ratio; CI: confidence interval; SD: standard deviation; SCD: sudden cardiac death; VF: ventricular fibrillation; VT: ventricular tachycardia; SGLT2I: sodium glucose cotransporter-2 inhibitor; DPP4I: dipeptidyl peptidase-4 inhibitor; CV: coefficient of variation.

| **Characteristics** | **Before matching** |  | **After matching** |  |
| --- | --- | --- | --- | --- |
|  | **All-cause mortality**  **HR [95% CI];P value** | **Cardiovascular mortality**  **HR [95% CI];P value** | **All-cause mortality**  **HR [95% CI];P value** | **Cardiovascular mortality**  **HR [95% CI];P value** |
| ***Demographics*** |  |  |  |  |
| Male gender | 0.98[0.94-1.04];0.5475 | 1.15[1.05-1.26];0.0035** | 0.91[0.84-0.98];0.0143* | 0.96[0.81-1.14];0.6465 |
| Female gender | 1.0[Reference] | 1.0[Reference] | 1.0[Reference] | 1.0[Reference] |
| Baseline age, years | 1.09[1.09-1.10];<0.0001*** | 1.13[1.13-1.14];<0.0001*** | 1.049[1.046-1.052];<0.0001*** | 1.16[1.15-1.17];<0.0001*** |
| <50 | 1.0[Reference] | 1.0[Reference] | 1.0[Reference] | 1.0[Reference] |
| [50-60] | 0.26[0.24-0.28];<0.0001*** | 0.13[0.10-0.16];<0.0001*** | 0.56[0.51-0.61];<0.0001*** | 0.04[0.02-0.07];<0.0001*** |
| [60-70] | 0.54[0.51-0.58];<0.0001*** | 0.34[0.30-0.39];<0.0001*** | 0.31[0.27-0.35];<0.0001*** | 0.41[0.32-0.53];<0.0001*** |
| [70-80] | 1.96[1.86-2.07];<0.0001*** | 1.83[1.66-2.03];<0.0001*** | 3.51[3.24-3.80];<0.0001*** | 2.63[2.17-3.18];<0.0001*** |
| >80 | 6.80[6.46-7.15];<0.0001*** | 11.69[10.68-12.81];<0.0001*** | 2.60[2.38-2.84];<0.0001*** | 11.93[10.04-14.16];<0.0001*** |
| ***Past comorbidities*** |  |  |  |  |
| Charlson standard comorbidity index | 1.54[1.53-1.56];<0.0001*** | 1.61[1.58-1.63];<0.0001*** | 1.42[1.39-1.45];<0.0001*** | 1.79[1.74-1.85];<0.0001*** |
| Diabetes with chronic complication | 2.07[1.73-2.48];<0.0001*** | 1.56[1.07-2.26];0.0197* | 1.67[1.26-2.23];0.0004*** | 2.83[1.72-4.65];<0.0001*** |
| Diabetes without chronic complication | 1.53[1.31-1.80];<0.0001*** | 1.78[1.36-2.33];<0.0001*** | 1.81[1.48-2.21];<0.0001*** | 5.16[3.89-6.85];<0.0001*** |
| Gout | 2.30[2.06-2.57];<0.0001*** | 2.93[2.44-3.53];<0.0001*** | 1.65[1.33-2.05];<0.0001*** | 2.20[1.45-3.35];0.0002*** |
| Hyperlipidaemia | 0.83[0.70-0.99];0.0385* | 0.89[0.65-1.20];0.4308 | 0.54[0.41-0.71];<0.0001*** | 0.78[0.47-1.31];0.3548 |
| Hypertension | 1.92[1.82-2.02];<0.0001*** | 2.15[1.95-2.36];<0.0001*** | 1.47[1.35-1.59];<0.0001*** | 3.33[2.81-3.94];<0.0001*** |
| Hypoglycemia | 3.42[2.89-4.05];<0.0001*** | 3.89[2.91-5.19];<0.0001*** | 3.23[2.06-5.07];<0.0001*** | 6.12[2.91-12.91];<0.0001*** |
| Immune mediated enterocolitis | 1.21[1.09-1.34];0.0005*** | 1.36[1.13-1.63];0.0012** | 1.04[0.88-1.24];0.6283 | 1.79[1.33-2.42];0.0001*** |
| Ischemic heart disease | 1.21[1.11-1.33];<0.0001*** | 1.17[0.99-1.39];0.0653 | 1.09[0.96-1.23];0.1931 | 1.97[1.57-2.47];<0.0001*** |
| Liver diseases | 1.04[0.88-1.24];0.6093 | 0.85[0.60-1.19];0.3375 | 0.73[0.57-0.95];0.0171* | 0.73[0.41-1.29];0.2749 |
| Peripheral vascular disease | 3.75[3.15-4.46];<0.0001*** | 3.78[2.77-5.17];<0.0001*** | 4.36[3.29-5.79];<0.0001*** | 3.20[1.52-6.74];0.0022** |
| Renal diseases | 4.32[3.89-4.81];<0.0001*** | 4.02[3.29-4.91];<0.0001*** | 2.13[1.47-3.09];0.0001*** | 6.66[4.11-10.80];<0.0001*** |
| Stroke/transient ischemic attack | 2.38[2.15-2.63];<0.0001*** | 3.03[2.57-3.57];<0.0001*** | 2.20[1.86-2.62];<0.0001*** | 7.15[5.64-9.07];<0.0001*** |
| Atrial fibrillation | 2.65[2.34-3.01];<0.0001*** | 3.37[2.75-4.14];<0.0001*** | 3.01[2.50-3.62];<0.0001*** | 6.85[5.12-9.17];<0.0001*** |
| VT/VF/SCD | 2.74[1.70-4.41];<0.0001*** | 2.12[0.79-5.66];0.1332 | 3.68[2.14-6.35];<0.0001*** | 5.79[2.16-15.48];0.0005*** |
| Anaemia | 3.06[2.81-3.33];<0.0001*** | 3.40[2.94-3.94];<0.0001*** | 2.16[1.80-2.59];<0.0001*** | 3.73[2.70-5.15];<0.0001*** |
| Overweight | 0.33[0.19-0.55];<0.0001*** | 0.08[0.01-0.55];0.0104* | 0.49[0.32-0.76];0.0013** | 0.00[0.00-Inf];0.9807 |
| Cancer | 2.29[2.06-2.55];<0.0001*** | 2.19[1.80-2.68];<0.0001*** | 2.03[1.67-2.47];<0.0001*** | 3.00[2.09-4.31];<0.0001*** |
| ***Medications*** |  |  |  |  |
| SGLT2I v.s. DPP4I | 0.16[0.15-0.17];<0.0001*** | 0.10[0.08-0.12];<0.0001*** | 0.23[0.21-0.26];<0.0001*** | 0.23[0.18-0.28];<0.0001*** |
| SGLT2I frequency | 1.02[1.01-1.02];<0.0001*** | 1.02[1.01-1.03];<0.0001*** | 1.02[1.01-1.02];<0.0001*** | 1.02[1.01-1.03];<0.0001*** |
| DPP4I frequency | 0.99[0.99-1.00];0.0018** | 1.00[0.99-1.00];0.2615 | 0.92[0.91-0.93];<0.0001*** | 1.00[0.99-1.01];0.8523 |
| SGLT2I duration, days | 1.000[1.000-1.000];0.0123* | 1.000[0.999-1.000];0.1366 | 1.000[1.000-1.000];0.0123* | 1.000[0.999-1.000];0.1366 |
| DPP4I duration, days | 1.000[1.000-1.000];<0.0001*** | 1.000[0.999-1.000];<0.0001*** | 0.998[0.998-0.998];<0.0001*** | 1.002[1.002-1.002];<0.0001*** |
| Metformin | 0.28[0.27-0.30];<0.0001*** | 0.28[0.25-0.31];<0.0001*** | 0.51[0.46-0.58];<0.0001*** | 0.26[0.21-0.32];<0.0001*** |
| Sulphonylurea | 1.06[1.00-1.13];0.0464* | 1.29[1.15-1.45];<0.0001*** | 0.36[0.33-0.39];<0.0001*** | 0.74[0.62-0.89];0.0012** |
| Insulin | 5.11[4.79-5.46];<0.0001*** | 7.85[6.83-9.03];<0.0001*** | 9.85[8.65-11.23];<0.0001*** | 6.71[5.23-8.61];<0.0001*** |
| Acarbose | 0.98[0.83-1.15];0.7939 | 0.95[0.71-1.28];0.7327 | 1.08[0.90-1.30];0.4066 | 2.86[2.17-3.77];<0.0001*** |
| Thiazolidinedione | 0.38[0.35-0.41];<0.0001*** | 0.27[0.23-0.33];<0.0001*** | 1.53[1.41-1.65];<0.0001*** | 0.17[0.12-0.24];<0.0001*** |
| Glucagon-like peptide-1 receptor agonists | 0.11[0.07-0.16];<0.0001*** | 0.05[0.02-0.16];<0.0001*** | 0.31[0.24-0.40];<0.0001*** | 1.15[0.85-1.58];0.3667 |
| Statins and fibrates | 0.30[0.29-0.32];<0.0001*** | 0.25[0.22-0.28];<0.0001*** | 0.46[0.42-0.49];<0.0001*** | 0.51[0.43-0.60];<0.0001*** |
| ***Subclinical biomarkers*** |  |  |  |  |
| Neutrophil-to-lymphocyte ratio | 1.04[1.03-1.04];<0.0001*** | 1.04[1.03-1.05];<0.0001*** | 1.03[1.02-1.04];<0.0001*** | 1.05[1.04-1.06];<0.0001*** |
| Platelet-to-lymphocyte ratio | 1.000[1.000-1.000];<0.0001*** | 1.000[1.000-1.001];<0.0001*** | 1.000[1.000-1.001];0.0051** | 1.000[1.000-1.001];0.0423* |
| Neutrophil-to-high-density lipoprotein ratio | 1.41[1.30-1.53];<0.0001*** | 1.42[1.23-1.65];<0.0001*** | 0.80[0.52-1.21];0.2890 | 0.82[0.34-1.97];0.6551 |
| Lymphocyte-to-high-density lipoprotein ratio | 0.000[0.000-0.001];<0.0001*** | 0.000[0.000-0.000];<0.0001*** | 0.000[0.000-0.000];<0.0001*** | 0.000[0.000-0.000];<0.0001*** |
| Lymphocyte-to-low-density lipoprotein ratio | 0.34[0.27-0.44];<0.0001*** | 0.33[0.22-0.50];<0.0001*** | 16.93[5.99-47.88];<0.0001*** | 0.86[0.44-1.68];0.6557 |
| Low density lipoprotein ratio-to-high density lipoprotein ratio | 0.98[0.95-1.02];0.3466 | 0.86[0.80-0.92];<0.0001*** | 0.66[0.62-0.71];<0.0001*** | 0.53[0.45-0.63];<0.0001*** |
| Total cholesterol-to-high density lipoprotein ratio | 0.98[0.95-1.00];0.0700 | 0.88[0.84-0.93];<0.0001*** | 0.67[0.64-0.71];<0.0001*** | 0.60[0.53-0.68];<0.0001*** |
| Triglyceride-glucose index | 0.88[0.84-0.92];<0.0001*** | 0.78[0.72-0.85];<0.0001*** | 0.58[0.54-0.62];<0.0001*** | 0.70[0.59-0.84];0.0001*** |
| Bilirubin-to-albumin ratio | 1.15[1.09-1.21];<0.0001*** | 1.04[0.86-1.27];0.6732 | 1.07[0.82-1.41];0.6064 | 0.97[0.49-1.92];0.9328 |
| Protein-to-creatinine ratio | 0.61[0.59-0.63];<0.0001*** | 0.63[0.60-0.66];<0.0001*** | 1.09[1.05-1.13];<0.0001*** | 0.46[0.42-0.51];<0.0001*** |
| Prognostic nutritional index | 0.959[0.957-0.962];<0.0001*** | 0.96[0.95-0.96];<0.0001*** | 0.96[0.95-0.97];<0.0001*** | 0.93[0.92-0.94];<0.0001*** |
| ***Complete blood counts*** |  |  |  |  |
| Mean corpuscular volume, fL | 1.02[1.02-1.03];<0.0001*** | 1.02[1.01-1.03];<0.0001*** | 1.08[1.07-1.09];<0.0001*** | 1.08[1.07-1.10];<0.0001*** |
| Eosinophil, x10^9/L | 1.06[0.94-1.18];0.3601 | 0.89[0.68-1.18];0.4246 | 0.28[0.20-0.40];<0.0001*** | 0.07[0.03-0.18];<0.0001*** |
| Lymphocyte, x10^9/L | 0.53[0.50-0.56];<0.0001*** | 0.47[0.43-0.51];<0.0001*** | 0.97[0.90-1.04];0.3958 | 0.44[0.36-0.54];<0.0001*** |
| Neutrophil, x10^9/L | 1.05[1.05-1.06];<0.0001*** | 1.07[1.05-1.08];<0.0001*** | 1.09[1.08-1.11];<0.0001*** | 1.11[1.08-1.13];<0.0001*** |
| White cell count, x10^9/L | 1.02[1.01-1.02];<0.0001*** | 1.02[1.01-1.03];<0.0001*** | 1.05[1.04-1.07];<0.0001*** | 0.95[0.91-1.00];0.0674 |
| Mean cell haemoglobin, pg | 1.06[1.05-1.07];<0.0001*** | 1.04[1.02-1.06];0.0001*** | 1.19[1.16-1.21];<0.0001*** | 1.22[1.17-1.27];<0.0001*** |
| Platelet, x10^9/L | 0.997[0.997-0.998];<0.0001*** | 0.997[0.996-0.998];<0.0001*** | 1.003[1.002-1.004];<0.0001*** | 0.990[0.988-0.992];<0.0001*** |
| Red cell count, x10^12/L | 0.38[0.37-0.40];<0.0001*** | 0.38[0.35-0.41];<0.0001*** | 0.44[0.40-0.47];<0.0001*** | 0.35[0.30-0.41];<0.0001*** |
| ***Liver and renal functions*** |  |  |  |  |
| Potassium, mmol/L | 1.14[1.07-1.20];<0.0001*** | 1.19[1.07-1.32];0.0010** | 0.71[0.65-0.78];<0.0001*** | 1.20[0.99-1.44];0.0582 |
| Albumin, g/L | 0.859[0.855-0.864];<0.0001*** | 0.85[0.84-0.86];<0.0001*** | 0.93[0.92-0.94];<0.0001*** | 0.84[0.82-0.86];<0.0001*** |
| Sodium, mmol/L | 0.95[0.94-0.96];<0.0001*** | 0.95[0.93-0.96];<0.0001*** | 1.11[1.09-1.13];<0.0001*** | 0.99[0.96-1.03];0.7573 |
| Urea, mmol/L | 1.089[1.085-1.092];<0.0001*** | 1.08[1.08-1.09];<0.0001*** | 1.07[1.07-1.08];<0.0001*** | 1.07[1.06-1.09];<0.0001*** |
| Protein, g/L | 0.95[0.94-0.95];<0.0001*** | 0.94[0.93-0.95];<0.0001*** | 1.03[1.02-1.04];<0.0001*** | 1.01[0.98-1.03];0.6243 |
| Creatinine, umol/L | 1.003[1.002-1.003];<0.0001*** | 1.002[1.002-1.003];<0.0001*** | 1.004[1.004-1.004];<0.0001*** | 1.004[1.003-1.005];<0.0001*** |
| Alkaline phosphatase, U/L | 1.00[1.00-1.01];<0.0001*** | 1.00[1.00-1.01];<0.0001*** | 1.006[1.005-1.007];<0.0001*** | 1.001[0.996-1.005];0.7958 |
| Aspartate transaminase, U/L | 1.000[1.000-1.001];0.4442 | 1.000[0.998-1.002];0.9622 | 1.00[1.00-1.01];0.0001*** | 0.97[0.95-0.99];0.0003*** |
| Alanine transaminase, U/L | 0.99[0.98-0.99];<0.0001*** | 0.97[0.97-0.98];<0.0001*** | 0.99[0.98-0.99];<0.0001*** | 0.92[0.91-0.94];<0.0001*** |
| Bilirubin, umol/L | 0.99[0.98-0.99];0.0001*** | 0.97[0.96-0.98];<0.0001*** | 0.99[0.98-1.00];0.0077** | 0.96[0.93-0.98];0.0002*** |
| ***Lipid and glucose profiles*** |  |  |  |  |
| Triglyceride, mmol/L | 0.94[0.92-0.96];<0.0001*** | 0.87[0.83-0.92];<0.0001*** | 0.68[0.64-0.73];<0.0001*** | 0.75[0.65-0.86];0.0001*** |
| SD of triglyceride | 0.93[0.87-0.98];0.0139* | 0.92[0.81-1.03];0.1504 | 0.93[0.82-1.06];0.2934 | 0.71[0.46-1.10];0.1291 |
| Low-density lipoprotein, mmol/L | 0.95[0.92-0.99];0.0104* | 0.86[0.80-0.93];0.0001*** | 0.89[0.84-0.95];0.0006*** | 0.82[0.71-0.96];0.0151* |
| SD of low-density lipoprotein | 1.23[1.10-1.39];0.0004*** | 1.14[0.91-1.43];0.2557 | 1.02[0.79-1.32];0.8655 | 0.53[0.28-1.01];0.0520 |
| High-density lipoprotein, mmol/L | 1.14[1.05-1.25];0.0026** | 1.47[1.26-1.71];<0.0001*** | 2.17[1.94-2.43];<0.0001*** | 3.23[2.57-4.06];<0.0001*** |
| SD of high-density lipoprotein | 14.91[10.55-21.06];<0.0001*** | 23.22[12.77-42.22];<0.0001*** | 66.87[32.37-138.14];<0.0001*** | 80.96[18.91-346.66];<0.0001*** |
| Total cholesterol, mmol/L | 0.95[0.92-0.98];0.0014** | 0.89[0.84-0.94];0.0001*** | 0.90[0.85-0.95];0.0001*** | 0.97[0.86-1.09];0.6100 |
| SD of total cholesterol | 1.25[1.14-1.36];<0.0001*** | 1.21[1.02-1.43];0.0297* | 1.18[0.98-1.42];0.0782 | 1.13[0.76-1.68];0.5419 |
| HbA1c, g/dL | 0.73[0.72-0.74];<0.0001*** | 0.72[0.70-0.74];<0.0001*** | 0.84[0.82-0.86];<0.0001*** | 0.77[0.73-0.81];<0.0001*** |
| Mean HbA1c, g/dL | 0.68[0.67-0.69];<0.0001*** | 0.67[0.65-0.69];<0.0001*** | 0.84[0.82-0.87];<0.0001*** | 0.71[0.67-0.76];<0.0001*** |
| Median HbA1c, g/dL | 0.69[0.68-0.70];<0.0001*** | 0.68[0.66-0.70];<0.0001*** | 0.84[0.82-0.87];<0.0001*** | 0.73[0.69-0.78];<0.0001*** |
| Variance of HbA1c | 1.13[1.11-1.15];<0.0001*** | 1.12[1.08-1.16];<0.0001*** | 1.07[1.02-1.12];0.0061** | 1.19[1.15-1.23];<0.0001*** |
| SD of HbA1c | 1.79[1.69-1.90];<0.0001*** | 1.72[1.54-1.91];<0.0001*** | 1.08[0.94-1.25];0.2794 | 2.35[1.97-2.81];<0.0001*** |
| SD/Initial HbA1c | 115.61[78.31-170.68];<0.0001*** | 90.18[42.98-189.19];<0.0001*** | 2.12[0.53-8.39];0.2857 | 241.33[44.56-1306.96];<0.0001*** |
| CV of HbA1c | 2622.03[1486.05-4626.40];<0.0001*** | 1698.00[590.18-4885.31];<0.0001*** | 3.87[0.76-19.66];0.1023 | 3569.97[374.59-34023.31];<0.0001*** |
| Variability independent of mean HbA1c | 3.85[3.41-4.36];<0.0001*** | 3.53[2.81-4.42];<0.0001*** | 1.20[0.88-1.63];0.2547 | 6.24[4.22-9.23];<0.0001*** |
| Fasting glucose, mmol/L | 1.02[1.01-1.02];<0.0001*** | 1.02[1.01-1.03];0.0002*** | 0.98[0.97-0.99];0.0028** | 1.01[0.99-1.03];0.5219 |
| Mean fasting glucose, mmol/L | 1.03[1.02-1.04];<0.0001*** | 1.04[1.03-1.06];<0.0001*** | 0.99[0.97-1.00];0.1739 | 1.03[1.00-1.07];0.0441* |
| Median fasting glucose, mmol/L | 1.02[1.01-1.03];<0.0001*** | 1.03[1.02-1.05];0.0001*** | 0.99[0.97-1.00];0.1122 | 1.02[0.99-1.06];0.2145 |
| Variance of fasting glucose | 1.003[1.002-1.003];<0.0001*** | 1.003[1.002-1.003];<0.0001*** | 1.001[0.999-1.002];0.4869 | 1.001[0.998-1.004];0.4351 |
| SD of fasting glucose | 1.11[1.10-1.12];<0.0001*** | 1.12[1.10-1.13];<0.0001*** | 1.04[1.02-1.07];0.0001*** | 1.06[1.01-1.10];0.0076** |
| SD/Initial fasting glucose | 1.77[1.68-1.86];<0.0001*** | 1.79[1.64-1.95];<0.0001*** | 1.65[1.47-1.86];<0.0001*** | 1.52[1.13-2.04];0.0052** |
| CV of fasting glucose | 12.63[10.39-15.36];<0.0001*** | 13.18[9.20-18.88];<0.0001*** | 4.69[3.25-6.76];<0.0001*** | 2.55[1.10-5.91];0.0285* |
| Variability independent of mean fasting glucose | 2.17[2.04-2.32];<0.0001*** | 2.26[2.02-2.53];<0.0001*** | 1.48[1.31-1.68];<0.0001*** | 1.43[1.11-1.85];0.0054** |
